# Adjusting for medication use in GWAS and its impact on Mendelian randomization analyses: an example of systolic blood pressure in UK Biobank

**DOI:** 10.64898/2026.01.09.26343783

**Authors:** Aaron Jun Yi Yap, Aimee L Hanson, Gareth J Griffith, Eleanor Sanderson

## Abstract

Medication use is common in large-scale population cohorts, and can modify phenotypic traits of interest. This can potentially bias effect estimates in genome-wide association studies (GWAS) and impact downstream analyses such as mendelian randomization (MR). The best approach to account for medication use in GWAS is unclear. In this study, we compared seven different methods of adjusting for antihypertensive use in a systolic blood pressure (SBP) GWAS of 407,960 White British individuals in the UK Biobank. We found that direct adjustments to measured SBP (adding constants, class-specific constants, censored normal regression) in general yielded a greater number of genome-wide significant variant associations and unmasked stronger GWAS effect estimates than unadjusted measures of SBP. Adjustment for class-specific constants showed the greatest difference relative to unadjusted GWAS. Restriction methods which limit the sample to either untreated individuals or age ranges with low levels of antihypertensive use had less power, due to reduced sample sizes. Effect estimates of treated individuals were deflated relative to untreated individuals, demonstrating the importance of medication adjustment. In MR analyses, we found no substantial differences in inverse-variance weighted (IVW) estimates when using differing exposure GWAS methods in estimation of the effect of SBP on coronary artery disease. Larger variations in IVW estimates were observed for the causal effect of body mass index on SBP across adjustment approaches. This suggests that bias may arise in MR analyses when the exposure included in the estimation affects the probability of treatment. Finally, we demonstrate that medication adjustment can reveal potentially novel genetic loci, offering additional insight into the biology of a trait.

## 1. Introduction

In the past two decades, genome-wide association studies (GWAS) have been widely conducted with the aim of understanding how genetic variants or single nucleotide polymorphisms (SNPs) in a population are linked to a trait or disease. The primary goal is then to examine and apply this knowledge to better understand disease processes, and devise prevention or treatment strategies to improve population health (Hirschhorn & Daly, 2005; Visscher et al., 2017). A complementary approach to understanding disease aetiology is mendelian randomization (MR), which uses trait associated SNPs from GWAS as instrumental variables (IV) to test for a causal effect of an exposure on an outcome of interest (Davey Smith & Ebrahim, 2003; Sanderson et al., 2022). MR analyses can be conducted using either individual-level data or summary-level GWAS outputs. Summary-level outputs have been more accessibly employed due to a wealth of publicly available GWAS summary statistics (Pierce & Burgess, 2013). Multivariable MR (MVMR) is an extension of MR which allows for the inclusion of two or more exposures in a single estimation (Burgess & Thompson, 2015; Sanderson et al., 2019).

Many recently published GWAS are conducted on, or include in meta-analyses, large-scale population cohorts such as the UK Biobank (Bycroft et al., 2018; Surendran et al., 2020). Medication use is often common in these cohorts. In the UK Biobank, approximately three in four participants had medication use recorded at the baseline assessment visit (Wu et al., 2019). While the design, sampling and quality control of GWAS are frequently discussed, the potential for bias in detected genetic effects arising from medication use is often not considered (Davies et al., 2024; McCarthy et al., 2008; Visscher et al., 2017; Weale, 2010). This is a potential concern when the medications may modify the phenotypic trait of interest. For example, patients who are treated with antihypertensives are expected to have a lower measured systolic blood pressure (SBP) than their untreated SBP. Thus, IV effect estimates taken from a GWAS of measured SBP in a population with substantial medication use may be biased (Barendse, 2011).

Some GWAS attempt to adjust for medication use to reduce this potential bias (Cho et al., 2009; Keaton et al., 2024; Kim et al., 2011; Sabatti et al., 2009; Surendran et al., 2020). Hereafter, we refer to antihypertensive use as ’treatment’, and individuals treated with antihypertensives as ’treated’. The most common approach is to apply a direct adjustment to the phenotypic values in treated individuals to approximate untreated values. In SBP GWAS, this typically involves adding a constant of 10 or 15 mmHg to measured SBP values of individuals who are treated (Keaton et al., 2024; Sabatti et al., 2009; Surendran et al., 2020). Another approach is to conduct GWAS in solely untreated individuals. This may include restriction to untreated individuals, or to younger individuals with an assumed low proportion of treatment. (Cho et al., 2009; Kim et al., 2011). However, conditioning on treatment in an MR analysis may introduce collider bias. Treatment is a downstream consequence of the trait, so conditioning on it can induce associations between the genetic IV and other causes of treatment. This opens a spurious path to the outcome and potentially biases the MR estimate. (Cole et al., 2010). Other previous work have also attempted to quantify and correct this bias. Walker et al. (2021) used simulations to show selecting on medication users can potentially induce bias. Chong et al. (2024) developed a ‘genetic empirical medication reduction approach’ to adjust the observed phenotypic value by conducting GWAS in treated and untreated individuals adjusting for medication use, and choosing the best model from the summed R^2^ value.

It is unclear if the most common approach of adding a uniform constant to all treated individuals is the most optimal approach to adjust measured SBP for treatment. Assuming a homogeneous treatment effect among all participants who are using a differing number and type of antihypertensives may be an oversimplification. The National Institute for Health and Care Excellence (NICE) in the UK provides guidance to add combinatory therapy in patients with inadequate control of hypertension. As such a greater number of antihypertensives may suggest more severe hypertension and higher untreated SBP (National Institute for Health and Care Excellence, 2019). Hence, taking into consideration the number and type of antihypertensives used could be a more reasonable approach. Tobin et al. (2005) have suggested alternatives, such as using a right-censored normal regression which derives the untreated SBP value in the treated using various predictors modelled in the untreated population.

MVMR can adequately adjust for bias arising from covariate-adjustment in a GWAS (Gilbody et al., 2025). Bias from covariate adjustment in a GWAS can arise when genetic variants influence levels of the covariate, and when there are shared genetic/environmental causes of the covariate and trait of interest (Aschard et al., 2015; Hartwig et al., 2021). We hypothesized that adding binary treatment status as a second exposure in an MVMR analysis may adjust for medication use. This is as opposed to modelling treatment status as a covariate in SBP GWAS. We further hypothesized that inadequate medication adjustments to measured SBP values in treated individuals could underrepresent or mask differences in untreated SBP. This may potentially underestimate genetic variants’ effect estimates on SBP, and bias downstream MR causal estimates. Furthermore, this bias might be reinforced and difficult to detect in GWAS meta-analyses given that the direction of bias may be consistent across studies.

Here we examine the impact of a range of different methods for medication adjustment in SBP GWAS and downstream MR analyses in the UK Biobank. The objectives of our study were to: (1) compare genetic effect estimates and significance of genetic variant associations between SBP GWAS with various medication-adjustment methods, and (2) to investigate if there were differences between the different medication-adjustment methods for summary-level data MR estimation of (i) SBP on coronary artery disease (CAD), (ii) Body mass index (BMI) on SBP, and (iii) SBP on natural hair colour as a negative control outcome. The motivation of this work was to examine the importance of applying different medication-adjustment methods in primary or secondary sensitivity analyses when using GWAS summary statistics for downstream analyses. Downstream analyses include summary-level data MR, derivation of polygenic risk scores, heritability calculations and genetic correlation studies.

## 2. Methods

### 2.1. Data source and participants

Genetic and health data was sourced from the UK Biobank, which consists of approximately 500,000 participants aged between 38 to 73 years, and who were recruited between years 2006 and 2010 across the UK (Bycroft et al., 2018). At recruitment and upon consent, participants provide a range of information (physical measures, demographic measures, medication use etc.) via questionnaires and interviews with a trained nurse. More details about the UK Biobank have been described previously (Collins, 2012). UK Biobank received ethical approval from the Research Ethics Committee (REC reference: 11/NW/0382). This research has been conducted using the UK Biobank Resource under Application Number 81499. All analyses were in accordance with the Declaration of Helsinki.

In our study, we included participants who self-identified as "White British" (self-reported ethnicity, UK Biobank Field 21000) and who additionally fell within a genetically similar cluster defined by principal components of genotype data (Bycroft et al., 2018). We also further excluded individuals who did not have any reported SBP values. The final number of participants in our sample was 407,960. Further details on sample quality control (QC) are described in *Section 2.2. Data quality control*. We ascertained participants’ SBP by using readings which were obtained at the initial assessment visit. Final SBP values were determined in the following sequence based on data availability: the mean of two automatic measurements (data-field 4080_i0), the value of one automatic measurement if two automatic readings were absent, the mean of two manual measurements if automatic measurements were absent (data-field 93_i0), the value of one manual measurement if two manual readings were absent, otherwise missing.

### 2.2. Data quality control: sample, genotyping and imputation

We applied the following sample QC measures by applying filters to pre-existing indicators in the UK Biobank, excluding individuals with a mismatch between reported sex and genetically inferred sex, sex chromosome aneuploidy and outlying heterozygosity or genotype missingness rates (Mitchell et al., 2019).

UK Biobank Axiom Array genotype data was filtered to exclude variants with missingness > 0.015, minor allele frequency (MAF) < 0.01 and Hardy-Weinberg equilibrium (HWE) p-value < 0.0001, with linkage disequlibrium (LD) pruning to an r^2^ threshold of 0.1 using PLINKv2.00 (Chang et al., 2015). A total of 140,331 variants were used to construct a sparse genetic relationship matrix (GRM) with GCTA (Yang et al., 2011), which sets GRM off-diagonal elements that are below 0.05 to 0, to correct for sample relatedness in GWAS analyses. We used the ‘*pcapred*’ package (Lawson, 2022) to derive principal components by projection onto pre-computed UK Biobank loadings. These loadings were generated by flashPCA of UK Biobank genotype data and are distributed with the package; components projected in this way have been shown to correlate >0.99 with the principal components released by UK Biobank (data-field 22009) (Hu et al., 2025).. The first 10 principal components (explaining 18.3% of the variance) were deemed sufficient to capture the major axes of variation in population structure.

We used genotype data imputed with UK10K haplotype and 1000 Genomes (1000G) Phase 3 reference panels available in the UK Biobank. Further documentation on phasing and imputation of UK Biobank data is provided elsewhere (Marchini, 2015). We applied varying imputation quality (INFO) score filters depending on MAF of the genetic variant, as imputation INFO scores have less reliability when allele frequencies are rare. We excluded when any of the following criteria were met: MAF < 0.001; MAF <0.01 & INFO <0.8; MAF < 1 & INFO < 0.6 and variants with missingness rate > 0.10. The final number of variants (including SNPs and small insertions or deletions) was 14,507,256.

### 2.3. Adjusting for antihypertensive use

We compared seven different methods for adjusting for antihypertensive use in GWAS of SBP, including unadjusted analyses (Table 1). These were: (1) unadjusted, (2) add constant +10 mmHg, (3) add class-specific constant; (4) censored normal regression, (5) restrict to untreated individuals, (6) restrict to 38-49 years, (7) MVMR with binary treatment status as a second exposure. To further compare the effects of having varying proportions of the population who are treated, we also conducted GWAS in restricted age populations: 50-59 years and 60-73 years respectively, as these age groups have higher prevalence of antihypertensive use. In addition, we conducted a GWAS in treated individuals with the aim to estimate the effects of treatment by comparing effect estimates between treated and untreated GWAS. Adjustment methods (2) to (4) directly alter the measured SBP values in individual-level data; Methods (5) and (6) apply restrictions to individuals who are untreated or have a low proportion on treatment; Method (7) involves including genetic variants strongly associated with binary treatment status in MVMR as a second exposure. More detailed descriptions of the methods are provided in Table 1.

**Table 1.**
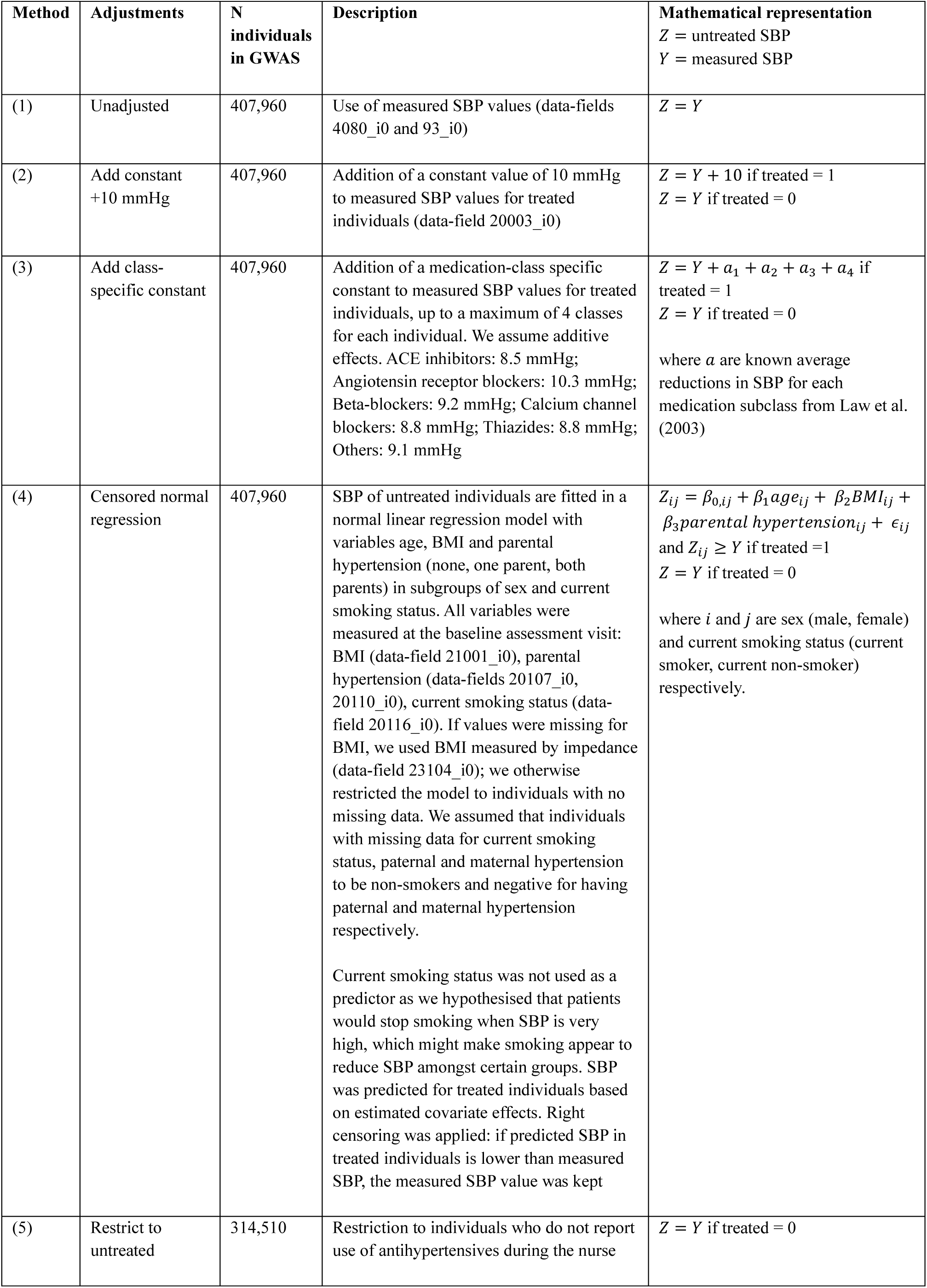
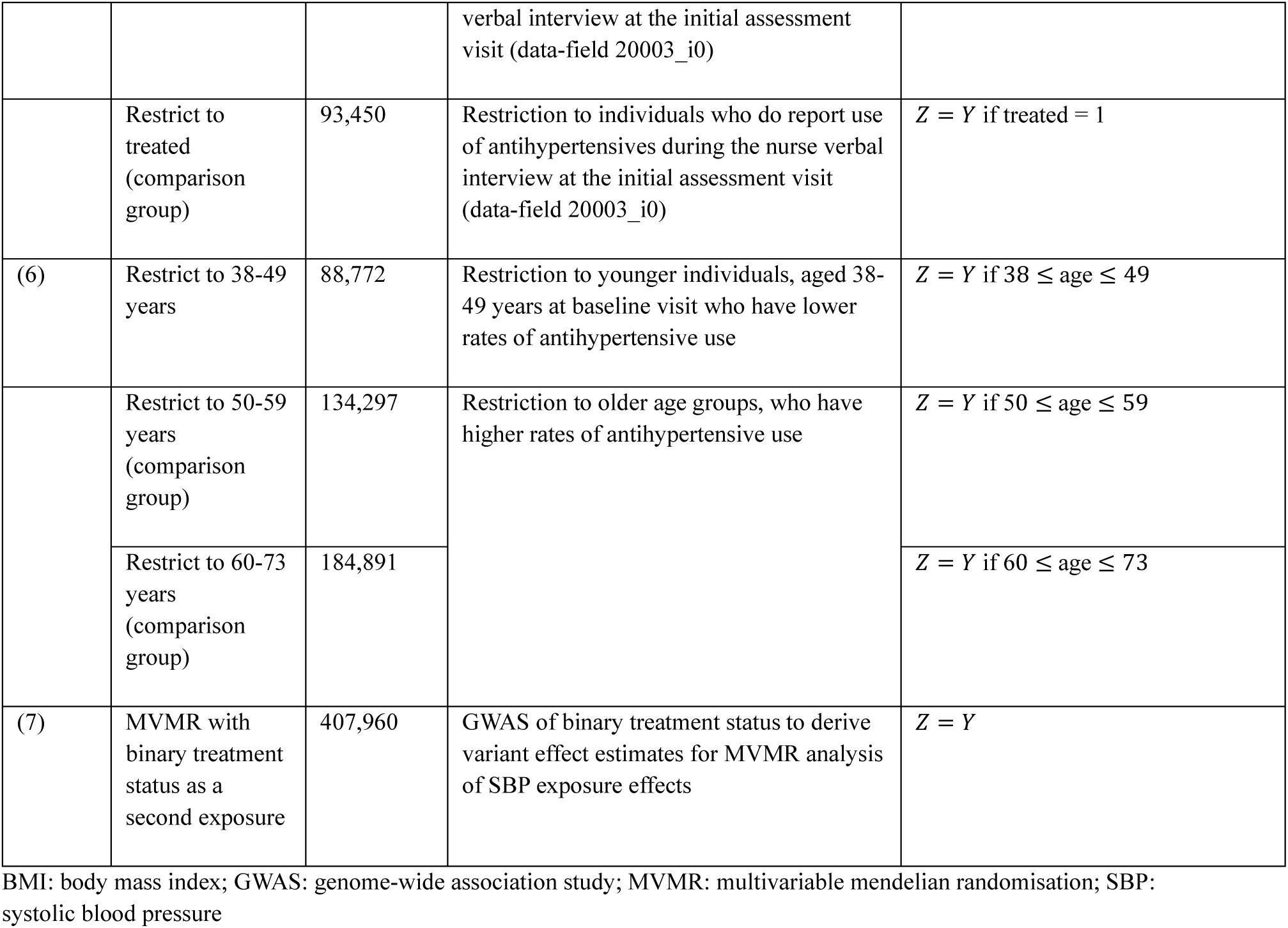
Antihypertensive adjustment methods applied in GWAS of systolic blood pressure.

We considered individuals to be treated if it was coded by the trained nurse during the verbal interview at the initial assessment visit (data-field 20003_i0). To determine type of antihypertensives used, we matched descriptions of active ingredients in data-field 20003_i0 to a list of standard medication names which was manually mapped in a previous study (Wu et al., 2019); The previous study did not include medications with less than ten counts in UK Biobank. We then matched standard medication names to antihypertensive British National Formulary (BNF) categories (See *Supplementary Table 1* for more information on the inclusion and exclusion criteria). These were then applied to Method (3), which added class-specific constants to participants who were treated. During this process, we found that 1,667 participants self-reported treatment in questionnaires but were not coded by the nurse and/or matched to active ingredients during the mapping process; we decided to consider them untreated.

For the addition of class-specific constants, we used mean values of SBP reduction reported from an analysis of 354 randomised trials, assuming an additive effect (Law, 2003). We decided to set a maximum limit of 4 classes as we wanted to minimise outliers in the SBP distributions. A low proportion of participants were found to be taking 5 or more classes of antihypertensives (n=675, 0.2%); for these participants, we included only the first four antihypertensives used by alphabetical order. Exploratory analyses showed no differences to the interquartile ranges when compared to not setting a limit. For the censored normal regression, we adapted the approach from a previous paper (Tobin et al., 2005), using risk factors for hypertension from the Framingham Heart Study (Parikh et al., 2008). We plotted the SBP distributions from various medication-adjustment methods, to check that the normality assumption of linearity would be met (*Supplementary Figure 1*).

### 2.4. Association analysis for systolic blood pressure

All GWAS were conducted using linear mixed models as implemented in fastGWA (Jiang et al., 2019) (with exception of binary treatment status trait, which used a generalised linear mixed model), with inclusion of age at baseline visit, sex, the first ten principal components and GRM as covariates. Binary treatment status GWAS included the same covariates, and was conducted to explore the utility of the MVMR approach for medication adjustment in MR analyses. Manhattan and Quantile-Quantile (QQ) plots are available in *Supplementary Figures 2-11*. We also generated Hudson plots using the ‘hudson’ package (Lucas et al., 2022): *Supplementary Figures 12-18* show a comparison of unadjusted with medication-adjusted GWAS; *Supplementary Figure 19* compares ‘add constant +10mmHg’ versus ‘add class-specific constant’; *Supplementary Figure 20* compares ‘restrict to untreated’ versus ‘restrict to treated’. We also conducted an LD score regression (LDSC) to ensure that we had adequate control of population stratification, and to calculate heritability (h^2^) estimates between different GWAS (Bulik-Sullivan et al., 2015). Full details of LDSC is available in Supplementary Text 1.

We conducted pair-wise comparisons of unclumped and clumped variants identified from both the unadjusted and adjusted GWAS. Clumping was conducted using the ‘ieugwasr’ package (via PLINKv1.90) (Hemani et al., 2024). Genome-wide significant variants (p < 5×10⁻⁸) were clumped using a window of 10,000 kb and an LD r² threshold of 0.001, with LD estimated from a European 1000 Genomes Phase 3 reference panel restricted to SNPs with MAF > 0.01. The reference panel comprised SNPs only and did not include indels. These clumped variants are henceforth referred to as ‘independent genetic variants’. Venn diagrams comparing the extent of overlap of unclumped genome-wide significant variants are available in *Supplementary Figures 21-22*.

The extent of overlap of unclumped variants between GWAS is influenced by the LD structure of genome-wide significant variants and by GWAS sample size, which affects power to detect associations exceeding the genome-wide significance threshold (P < 5 × 10⁻⁸). To account for this, all further analyses were restricted to independent genetic variants. We compared the estimated effect and significance level of independent genetic variants for SBP identified between unadjusted and medication-adjusted GWAS. Unadjusted effect estimates were regressed against medication-adjusted estimates to obtain the regression slope, and a two-tailed T-test was used to assess whether this slope differed significantly from 1 (indicating agreement between models), as shown below:

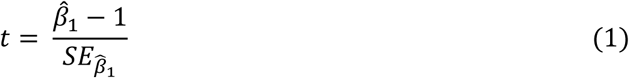

Where 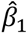 is the estimated regression slope and 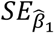 is the standard error of the regression slope estimate.

We also conducted a test for heterogeneity estimated by the Cochran’s Q statistic; This is represented in the equation below:

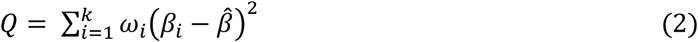

Where *k* is the number of effect estimates to be compared (we considered pair-wise comparisons), *ω*_i_ is the inverse variance weight of *β*_i_, *β*_i_ is a vector of beta values or effect estimates from unadjusted and adjusted GWAS respectively, 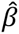 is the inverse-variance weighted mean estimate across *β*_i_, given by:

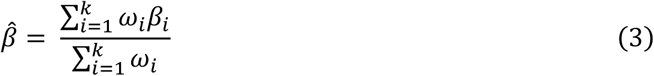

Where *ω*_i_ is:

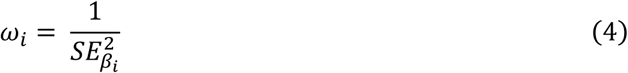

where 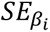 is the standard error of *β*_i_.

We used the Holm-Bonferroni method to correct for multiple testing (Holm, 1979). We considered that there was evidence of heterogeneity if Bonferroni p-values were less than 0.05.

We also tabulated a list of independent genetic variants which were genome-wide significant in direct medication-adjustment methods but not in the unadjusted GWAS. We include further details for these variants, such as whether they were novel with respect to unadjusted analyses, and whether they had been previously published in literature (Hemani et al., 2024; Magno & Maia, 2020). Further details are provided in *Supplementary Text 2*.

### 2.5. GWAS for coronary artery disease, body mass index and natural hair colour

We chose GWAS for CAD and BMI which did not contain UK Biobank participants to avoid sample overlap. The CAD GWAS by the CARDIoGRAMplusC4D Consortium was based on a meta-analysis of 184,305 cases and controls pooled from 48 separate studies, of which 77% were of European descent (Nikpay et al., 2015). As the summary statistics of the participants of European ancestry were available, we used this subset in our MR analyses. The BMI GWAS was based on a meta-analysis of 339,224 individuals pooled from 125 studies, of which 95% of individuals were of European descent (Locke et al., 2015).

The natural hair colour GWAS (for negative control outcome) was conducted in the UK Biobank with 463,005 individuals (Sanderson, Richardson, et al., 2021). Although this originates from the same sample as our cohort, we anticipate any overfitting due to sample overlap to bias estimates away from the null. This is less of a concern because the goal of the negative control outcome is not to estimate causal effects but detect potential bias arising from population stratification. If we observed negative control effect estimates that span the null, we would be more confident that our analyses were less likely to be biased by population stratification.

### 2.6. Summary-level MR analyses

We extracted independent genetic variants to instrument exposures in MR analyses by using LD-clumped variants. Information on how LD-clumping was conducted is provided above. We conducted summary-level MR analyses to investigate the effect of (i) SBP on CAD, (ii) BMI on SBP, (iii) SBP on natural hair colour, using GWAS for SBP with each of the seven proposed medication adjustments. We used the ‘TwoSampleMR’ and ‘MVMR’ packages to conduct the univariable and multivariable MR analyses respectively (Hemani et al., 2017, 2018; Sanderson, Spiller, et al., 2021). For data harmonisation, we used allele frequency to infer forward strand alleles; palindromic SNPs with MAF above 0.42 were discarded. Our primary MR method was inverse variance weighting (IVW); we also reported MR Egger, weighted median and weighted mode estimates in *Supplementary Tables 3-5* as sensitivity analyses. Odds ratios per standard deviation increase in SBP were reported for SBP-CAD analyses.

### 2.7. Assumptions of MR and MVMR

The three core IV assumptions for MR are: (1) relevance: SNPs are robustly associated with the exposure; (2) exchangeability: there are no confounders for the SNPs-outcome relationship; (3) exclusion restriction: SNPs do not have an independent effect on the outcome other than through the exposure (Sanderson et al., 2022). MVMR shares the same core assumptions with a modification to the first assumption: since MVMR has two or more exposures, SNPs should be robustly associated with each exposure conditional on the other exposure(s) included. To check the robustness of our assumptions for MVMR, we calculated the conditional F-statistic to test our instrument strength. A conditional F-statistic greater than 10 means we are able to reject the a null hypothesis that the instruments weakly predict the exposure (Sanderson, Spiller, et al., 2021).

### 2.8. Code availability

Data preparation and GWAS analyses were conducted on the UK Biobank Research Analysis Platform. All MR analyses were conducted in R version 4.4.1. All codes are available online on GitHub: https://github.com/aaronjyyap/med-adjusted-GWAS-MR.

## 3. Results

### 3.1. Characteristics of cohort

Table 2 shows the characteristics of the White British cohort (N=407,960) included in our study. The median age at baseline was 58 years. 22.9% (N=93,450) of participants were recorded to be on antihypertensive therapy; this differed by age group: 6.8% for 38-49 years, 18.1% for 50-59 years, 34.1% for 60-73 years. Median measured SBP also increases with age group (38-49 years: 128.0mmHg; 50-59 years: 135.0mmHg; 60-73 years: 142.5mmHg). In treated individuals, the measured SBP is slightly higher than in the untreated (SBP, median = treated: 143.0mmHg; untreated: 135.0mmHg). Applying direct adjustments onto SBP values using Methods (2) to (4) resulted in treated having higher adjusted SBP values – the median SBP values were 153.0, 158.5, 148.5 for the ‘add constant +10 mmHg’, ‘add class-specific constant’ and ‘censored normal regression’ methods for medication adjustment respectively. The correlation for measured SBP values and treatment was low (r=0.164).

**Table 2.**
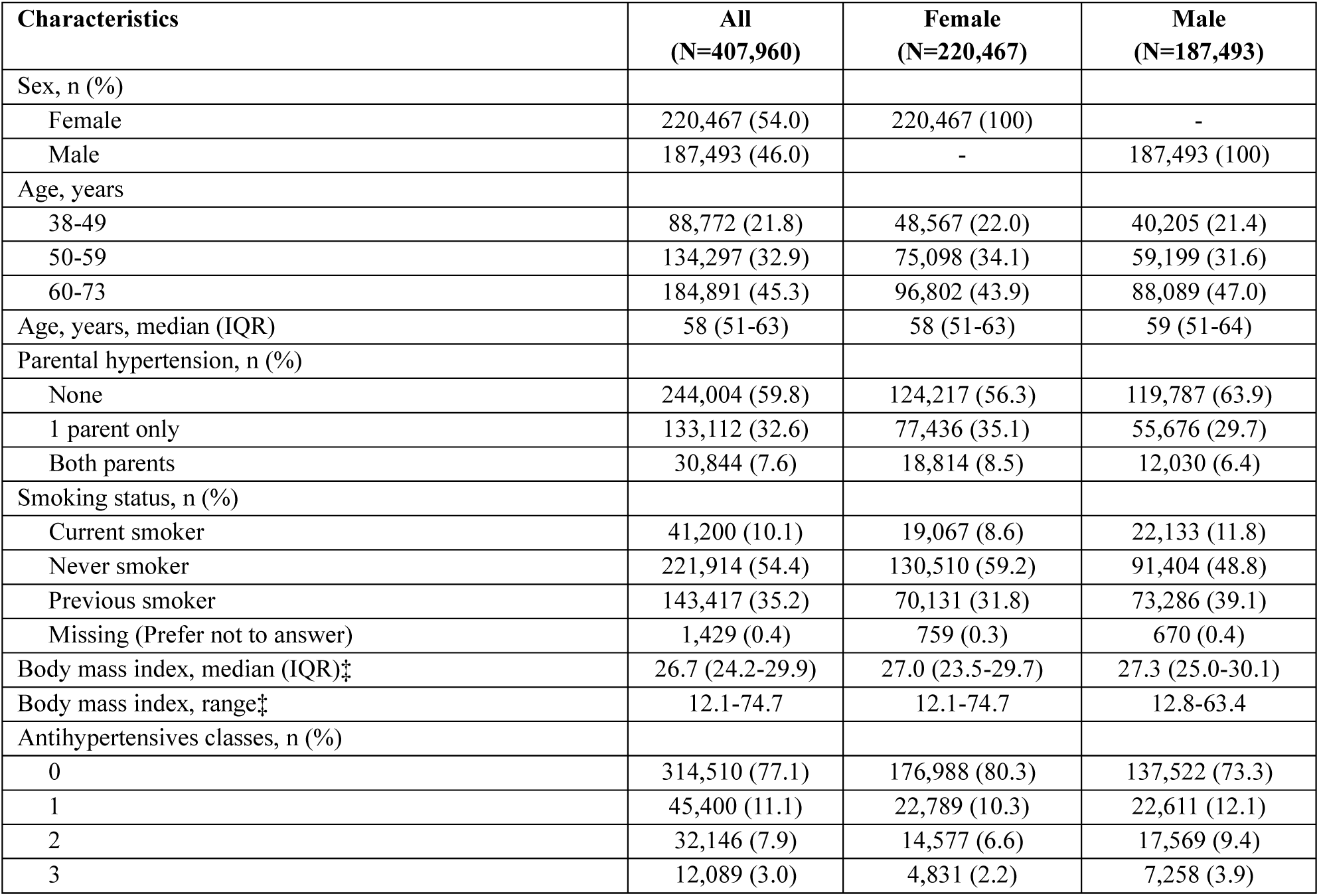
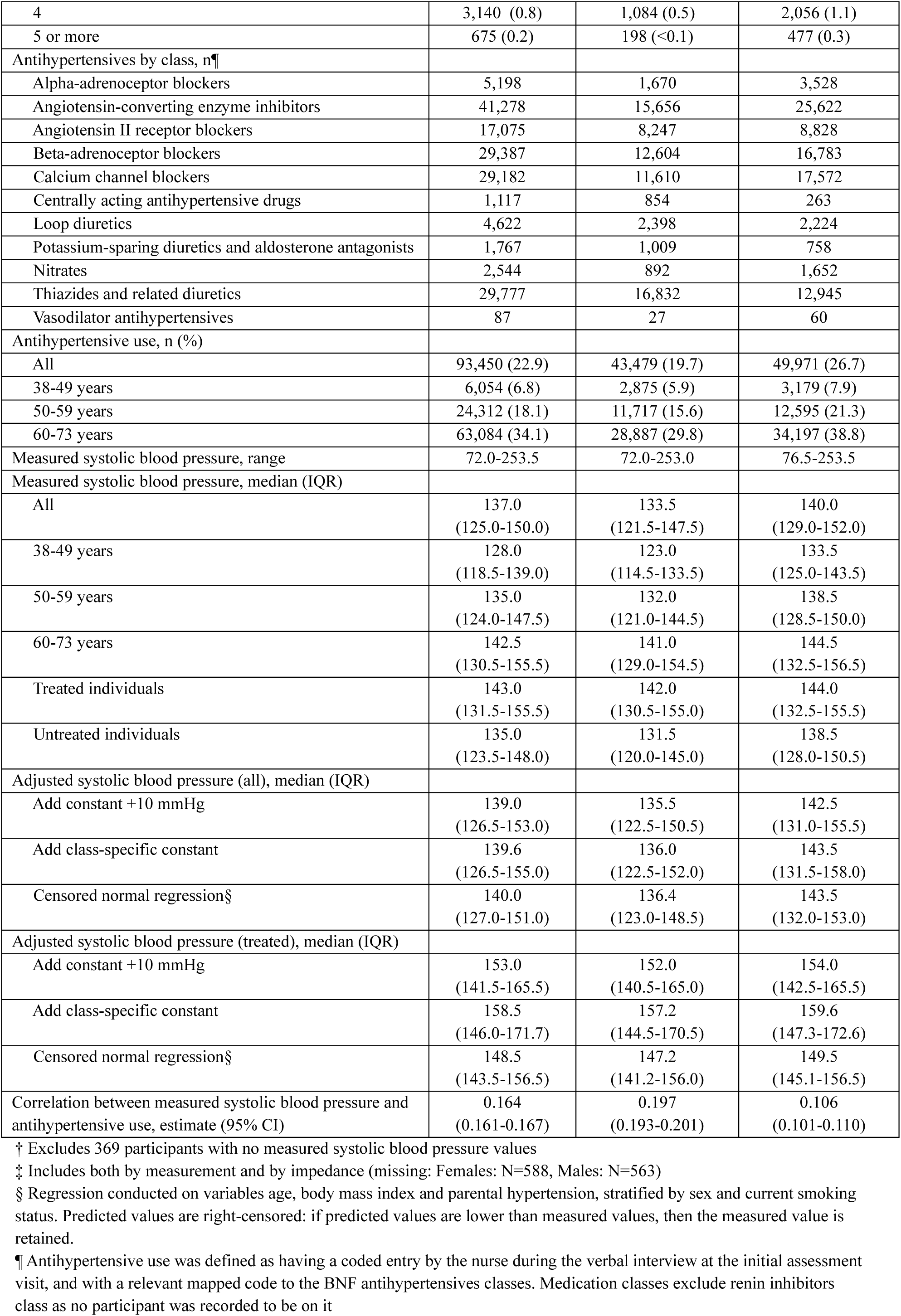
Characteristics of White British cohort in UK Biobank†.

| Characteristics | All<br>(N=407,960) | Female<br>(N=220,467) | Male<br>(N=187,493) |
| --- | --- | --- | --- |
| Sex, n (%) |  |  |  |
| Female | 220,467 (54.0) | 220,467 (100) | - |
| Male | 187,493 (46.0) | - | 187,493 (100) |
| Age, years |  |  |  |
| 38-49 | 88,772 (21.8) | 48,567 (22.0) | 40,205 (21.4) |
| 50-59 | 134,297 (32.9) | 75,098 (34.1) | 59,199 (31.6) |
| 60-73 | 184,891 (45.3) | 96,802 (43.9) | 88,089 (47.0) |
| Age, years, median (IQR) | 58 (51-63) | 58 (51-63) | 59 (51-64) |
| Parental hypertension, n (%) |  |  |  |
| None | 244,004 (59.8) | 124,217 (56.3) | 119,787 (63.9) |
| 1 parent only | 133,112 (32.6) | 77,436 (35.1) | 55,676 (29.7) |
| Both parents | 30,844 (7.6) | 18,814 (8.5) | 12,030 (6.4) |
| Smoking status, n (%) |  |  |  |
| Current smoker | 41,200 (10.1) | 19,067 (8.6) | 22,133 (11.8) |
| Never smoker | 221,914 (54.4) | 130,510 (59.2) | 91,404 (48.8) |
| Previous smoker | 143,417 (35.2) | 70,131 (31.8) | 73,286 (39.1) |
| Missing (Prefer not to answer) | 1,429 (0.4) | 759 (0.3) | 670 (0.4) |
| Body mass index, median (IQR)‡ | 26.7 (24.2-29.9) | 27.0 (23.5-29.7) | 27.3 (25.0-30.1) |
| Body mass index, range‡ | 12.1-74.7 | 12.1-74.7 | 12.8-63.4 |
| Antihypertensives classes, n (%) |  |  |  |
| 0 | 314,510 (77.1) | 176,988 (80.3) | 137,522 (73.3) |
| 1 | 45,400 (11.1) | 22,789 (10.3) | 22,611 (12.1) |
| 2 | 32,146 (7.9) | 14,577 (6.6) | 17,569 (9.4) |
| 3 | 12,089 (3.0) | 4,831 (2.2) | 7,258 (3.9) |
| 4 | 3,140 (0.8) | 1,084 (0.5) | 2,056 (1.1) |
| 5 or more | 675 (0.2) | 198 (<0.1) | 477 (0.3) |
| Antihypertensives by class, n¶ |  |  |  |
| Alpha-adrenoceptor blockers | 5,198 | 1,670 | 3,528 |
| Angiotensin-converting enzyme inhibitors | 41,278 | 15,656 | 25,622 |
| Angiotensin II receptor blockers | 17,075 | 8,247 | 8,828 |
| Beta-adrenoceptor blockers | 29,387 | 12,604 | 16,783 |
| Calcium channel blockers | 29,182 | 11,610 | 17,572 |
| Centrally acting antihypertensive drugs | 1,117 | 854 | 263 |
| Loop diuretics | 4,622 | 2,398 | 2,224 |
| Potassium-sparing diuretics and aldosterone antagonists | 1,767 | 1,009 | 758 |
| Nitrates | 2,544 | 892 | 1,652 |
| Thiazides and related diuretics | 29,777 | 16,832 | 12,945 |
| Vasodilator antihypertensives | 87 | 27 | 60 |
| Antihypertensive use, n (%) |  |  |  |
| All | 93,450 (22.9) | 43,479 (19.7) | 49,971 (26.7) |
| 38-49 years | 6,054 (6.8) | 2,875 (5.9) | 3,179 (7.9) |
| 50-59 years | 24,312 (18.1) | 11,717 (15.6) | 12,595 (21.3) |
| 60-73 years | 63,084 (34.1) | 28,887 (29.8) | 34,197 (38.8) |
| Measured systolic blood pressure, range | 72.0-253.5 | 72.0-253.0 | 76.5-253.5 |
| Measured systolic blood pressure, median (IQR) |  |  |  |
| All | 137.0<br>(125.0-150.0) | 133.5<br>(121.5-147.5) | 140.0<br>(129.0-152.0) |
| 38-49 years | 128.0<br>(118.5-139.0) | 123.0<br>(114.5-133.5) | 133.5<br>(125.0-143.5) |
| 50-59 years | 135.0<br>(124.0-147.5) | 132.0<br>(121.0-144.5) | 138.5<br>(128.5-150.0) |
| 60-73 years | 142.5<br>(130.5-155.5) | 141.0<br>(129.0-154.5) | 144.5<br>(132.5-156.5) |
| Treated individuals | 143.0<br>(131.5-155.5) | 142.0<br>(130.5-155.0) | 144.0<br>(132.5-155.5) |
| Untreated individuals | 135.0<br>(123.5-148.0) | 131.5<br>(120.0-145.0) | 138.5<br>(128.0-150.5) |
| Adjusted systolic blood pressure (all), median (IQR) |  |  |  |
| Add constant +10 mmHg | 139.0<br>(126.5-153.0) | 135.5<br>(122.5-150.5) | 142.5<br>(131.0-155.5) |
| Add class-specific constant | 139.6<br>(126.5-155.0) | 136.0<br>(122.5-152.0) | 143.5<br>(131.5-158.0) |
| Censored normal regression§ | 140.0<br>(127.0-151.0) | 136.4<br>(123.0-148.5) | 143.5<br>(132.0-153.0) |
| Adjusted systolic blood pressure (treated), median (IQR) |  |  |  |
| Add constant +10 mmHg | 153.0<br>(141.5-165.5) | 152.0<br>(140.5-165.0) | 154.0<br>(142.5-165.5) |
| Add class-specific constant | 158.5<br>(146.0-171.7) | 157.2<br>(144.5-170.5) | 159.6<br>(147.3-172.6) |
| Censored normal regression§ | 148.5<br>(143.5-156.5) | 147.2<br>(141.2-156.0) | 149.5<br>(145.1-156.5) |
| Correlation between measured systolic blood pressure and antihypertensive use, estimate (95% CI) | 0.164<br>(0.161-0.167) | 0.197<br>(0.193-0.201) | 0.106<br>(0.101-0.110) |

† Excludes 369 participants with no measured systolic blood pressure values
‡ Includes both by measurement and by impedance (missing: Females: N=588, Males: N=563)
§ Regression conducted on variables age, body mass index and parental hypertension, stratified by sex and current smoking status. Predicted values are right-censored: if predicted values are lower than measured values, then the measured value is retained.
¶ Antihypertensive use was defined as having a coded entry by the nurse during the verbal interview at the initial assessment visit, and with a relevant mapped code to the BNF antihypertensives classes. Medication classes exclude renin inhibitors class as no participant was recorded to be on it

### 3.2. Comparing SBP associations from medication adjusted GWAS methods

Figure 1 compares the significance of independent genetic variants from unadjusted and medication-adjusted GWAS. Both the unadjusted and direct adjustment methods identified largely overlapping genome-wide significant independent genetic variants, although direct adjustment methods generally yielded more significant p-values. Furthermore, direct adjustment methods were able to detect additional independent genetic variants. Adding class-specific constants yielded the highest number of additional independent genetic variants not detected in the unadjusted GWAS (n=149), followed by adding constant +10mmHg (n=84) and censored normal regression (n=67). The list of additional variants are in *Supplementary Table 7; also see Section 3.6*.

**Figure 1.**
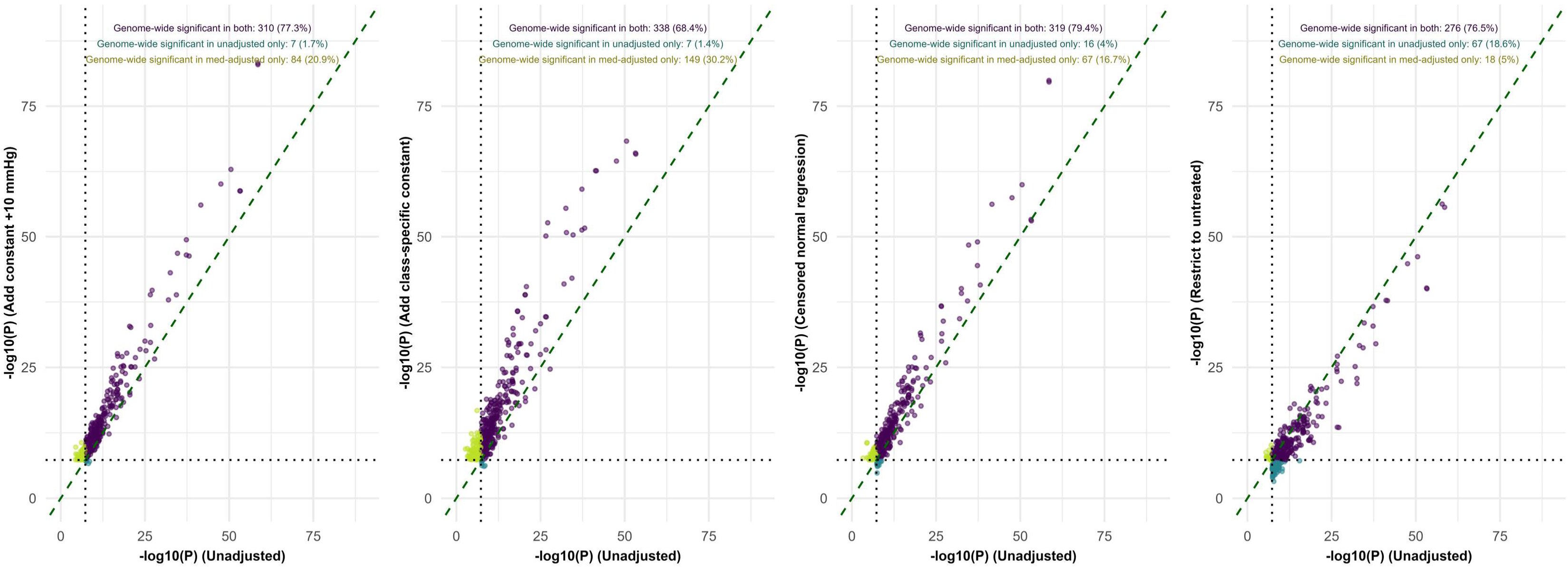
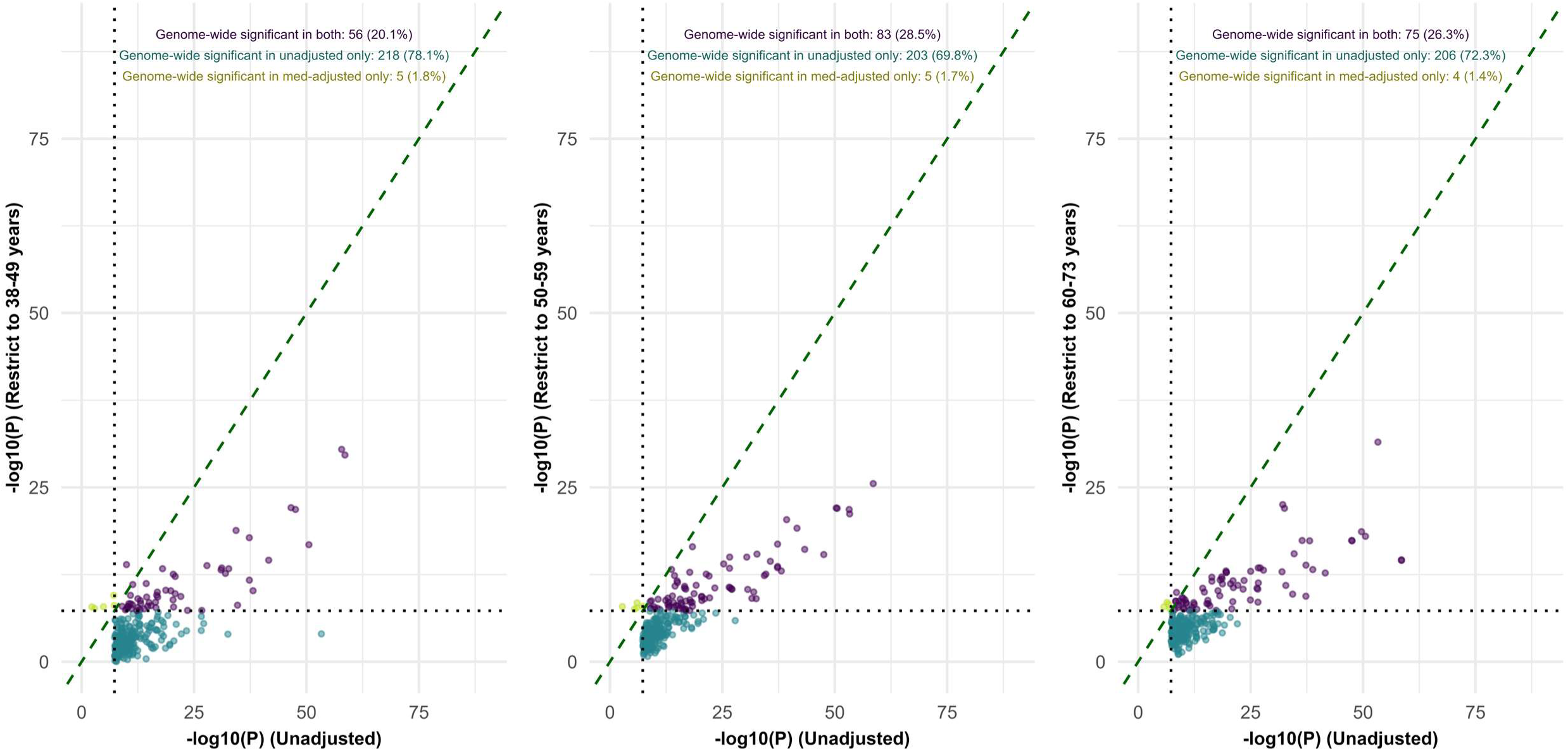
Scatter plot of P-values of independent genetic variants from unadjusted and medication-adjusted GWAS. GWAS: genome-wide association study; P: p-value The x-axis and y-axis shows the p-values in the unadjusted and adjusted GWAS respectively. Each dot represents an independent genetic variant from the unadjusted or adjusted GWAS. Purple dots indicate variants which are genome-wide significant in both unadjusted and adjusted GWAS; Dark green dots show variants which are genome-wide significant in the unadjusted GWAS but not the adjusted GWAS; Light green dots show variants which are genome-wide significant in the adjusted GWAS but not the unadjusted GWAS. The dotted diagonal green line has a gradient of 1; if p-values from both GWAS are similar, we would expect to observe dots along the green line. A steeper slope greater than 1 indicates that the adjusted GWAS has a more significant p-value than the unadjusted GWAS, and vice-versa.

Conversely, restriction-based adjustment methods displayed a trend of having less significant p-values, with more independent genetic variants not surpassing the genome-wide significance threshold than in the unadjusted analysis, as was most apparent in stratified age group GWAS.

Figure 2 shows the effect estimates of independent genetic variants from the unadjusted versus medication-adjusted GWAS. We observe that effect estimates tend to be increased when medication adjustment is applied to SBP values as compared to the unadjusted GWAS. Differences were largest when adding class-specific constants (slope=1.28, p=1.06×10^-35^), followed by adding constant +10mmHg (slope=1.17, p=4.52×10^-36^). There was evidence of significant heterogeneity in effect estimates for some variants when comparing the unadjusted and adding class-specific constants GWAS (rs1275979 [*KCNK3*], rs16998073 [*FGF5*], rs13219548 [intergenic]) and the GWAS restricted to 38-49 years (rs2392929 [*RP5-884M6.1*]). Further information on these variants are provided in Table 3.

**Figure 2.**
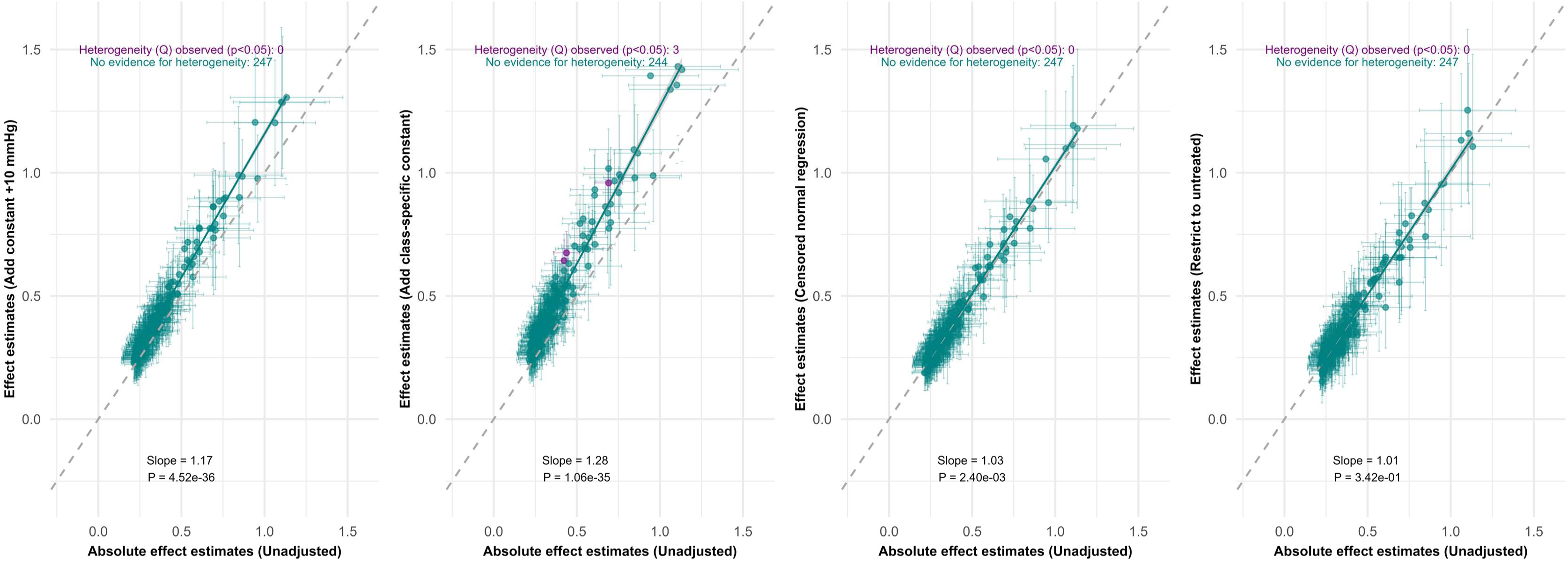
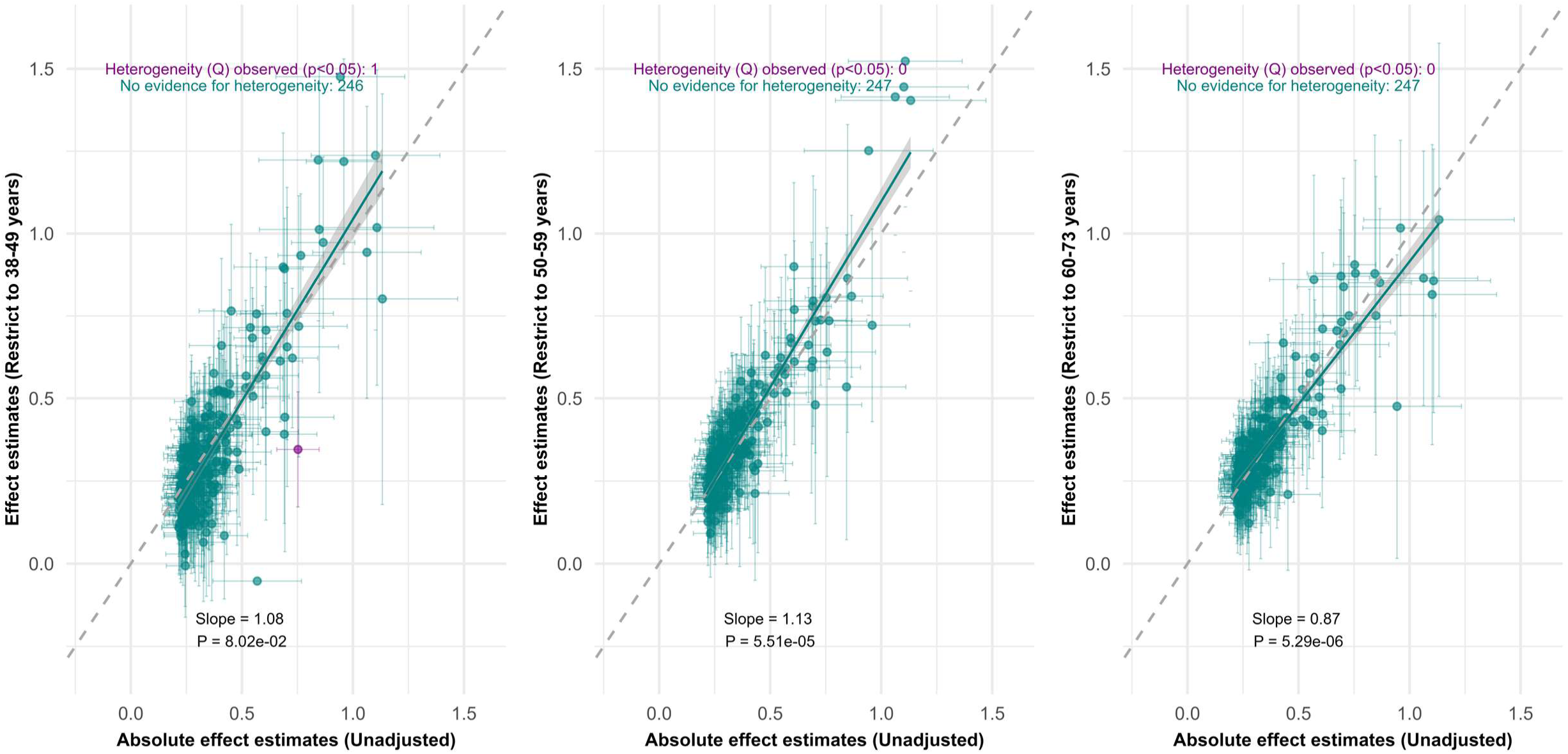
Scatter plot of effect estimates of independent genetic variants from unadjusted GWAS (reference) versus adjusted GWAS. GWAS: genome-wide association study; P: p-value The x-axis shows the absolute effect estimates of independent genetic variants from unadjusted GWAS. The y-axis shows the corresponding effect estimates of these variants in the adjusted GWAS. Each dot represents a single genetic variant with its corresponding standard error intervals. Green dots indicate that there is no evidence for heterogeneity of effect estimates for genetic variants. Purple dots indicate evidence (p<0.05) of heterogeneity between effect estimates for the same genetic variant in both GWAS after correction for multiple testing. The grey shaded region shows the 95% confidence intervals around the fitted regression line. The dotted green line has a gradient of 1; if effect estimates from both GWAS are similar, we will observe dots close to the green line. A positive slope >1 indicates stronger absolute effect estimates in the adjusted GWAS, a positive slope <1 indicates stronger absolute effect estimated in the unadjusted GWAS. The reported p-values are from a two-tailed t-test evaluating if the slope term differs from 1.

**Table 3.** Genetic variants with heterogeneous effect estimates in the unadjusted and medication-adjusted GWAS†.

| rsID | Reference GWAS |  |  | Comparison GWAS |  |  | Chr | Position<br>(GRCh37/hg 19) | Nearest gene | Gene context | Gene function |
| --- | --- | --- | --- | --- | --- | --- | --- | --- | --- | --- | --- |
|  | Name | Beta | Standard Error | Name | Beta | Standard Error |  |  |  |  |  |
| Unadjusted versus medication-adjusted GWAS |  |  |  |  |  |  |  |  |  |  |  |
| rs1275979 | Unadjusted | -0.438 | 0.040 | Add class-specific constant | -0.675 | 0.044 | 2 | 26,923,568 | KCNK3 | Intron variant | Encodes potassium channel proteins |
| rs16998073 | Unadjusted | 0.692 | 0.043 | Add class-specific constant | 0.959 | 0.047 | 4 | 81,184,341 | FGF5 | Upstream gene variant | Regulates cell growth, tissue repair, tumour and hair growth |
| rs13219548 | Unadjusted | 0.423 | 0.039 | Add class-specific constant | 0.643 | 0.043 | 6 | 127,165,290 | - | Intergenic variant | - |
| rs2392929 | Unadjusted | 0.753 | 0.049 | Restrict to 38-49 years | 0.345 | 0.089 | 7 | 106,414,069 | RP5-884M6.1 | Upstream gene variant | No established function |
| rs12509595 | Add class-specific constant | 0.964 | 0.047 | Unadjusted | 0.694 | 0.043 | 4 | 81,182,554 | - | Intergenic variant | - |
| rs13112725 | Restrict to 38-49 years | 0.640 | 0.083 | Unadjusted | 0.293 | 0.045 | 4 | 106,911,742 | NPNT | Intron variant | Encodes protein involved in renal development, cell adhesion, spreading and survival mediated by integrin |
| rs196562 | Restrict to 38-49 years | -0.640 | 0.113 | Unadjusted | -0.172 | 0.062 | 7 | 36,298,800 | EEPD1 | Intron variant | Enables DNA binding and catalytic activity, involves in positive regulation of cholesterol efflux |
| rs113475805 | Restrict to 38-49 years | -0.436 | 0.078 | Unadjusted | -0.137 | 0.043 | 12 | 13,853,938 | GRIN2B | Intron variant | Involved in brain development, synaptic transmission, plasticity, learning, memory |
| rs11685990 | Restrict to 50-59 years | -0.411 | 0.072 | Unadjusted | -0.131 | 0.042 | 2 | 56,381,388 | RP11-481J13.1 | Intron variant | No established function |
| Untreated versus treated GWAS |  |  |  |  |  |  |  |  |  |  |  |
| rs731024 | Restrict to untreated | -0.302 | 0.046 | Restrict to treated | 0.128 | 0.090 | 1 | 6,644,723 | ZBTB48 | Intron variant | Maintain telomere stability, control transcription of genes by binding to regulatory regions |
| rs861583 | Restrict to untreated | 0.277 | 0.045 | Restrict to treated | -0.096 | 0.086 | 1 | 184,734,785 | - | Intergenic variant | - |
| rs10032845 | Restrict to untreated | 0.298 | 0.045 | Restrict to treated | -0.061 | 0.085 | 4 | 26,801,574 | - | Intergenic variant | - |
| rs11099097 | Restrict to untreated | 0.767 | 0.048 | Restrict to treated | 0.240 | 0.091 | 4 | 81,167,309 | - | Intergenic variant | - |
| rs877116 | Restrict to untreated | -0.427 | 0.044 | Restrict to treated | -0.025 | 0.085 | 8 | 10,712,945 | - | Intergenic variant | - |
| rs73563812 | Restrict to untreated | -0.439 | 0.051 | Restrict to treated | -0.019 | 0.099 | 8 | 25,900,405 | *EBF2* | Intron variant | Controls expression of genes involved in developmental processes |
| rs6990308 | Restrict to untreated | 0.299 | 0.050 | Restrict to treated | -0.140 | 0.095 | 8 | 65,462,235 | *RP11-21C4.4* | Downstream gene variant | No established function |
| rs12258967 | Restrict to untreated | -0.605 | 0.047 | Restrict to treated | -0.203 | 0.092 | 10 | 18,727,959 | *CACNB2* | Intron variant | Modulates calcium influx into cells. Also has been linked to Lambert-Eaton myasthenic syndrome and Brugada syndrome |
| rs7070797 | Restrict to untreated | -0.585 | 0.062 | Restrict to treated | -0.067 | 0.124 | 10 | 63,551,773 | *RP11-491H19.1* | Intron variant | No established function |
| rs1801253 | Restrict to untreated | 0.439 | 0.050 | Restrict to treated | -0.131 | 0.097 | 10 | 115,805,056 | *ADRB1* | Missense variant | Mediates epinephrine and norepinephrine through beta-1 adrenoceptors in the heart |
| rs3736290 | Restrict to untreated | 0.379 | 0.044 | Restrict to treated | -0.043 | 0.085 | 15 | 40,321,351 | *EIF2AK4* | Intron variant | Downregulates protein synthesis during amino acid deprivation |
| rs4886619 | Restrict to untreated | -0.466 | 0.049 | Restrict to treated | -0.043 | 0.094 | 15 | 75,106,138 | *LMAN1L* | Intron variant | Transport glycoproteins from endoplasmic reticulum to ER-Golgi intermediate compartment |
| rs11102485 | Restrict to treated | 0.889 | 0.152 | Restrict to untreated | 0.405 | 0.082 | 1 | 113,052,923 | *WNT2B* | Intron variant | Regulates embryonic development, stem cell maintenance, tissue homeostasis |
| rs7913920 | Restrict to treated | -0.543 | 0.089 | Restrict to untreated | -0.181 | 0.046 | 10 | 104,923,628 | *NT5C2* | Intron variant | Regulate nucleoside pools in cytosol through cellular purine metabolism |

Chr: chromosome; GWAS: genome-wide association study
† Gene information from Ensembl VEP (GRCh37/hg19)

We observed a similar trend when we compared independent genetic variants in the medication-adjusted GWAS (note: clumping conducted in medication-adjusted instead of unadjusted GWAS). (*Supplementary Figure 25*). Effect estimates were deflated in the unadjusted GWAS relative to medication-adjusted GWAS, with adding class-specific constants (slope=0.76, p=2.77×10^-84^) displaying the greatest difference. In addition, we observe an attenuation of effect estimates with increasing age groups. We also found that there were several genetic variants which had stronger effect estimates in the medication-adjusted GWAS (Table 3). These were: rs12509595 [intergenic], rs13112725 [*NPNT*], rs196562 [*EEPD1*], rs113475805 [*GRIN2B*] and rs11685990 [*RP11-481J13.1*].

### 3.3. Comparing add constant +10mmHg with add class-specific constant

We also compared the influence of adding constant +10mmHg to measured SBP values in treated individuals with adding class-specific constant on SBP GWAS results (*Supplementary Figures 23, 26, 27*). 4 (0.8%) independent genetic variants were genome-wide significant in the adding constant +10mmHg but not in adding class-specific constant GWAS. In comparison, there were more independent genetic variants (n=63, 13.2%) that were genome-wide significant in the adding class-specific constant GWAS. Effect estimates were on average slightly higher in magnitude in the adding class-specific constant medication-adjustment method with no evidence for heterogeneity after correction for multiple testing.

### 3.4. Comparing effect estimates of untreated versus treated GWAS

When we compared independent genetic variants that reached genome-wide significance in the SBP GWAS restricted to untreated (with no modification to measured SBP values) with the treated, we observed that effect estimates were deflated in the GWAS of treated individuals (slope = 0.57, p=1.31×10^-10^) (Figure 3). As a result of the deflated effect estimates as well as reduced sample size, there were fewer independent genetic variants detected in the treated GWAS (n=10) (*Supplementary Figure 28*). More details of genetic variants with heterogeneous effect estimates between untreated and treated GWAS can be found in Table 3. A scatter plot of p-values of untreated versus treated GWAS can be found in *Supplementary Figure 24*. We found no overlap between heterogeneous variants in unadjusted versus medication-adjusted GWAS, and heterogeneous variants in untreated versus treated GWAS.

**Figure 3.**
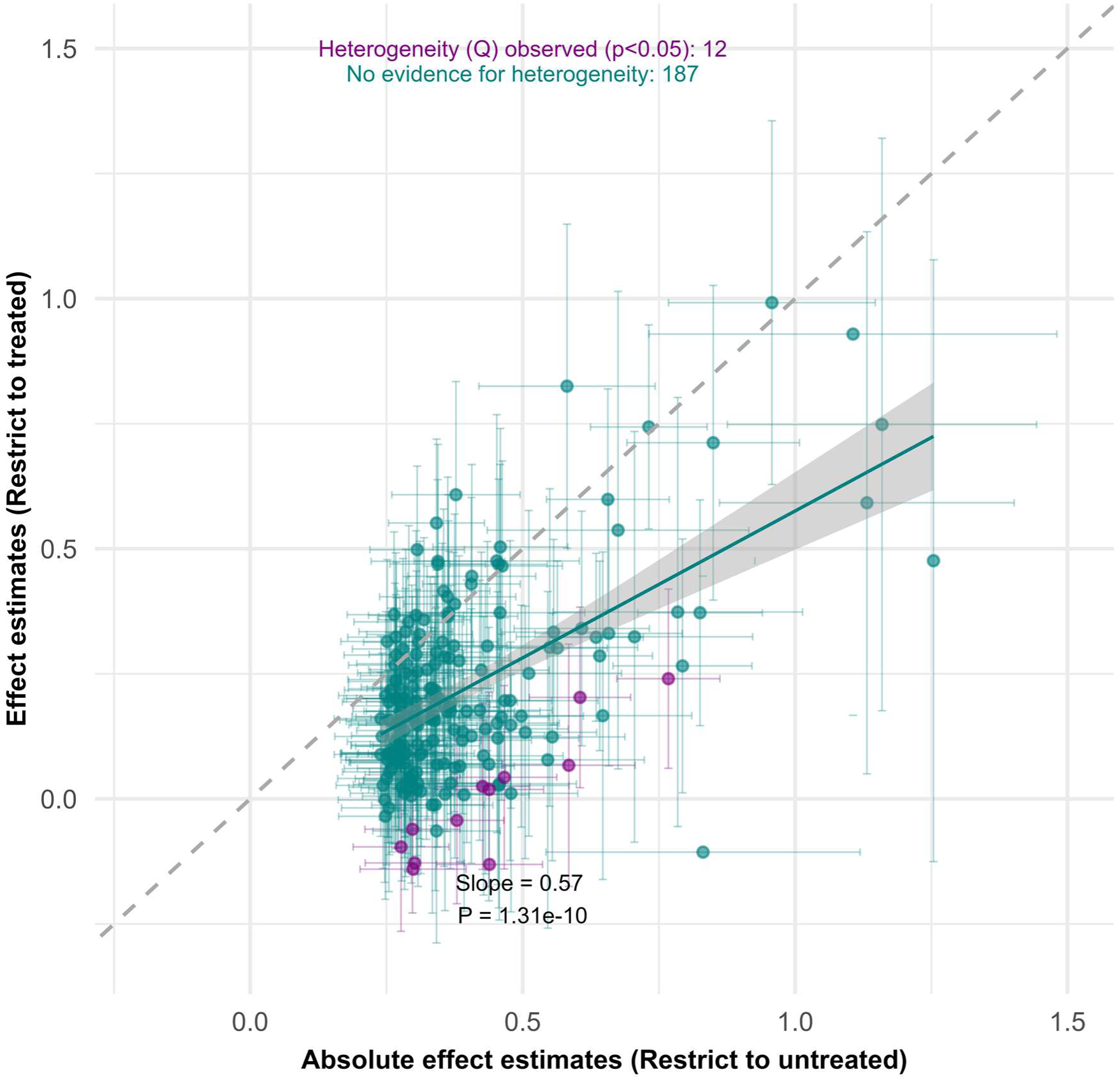
Scatter plot of independent genetic variants effect estimates of untreated versus treated GWAS. GWAS: genome-wide association study; P: p-value The x-axis shows the absolute effect estimates of independent genetic variants from GWAS restricted to untreated individuals. The y-axis shows the corresponding effect estimates of these variants in the GWAS restricted to treated individuals. Each dot represents a single genetic variant with its corresponding standard error intervals. Green dots indicate that there is no evidence for heterogeneity of effect estimates for genetic variants. Purple dots indicate evidence (p<0.05) of heterogeneity between effect estimates for the same genetic variant in both GWAS after correction for multiple testing. The grey shaded region shows the 95% confidence intervals around the fitted regression line. The dotted green line has a gradient of 1; if effect estimates from both GWAS are similar, we will observe dots close to the green line. A positive slope <1 indicates stronger absolute effect estimated in the GWAS restricted to untreated individuals. The reported p-values are from a two-tailed t-test evaluating if the slope term differs from 1.

### 3.5. SNP-based heritability

We observed that different medication-adjustment methods had different SNP-based h^2^ estimates (*Supplementary Table 2*). Adding class-specific constants and restricting to 38-49 years had the highest SNP-based h² values (0.182 and 0.188 respectively) as compared to unadjusted (0.132). Adding constant +10 mmHg yielded a SNP-based h² value of 0.162.

### 3.6. Novel independent genetic variants from medication-adjusted GWAS

The full list of independent genetic variants which were detected in direct medication-adjusted GWAS but not in the unadjusted GWAS can be found in *Supplementary Table 7*. We found that adding constant +10 mmHg, adding class-specific constant, and censored normal regression identified 12, 31 and 16 independent genetic variants respectively which had not been previously published in literature. These included genes known to have cardiometabolic functions (*CDKAL1*, *RORA*, *SLC39A14*, *PALLD*, *HSD17B12*, *PKHD1*), although there were also genes with little known SBP connection.

### 3.7. MR findings

Figure 4 show forest plots for summary-level data MR IVW estimates for the causal effect of (1) SBP on CAD, (2) BMI on SBP, and (3) SBP on natural hair colour. We observe similar odd ratios (ORs) for risk of CAD per standard deviation increase in SBP across various medication-adjustment methods, although there is increased precision with methods using larger number of SNPs. With age-restricted cohorts, we observe an attenuation of OR towards the null with increasing age. In MVMR analyses with binary treatment status as a second exposure, we observe bias towards the null. This can be attributed to low F-statistics for SBP conditional on antihypertensive use, and antihypertensive use conditional on SBP (conditional F-statistics: 8.6 and 8.3 respectively). This suggests the presence of weak instrument bias, which is illustrated in Figure 5 (Sanderson, Spiller, et al., 2021).

**Figure 4.**
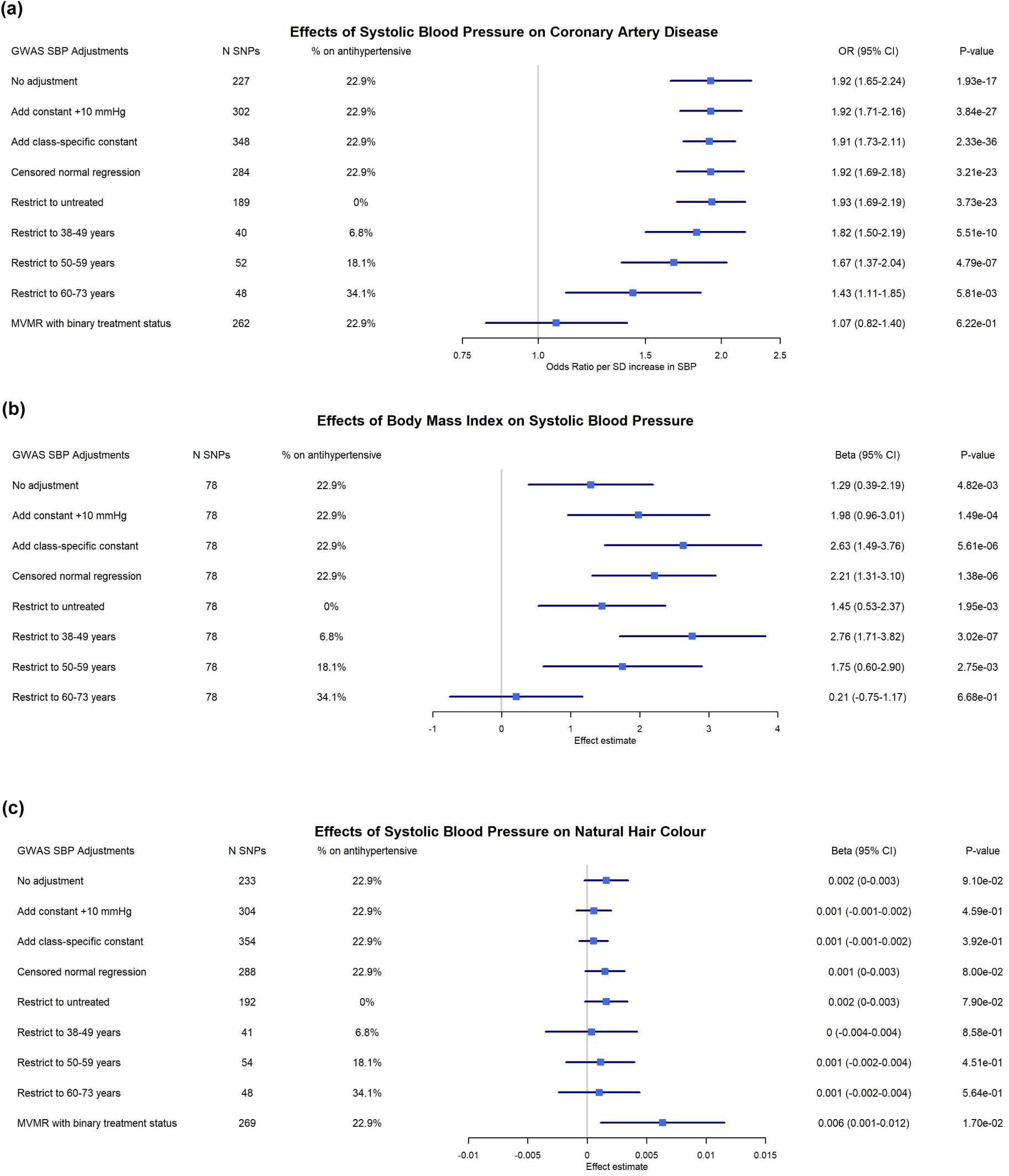
Inverse variance weighted MR results. GWAS: genome-wide association study; MR: mendelian randomization; MVMR: multivariable mendelian randomization; SBP: systolic blood pressure; SD: standard deviation; SNPs: single nucleotide polymorphisms Summary-level data MR inverse variance weighted estimates are shown in the figure, with corresponding 95% confidence intervals and p-values for each applied adjustment method. The following effects are estimated: (a) odds ratio for each standard deviation increase in SBP on coronary artery disease, (b) Effect estimates of body mass index on SBP, (c) effect estimates of SBP on natural hair colour as a negative control outcome. There is less variation in effects when SBP is the exposure, but more variation in effects when SBP is the outcome (and when the exposure influences the probability of receiving treatment and hence altering the outcome phenotype level directly). MVMR analysis was subjected to weak instrument bias.

**Figure 5.**
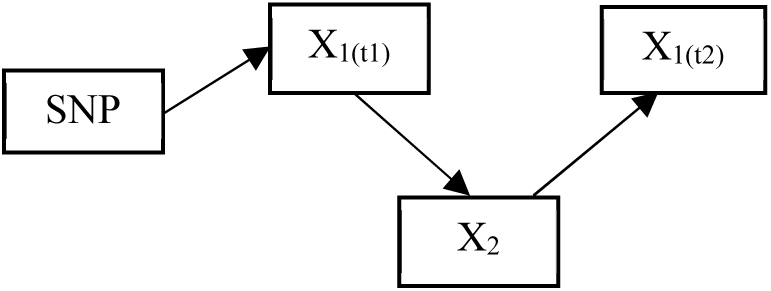
Weak instrument bias in multivariable mendelian randomization. SNP: single nucleotide polymorphism, used as an instrumental variable in a mendelian randomization analysis X_1(t1)_ represents the first exposure at time 1, X_1(t2)_ represents the first exposure at time 2, and X_2_ represents the second exposure. In our study, X_1(t1)_ is untreated systolic blood pressure, X_2_ is antihypertensive use, X_1(t2)_ is measured systolic blood pressure. We postulate this relationship between the variables which could result in the observed weak instrument bias. Although both X_1(t2)_ and X_2_ could be both robustly associated with identified SNPs, the association of X_1(t2)_ conditional on X_2_ and X_2_ conditional on X_1(t2)_ can be weak.

In our MR analysis of the effect of BMI on SBP (Figure 4b), we observe larger variation in the effects of BMI on SBP across various medication-adjustment methods. The largest effect estimates per unit increase in BMI was when we added class-specific constants (beta=2.63, 95% CI: 1.49-3.76).

Unadjusted analyses yielded similar effect estimates as restricting to untreated individuals – a potential explanation is that both approaches underrepresent or mask the highest or most extreme of blood pressure values within the cohort. Restricting analyses to 38-49 years resulted in similar findings as to when other direct adjustment methods were applied. We also observed an attenuation of effect estimates towards the null with increasing age groups (with increasing proportion of treated individuals).

In negative control analyses of SBP with natural hair colour, confidence intervals spanned the null with all adjustment methods excepting MVMR analyses which showed positive results. F-statistics for all univariable MR IVW analyses can be found in *Supplementary Table 6*.

## 4. Discussion

Adjusting for medication use is currently only applied in a limited number of settings, and even when performed, its appropriateness has been rarely assessed in practice. In this study therefore, we wanted to evaluate various medication adjustment methods and its impact on significance of genetic associations, strength of effect estimates and downstream MR estimates.

Our findings support our initial hypothesis – that effect estimates of genetic variants strongly associated with high SBP are deflated when no medication adjustment is applied. The largest difference can be observed when we compare between the most extreme of groups (e.g., between treated versus untreated, or, in restricted population of 60-73 years versus unrestricted and unadjusted population). This aligns with our expectation that the bias associated with unmodelled medication use is more pronounced in more heavily treated populations. An alternative explanation however is selection bias – that, by restricting the GWAS to individuals aged 60-73 years, we are conditioning on a ‘healthier’ population having survived till an older age. That is, more susceptible genotypes may have died or were not able to participate in the UK Biobank study. We also cannot dismiss potential age-varying loci effects for SBP, although current evidence suggests that this is not substantial in UK Biobank (Leyden et al., 2026).

Across medication-adjustment methods, we found that applying direct adjustments to measured SBP values could unmask stronger GWAS effect estimates, detect new genetic signals as a consequence of adjustment and retain power because of the preservation of sample size. Adjustment by adding class-specific constants on average resulted in the strongest effect estimates and most significant p-values. In contrast, restriction-based approaches (i.e., restrict to untreated, restrict to 38-49 years) identified fewer variants with less significant p-values, although effect estimates were variable. This is likely attributed to smaller sample sizes and hence lower power. We also observed that SNP heritability also differed by medication-adjustment method, varying by more than 5% between unadjusted and medication-adjusted analyses.

We showed that the various direct medication-adjusted GWAS approaches could reveal new genetic associations for SBP trait (Supplementary Table 7). We discuss a few of them here, with potential relevance to SBP through cardiometabolic pathways. *CDKAL1* has been associated with insulin secretion, and regulation of mitochondrial function in adipose tissue (Ghosh et al., 2022; Palmer et al., 2017; Watanabe et al., 2013). *RORA* is known to regulate lipid metabolism and has been linked with atherosclerosis (Lau et al., 2008; Lee et al., 2021; Mamontova et al., 1998). *SLC39A14*, with known zinc and manganese transporter function, has been shown to contribute to endosomal insulin receptor trafficking and glucose metabolism (Aydemir et al., 2016; Aydemir & Cousins, 2018; Maxel et al., 2019). *PALLD* has been speculated to play roles in cardiovascular disease (Jin, 2011). *HSD17B12* has been shown to have crucial functions for metabolic homeostasis in mice (Heikelä et al., 2020; Rantakari et al., 2010). *PKHD1* has been implicated in autosomal recessive polycystic kidney disease which features hypertension at a young age; although rare, it could be plausible a common variation near this gene could influence blood pressure through the renal pathway (Ward et al., 2002). These suggest that medication adjustment shows promise in enhancing our understanding of a trait, although exact mechanisms and its relevance to blood pressure should be explored in future studies.

We also observed there were several independent genetic variants with heterogeneous effect estimates between the unadjusted and medication-adjusted GWAS. Most variants were not mapped to canonical hypertension loci (Table 3). However, of the heterogeneous variants, *KCNK3* has been recently linked to resistant hypertension (Armstrong et al., 2023; Ebinger et al., 2026; Yamazaki et al., 2023). *FGF5* was also previously identified as a locus for blood pressure (Park et al., 2014). *NPNT* and *FGF5* were also previously discovered in a large European discovery meta-analysis of 757,601 participants (Evangelou et al., 2018): rs13112725 (phenotype: SBP, beta: 0.414, SE: 0.036) and rs16998073 (phenotype: *diastolic* blood pressure, beta: -0.494, SE: 0.019) respectively. For rs13112725, we observed an interesting trend of beta [SE] values in our study: 0.293 [0.045] (no adjustment), 0.369 [0.047] (add constant +10mmHg), 0.425 [0.050] (add class-specific constant), 0.640 [0.083] (restrict to 38-49 years). We postulate that younger individuals with this variant may have little of other lifestyle factors which increases SBP and hence may benefit most from treatment. This may lead to increased effect estimates when we apply medication adjustment in this subgroup as compared to the general population. This further emphasizes that applying restrictions to cohorts can change effect estimates which requires further consideration of its implications when used in downstream analyses.

Heterogeneous variants in the *untreated versus treated* GWAS also revealed genes which have been previously reported to be associated with blood pressure. *WNT2B* has been discussed to contribute to resistant hypertension, with potential links through primary aldosteronism (Ebinger et al., 2026; Naito et al., 2023; Yamazaki et al., 2023). *CACNB2* was identified in the CHARGE targeted-sequencing study as a blood pressure candidate region, and in Biobank Japan as a gene influencing the effectiveness of angiotensin II receptor blocker treatment (Morrison et al., 2014; Yamazaki et al., 2023). *ADRB1* is a β1-adrenergic receptor gene and its polymorphisms can contribute to differential susceptibility to hypertension. (Katsukunya et al., 2024; Pacanowski et al., 2008). *NT5C2* was shown to be likely causal for hypertension in zebrafish (Vishnolia et al., 2020). *EBF2* was reported as a new locus for blood pressure (Wain et al., 2017). These show that analyses comparing effect estimates between untreated and treated populations could reveal novel locus in traits of interest. In totality, applying medication adjustments to measured SBP may unmask pleiotropic effects or reveal potential secondary pathways affecting SBP, although it is also possible that these are statistical artefacts which have arisen by chance or by sample structure (subgroups).

In MR analyses of the causal effect of SBP on CAD, we found that there was little difference in observed odd ratios between unadjusted and medication-adjusted approaches. However, we observed an attenuation towards the null with increasing age group. This could suggest that in heavily treated populations, it is more challenging to accurately estimate effects without sufficient medication-adjustments. It is plausible that if treatment is not accounted for in the GWAS of heavily treated populations (i.e., in 60-73 years), direct SNP effects on SBP are likely deflated or even inverted. This could occur if a variant strongly predisposing to high unmedicated SBP was statistically associated with reduced SBP due to treatment effects in the cohort. As a consequence, we would observe deflated causal estimates for the effect of SBP on CAD, especially since treatment had not been accounted for in the CAD GWAS. Adjusting the CAD GWAS itself for medication use may be inappropriate. Treatment status is a common consequence of both SBP and other risk factors for CAD, making it a collider. Conditioning on it would open a non-causal path linking the genetic instrument to CAD and thereby bias the estimate. This is illustrated by the path Genotype → SBP → [Treatment] ← CAD risk factors → CAD, where brackets denote conditioning.

We also found a biased estimate in the MVMR analysis as a likely result of weak instrument bias (conditional F-statistic<10 in both exposures). Figure 5 provides a possible explanation of the observed findings. Although we had selected SNPs robustly associated with both measured SBP and binary treatment status, we still observed conditionally weak instruments, indicating that the SNPs were unable to jointly strongly predict both exposures and the analysis was likely to be biased by weak instrument bias. MVMR effect estimates can be biased by weak instruments away from the null, which was observed in negative control analyses (Sanderson, Spiller, et al., 2021).

When examining the effect of BMI on SBP, we observed larger variations of IVW effects across various medication-adjustment methods. A possible explanation for this finding is that BMI causally influences whether an individual would be prescribed antihypertensives or not. More simply, the higher an individual’s BMI sits outside the ‘healthy’ range, the more likely the individual would receive treatment. This is because the individual with a higher BMI would have increased risks of cardiometabolic diseases which indicates antihypertensive therapy, and being on antihypertensives would directly affect measured SBP values. This would imply that when the phenotype of interest affected by medications is the outcome, and when the exposure can directly affect the probability of receiving medications, then considerations for medication adjustment becomes increasingly more important. While it is difficult to claim a single ‘gold standard’ method, our findings suggest that in the case of BMI and SBP, unadjusted analyses biases effect estimates towards the null, although it is still quite clear from our analyses that each kg/m^2^ increase in BMI leads to an increase in SBP.

When considering medication-adjustment for the SBP trait, we would recommend adding class-specific constants to approximate untreated SBP. This approach more accurately estimates untreated SBP given that it accounts for the number and type of antihypertensives used. After all, use of more than two antihypertensives is indicative of more severe or treatment-resistant hypertension (National Institute for Health and Care Excellence, 2019; Smith et al., 2020). We recognise that this is not always feasible, and hence we make the following recommendations: If precision of downstream estimates are important, or if the investigator wishes to discover new genetic associations, then it would be ideal to make every effort to derive a more accurate phenotype. Various direct medication adjustments could also be applied to examine how sensitive estimates are. However, to broadly ascertain causal direction or relationship (e.g., using MR), we have shown that it does not largely affect conclusions (NB: the investigator should apply more caution when estimating effects of characteristics that affect probability of treatment on traits that are affected by that treatment). We do not favour restriction-based approaches, as it can introduce selection or collider-stratification bias (Hernán et al., 2004; Munafò et al., 2018). This is because treatment status is also dependent on many other factors besides untreated SBP, such as ethnicity or age (Rison et al., 2023). Hence the GWAS beta values may contain some effect of non-SBP treatment factors, which may induce bias in downstream analyses.

When performing large-scale GWAS meta-analyses, investigators should consider if medication adjustment had been applied, and the type of adjustment that had been applied in each study. Investigators should also consider reporting stratified results by type of adjustment. Future work could examine the impact of combining different medication adjustments together, and to investigate the potential biases which might arise. Also, for summary statistics which do not apply a medication adjustment, future work could examine the utility of applying a factor adjustment to correct for deflated effect estimates. This could be applied to genome-wide significant variants in a sensitivity analysis before proceeding to conduct downstream analyses. In our cohort, with 23.1% of individuals on antihypertensive treatment, we observe an average inflation of effect estimates by 17-28% (Figure 2, slope) across genome-wide significant variants when adjusting SBP measures by adding constants or class-specific constants. This however assumes that effects of treatment is uniform across all variants, which is likely untrue.

In addition, we have examined SBP in detail here, but other phenotypes could be examined in future studies. A feasible example is adjusting for statin use on LDL-c, or hypoglycaemics on HbA1c. However, we recognise that the relationship between phenotypes and medications are not always as straightforward to conceptualise – adjusting for mental health medications for example on depression scores poses a potential challenge. For instance, it is difficult to accurately quantify how much each antidepressant improves depression symptoms given variable treatment response and heterogeneity of depression (Athira et al., 2020). Future work can also examine the impact of other exposures which can increase probability of treatment to evaluate extent of biases to downstream estimates. For SBP, these may include factors relating to obesity (waist-to-hip ratio, waist circumference), and risk factors for cardiovascular disease (diabetes/insulin resistance, high LDL-c). In the next decade, with increasing knowledge on precision medicine and what drives an individual’s response to SBP, future work can explore an individual-level correction (rather than the average class-specific one we have applied here) (Denny & Collins, 2021; Mendez et al., 2025). This could include prediction of treatment response, including treatment-resistant hypertension, or other additional variables like adherence or duration of treatment.

A major strength of this study is its systematic assessment of how various medication adjustment methods can change estimates of direct genetic effects in GWAS of a physiological measure like systolic blood pressure. Furthermore, we describe and discuss the impact of instrumenting these genetic variants in univariable and multivariable MR when the phenotype of interest is the exposure or outcome. As UK Biobank is a very comprehensive resource with information of medications use collected at baseline assessment visit, we were able to ascertain medication subclasses and test a novel method of adjusting for both the use of antihypertensives while taking into consideration the type and number of antihypertensives patients were on. However, we have assumed that medications have purely additive effects which may be overly simplistic.

The UK Biobank is also known to be a highly selected sample, sampling more educated and healthy individuals than the general population (Schoeler et al., 2023). In our analyses, the proportion of the cohort treated with antihypertensives seemed to be relatively low, even among older age groups (34.1%). In addition, treatment-seeking has been shown to vary meaningfully across socio-economic, educational and ethnic groups within the UK Biobank. Here we have only considered the correction of SBP by receipt of treatment, assuming a uniform likelihood of treatment given a certain latent hypertensive propensity. However, there are likely further corrections that may improve this model using demographic subgroups. For instance, some SNPs may be associated with treatment receipt independently of SBP, such as through education or experience of healthcare discrimination. If medication use is not modelled explicitly, the SBP effect estimates for these SNPs would be increased. Whilst this is as yet unlikely to impact analyses thresholding at genome-wide significance, more subtle biases such as these may be more impactful for less strictly filtered genetic signals. In addition, as UK Biobank consists of predominantly White British data, and because we chose the White British cohort to avoid potential issues with population stratification, we are unable to generalise these findings to non-White British ethnicities; future studies could be conducted in other biobanks cohorts.

## 5. Conclusion

In conclusion, using systolic blood pressure as an example, we have demonstrated that not adjusting for medication use in GWAS can lead to deflation in the estimates of direct genetic effects. To examine the potential of bias arising from inadequate or no adjustment for medication use, we recommend applying different medication-adjustment methods in GWAS where feasible. Such results can then be used as sensitivity analyses in any downstream work such as heritability estimation, polygenic scores or Mendelian randomization. This is especially important in heavily treated populations where effect estimates are likely most strongly attenuated, or if the goal of the investigator is to obtain precise estimates or discover new genetic loci to target. Adjustment methods may include direct modifications of phenotypic values or restriction to untreated (or younger) individuals; however, restriction-based approaches detect fewer genetic signals due to smaller sample sizes. For systolic blood pressure, we found that the most common approach of adjusting for antihypertensives by adding a constant yielded results consistent with those obtained using alternative adjustment methods. However, we favour adding class-specific constants which we suggest better approximates untreated SBP. Greater caution is needed when estimating the effects of characteristics (that influence treatment decisions) on traits that are affected by that treatment in downstream analyses such as Mendelian randomization. Future work can examine medication adjustment in other phenotypes, especially medications which are commonly used in the population.

## Data Availability

The individual-level data that support the findings of this study are available with the permission of the UK Biobank (http://www.ukbiobank.ac.uk). We conducted this study using the UK Biobank resource under an approved data application (ref: 81499). The GWAS summary statistics of systolic blood pressure with various medication adjustments produced in the present study is or will be (at the time of submission) publicly available on OpenGWAS. Other GWAS summary statistics used in the study for Mendelian randomization analyses are available on OpenGWAS (https://opengwas.io/).

https://opengwas.io/

## Acknowledgements

This research has been conducted using the UK Biobank Resource under Application Number 81499.

## Conflict of interest disclosure

The authors have no conflicts of interest to declare.

## Funding statement

AJYY is supported by the Wellcome Trust (218495/Z/19/Z). ES is supported by the Medical Research Council (UKRI077). The MRC Integrative Epidemiology Unit is supported by the Medical Research Council (MC_UU_00032/1) and the University of Bristol. GJG is supported by an MQ Fellows Award (MQF22/22).

## Notes

### Competing Interest Statement

The authors have declared no competing interest.

### Author Declarations

UK Biobank received ethical approval from the Research Ethics Committee (REC reference: 11/NW/0382). This research has been conducted using the UK Biobank Resource under Application Number 81499.

### Summary of Updates

Table 3 revised; Section 3.5.: Additional SNP-based heritability across different GWAS conducted; Section 3.6.: Novel independent genetic variants from medication-adjusted GWAS reported and discussed; Introduction, Methods, Results and Discussion revised; Supplemental files updated

## References

Armstrong, N. D., Srinivasasainagendra, V., Ammous, F., Assimes, T. L., Beitelshees, A. L., Brody, J., Cade, B. E., Ida Chen, Y.-D., Chen, H., de Vries, P. S., Floyd, J. S., Franceschini, N., Guo, X., Hellwege, J. N., House, J. S., Hwu, C.-M., Kardia, S. L. R., Lange, E. M., Lange, L. A., … Arnett, D. K. (2023). Whole genome sequence analysis of apparent treatment resistant hypertension status in participants from the Trans-Omics for Precision Medicine program. Frontiers in Genetics, 14. 10.3389/fgene.2023.1278215

Aschard, H., Vilhjálmsson, B. J., Joshi, A. D., Price, A. L., & Kraft, P. (2015). Adjusting for Heritable Covariates Can Bias Effect Estimates in Genome-Wide Association Studies. American Journal of Human Genetics, 96(2), 329. 10.1016/j.ajhg.2014.12.021

Athira, K. V., Bandopadhyay, S., Samudrala, P. K., Naidu, V. G. M., Lahkar, M., & Chakravarty, S. (2020). An Overview of the Heterogeneity of Major Depressive Disorder: Current Knowledge and Future Prospective. Current Neuropharmacology, 18(3), 168–187. 10.2174/1570159X17666191001142934

Aydemir, T. B., & Cousins, R. J. (2018). The Multiple Faces of the Metal Transporter ZIP14 (SLC39A14). The Journal of Nutrition, 148(2), 174–184. 10.1093/jn/nxx041

Aydemir, T. B., Troche, C., Kim, M.-H., & Cousins, R. J. (2016). Hepatic ZIP14-mediated Zinc Transport Contributes to Endosomal Insulin Receptor Trafficking and Glucose Metabolism*. Journal of Biological Chemistry, 291(46), 23939–23951. 10.1074/jbc.M116.748632

Barendse, W. (2011). The effect of measurement error of phenotypes on genome wide association studies. BMC Genomics, 12, 232. 10.1186/1471-2164-12-232

Bulik-Sullivan, B. K., Loh, P.-R., Finucane, H. K., Ripke, S., Yang, J., Patterson, N., Daly, M. J., Price, A. L., & Neale, B. M. (2015). LD Score regression distinguishes confounding from polygenicity in genome-wide association studies. Nature Genetics, 47(3), 291–295. 10.1038/ng.3211

Burgess, S., & Thompson, S. G. (2015). Multivariable Mendelian Randomization: The Use of Pleiotropic Genetic Variants to Estimate Causal Effects. American Journal of Epidemiology, 181(4), 251–260. 10.1093/aje/kwu283

Bycroft, C., Freeman, C., Petkova, D., Band, G., Elliott, L. T., Sharp, K., Motyer, A., Vukcevic, D., Delaneau, O., O’Connell, J., Cortes, A., Welsh, S., Young, A., Effingham, M., McVean, G., Leslie, S., Allen, N., Donnelly, P., & Marchini, J. (2018). The UK Biobank resource with deep phenotyping and genomic data. Nature, 562(7726), 203–209. 10.1038/s41586-018-0579-z

Chang, C. C., Chow, C. C., Tellier, L. C., Vattikuti, S., Purcell, S. M., & Lee, J. J. (2015). Second-generation PLINK: Rising to the challenge of larger and richer datasets. Gigascience, 4(1), s13742–015-0047–0048. 10.1186/s13742-015-0047-8

Cho, Y. S., Go, M. J., Kim, Y. J., Heo, J. Y., Oh, J. H., Ban, H.-J., Yoon, D., Lee, M. H., Kim, D.-J., Park, M., Cha, S.-H., Kim, J.-W., Han, B.-G., Min, H., Ahn, Y., Park, M. S., Han, H. R., Jang, H.-Y., Cho, E. Y., … Kim, H.-L. (2009). A large-scale genome-wide association study of Asian populations uncovers genetic factors influencing eight quantitative traits. Nature Genetics, 41(5), 527–534. 10.1038/ng.357

Chong, A. H. W., Kintu, C., Cho, Y., Fatumo, S., Torres, J., Smith, G. D., Gaunt, T. R., & Hemani, G. (2024). Adjusting for medication status in genome-wide association studies (p. 2024.02.19.24303028). medRxiv. 10.1101/2024.02.19.24303028

Cole, S. R., Platt, R. W., Schisterman, E. F., Chu, H., Westreich, D., Richardson, D., & Poole, C. (2010). Illustrating bias due to conditioning on a collider. International Journal of Epidemiology, 39(2), 417–420. 10.1093/ije/dyp334

Collins, R. (2012). What makes UK Biobank special? The Lancet, 379(9822), 1173–1174. 10.1016/S0140-6736(12)60404-8

Davey Smith, G., & Ebrahim, S. (2003). ‘Mendelian randomization’: Can genetic epidemiology contribute to understanding environmental determinants of disease?*. International Journal of Epidemiology, 32(1), 1–22. 10.1093/ije/dyg070

Davies, N. M., Hemani, G., Neiderhiser, J. M., Martin, H. C., Mills, M. C., Visscher, P. M., Yengo, L., Young, A. S., & Keller, M. C. (2024). The importance of family-based sampling for biobanks. Nature, 634(8035), 795–803. 10.1038/s41586-024-07721-5

Denny, J. C., & Collins, F. S. (2021). Precision medicine in 2030—Seven ways to transform healthcare. Cell, 184(6), 1415–1419. 10.1016/j.cell.2021.01.015

Ebinger, J. E., Kauko, A., Vaura, F., Hage, P., Sundström, J., FinnGen, Joung, S. Y., Cheng, S., & Niiranen, T. J. (2026). GWAS and Replication Analysis of Apparent Treatment-Resistant Hypertension. Hypertension, 83(2), e25719. 10.1161/HYPERTENSIONAHA.125.25719

Evangelou, E., Warren, H. R., Mosen-Ansorena, D., Mifsud, B., Pazoki, R., Gao, H., Ntritsos, G., Dimou, N., Cabrera, C. P., Karaman, I., Ng, F. L., Evangelou, M., Witkowska, K., Tzanis, E., Hellwege, J. N., Giri, A., Velez Edwards, D. R., Sun, Y. V., Cho, K., … Caulfield, M. J. (2018). Genetic analysis of over one million people identifies 535 new loci associated with blood pressure traits. Nature Genetics, 50(10), 1412–1425. 10.1038/s41588-018-0205-x

Ghosh, C., Das, N., Saha, S., Kundu, T., Sircar, D., & Roy, P. (2022). Involvement of Cdkal1 in the etiology of type 2 diabetes mellitus and microvascular diabetic complications: A review. Journal of Diabetes & Metabolic Disorders, 21(1), 991–1001. 10.1007/s40200-021-00953-6

Gilbody, J., Borges, M. C., Davey Smith, G., & Sanderson, E. (2025). Multivariable MR Can Mitigate Bias in Two-Sample MR Using Covariable-Adjusted Summary Associations. Genetic Epidemiology, 49(1), e22606. 10.1002/gepi.22606

Hartwig, F. P., Tilling, K., Davey Smith, G., Lawlor, D. A., & Borges, M. C. (2021). Bias in two-sample Mendelian randomization when using heritable covariable-adjusted summary associations. International Journal of Epidemiology, 50(5), 1639–1650. 10.1093/ije/dyaa266

Heikelä, H., Ruohonen, S. T., Adam, M., Viitanen, R., Liljenbäck, H., Eskola, O., Gabriel, M., Mairinoja, L., Pessia, A., Velagapudi, V., Roivainen, A., Zhang, F.-P., Strauss, L., & Poutanen, M. (2020). Hydroxysteroid (17β) dehydrogenase 12 is essential for metabolic homeostasis in adult mice. American Journal of Physiology-Endocrinology and Metabolism, 319(3), E494– E508. 10.1152/ajpendo.00042.2020

Hemani, G., Elsworth, B., Palmer, T., & Rasteiro, R. (2024). ieugwasr: Interface to the ‘OpenGWAS’ Database API*. R package version 1.0.2*. https://mrcieu.github.io/ieugwasr/

Hemani, G., Tilling, K., & Smith, G. D. (2017). Orienting the causal relationship between imprecisely measured traits using GWAS summary data. PLOS Genetics, 13(11), e1007081. 10.1371/journal.pgen.1007081

Hemani, G., Zheng, J., Elsworth, B., Wade, K. H., Haberland, V., Baird, D., Laurin, C., Burgess, S., Bowden, J., Langdon, R., Tan, V. Y., Yarmolinsky, J., Shihab, H. A., Timpson, N. J., Evans, D. M., Relton, C., Martin, R. M., Davey Smith, G., Gaunt, T. R., & Haycock, P. C. (2018). The MR-Base platform supports systematic causal inference across the human phenome. eLife, 7, e34408. 10.7554/eLife.34408

Hernán, M. A., Hernández-Díaz, S., & Robins, J. M. (2004). A Structural Approach to Selection Bias. Epidemiology, 15(5), 615. 10.1097/01.ede.0000135174.63482.43

Hirschhorn, J. N., & Daly, M. J. (2005). Genome-wide association studies for common diseases and complex traits. Nature Reviews Genetics, 6(2), 95–108. 10.1038/nrg1521

Holm, S. (1979). A Simple Sequentially Rejective Multiple Test Procedure. Scandinavian Journal of Statistics, 6(2), 65–70.

Hu, S., Ferreira, L. A. F., Shi, S., Hellenthal, G., Marchini, J., Lawson, D. J., & Myers, S. R. (2025). Fine-scale population structure and widespread conservation of genetic effect sizes between human groups across traits. Nature Genetics, 57(2), 379–389. 10.1038/s41588-024-02035-8

Jiang, L., Zheng, Z., Qi, T., Kemper, K. E., Wray, N. R., Visscher, P. M., & Yang, J. (2019). A resource-efficient tool for mixed model association analysis of large-scale data. Nature Genetics, 51(12), 1749–1755. 10.1038/s41588-019-0530-8

Jin, L. (2011). The actin associated protein palladin in smooth muscle and in the development of diseases of the cardiovasculature and in cancer. Journal of Muscle Research and Cell Motility, 32(1), 7–17. 10.1007/s10974-011-9246-9

Katsukunya, J. N., Jones, E., Soko, N. D., Blom, D., Sinxadi, P., Rayner, B., & Dandara, C. (2024). Genetic Variation in ABCB1, ADRB1, CYP3A4, CYP3A5, NEDD4L and NR3C2 Confers Differential Susceptibility to Resistant Hypertension among South Africans. Journal of Personalized Medicine, 14(7), 664. 10.3390/jpm14070664

Keaton, J. M., Kamali, Z., Xie, T., Vaez, A., Williams, A., Goleva, S. B., Ani, A., Evangelou, E., Hellwege, J. N., Yengo, L., Young, W. J., Traylor, M., Giri, A., Zheng, Z., Zeng, J., Chasman, D. I., Morris, A. P., Caulfield, M. J., Hwang, S.-J., … Warren, H. R. (2024). Genome-wide analysis in over 1 million individuals of European ancestry yields improved polygenic risk scores for blood pressure traits. Nature Genetics, 56(5), 778–791. 10.1038/s41588-024-01714-w

Kim, Y. J., Go, M. J., Hu, C., Hong, C. B., Kim, Y. K., Lee, J. Y., Hwang, J.-Y., Oh, J. H., Kim, D.-J., Kim, N. H., Kim, S., Hong, E. J., Kim, J.-H., Min, H., Kim, Y., Zhang, R., Jia, W., Okada, Y., Takahashi, A., … Cho, Y. S. (2011). Large-scale genome-wide association studies in east Asians identify new genetic loci influencing metabolic traits. Nature Genetics, 43(10), 990– 995. 10.1038/ng.939

Lau, P., Fitzsimmons, R. L., Raichur, S., Wang, S.-C. M., Lechtken, A., & Muscat, G. E. O. (2008). The Orphan Nuclear Receptor, RORα, Regulates Gene Expression That Controls Lipid Metabolism: STAGGERER (SG/SG) MICE ARE RESISTANT TO DIET-INDUCED OBESITY*. Journal of Biological Chemistry, 283(26), 18411–18421. 10.1074/jbc.M710526200

Law, M. R. (2003). Value of low dose combination treatment with blood pressure lowering drugs: Analysis of 354 randomised trials. BMJ, 326(7404), 1427–0. 10.1136/bmj.326.7404.1427

Lawson, D. (2022). *R package pcapred* [R]. https://github.com/danjlawson/pcapred

Lee, J. M., Kim, H., & Baek, S. H. (2021). Unraveling the physiological roles of retinoic acid receptor-related orphan receptor α. Experimental & Molecular Medicine, 53(9), 1278–1286. 10.1038/s12276-021-00679-8

Leyden, G. M., Pagoni, P., Power, G. M., Carslake, D., Richardson, T. G., Tilling, K., Hemani, G., Davey Smith, G., & Sanderson, E. (2026). An evaluation of age-varying genetic effects underlying body-mass index and blood pressure in the UK Biobank. PLOS Genetics, 22(3), e1012080. 10.1371/journal.pgen.1012080

Locke, A. E., Kahali, B., Berndt, S. I., Justice, A. E., Pers, T. H., Day, F. R., Powell, C., Vedantam, S., Buchkovich, M. L., Yang, J., Croteau-Chonka, D. C., Esko, T., Fall, T., Ferreira, T., Gustafsson, S., Kutalik, Z., Luan, J., Mägi, R., Randall, J. C., … Speliotes, E. K. (2015). Genetic studies of body mass index yield new insights for obesity biology. Nature, 518(7538), 197–206. 10.1038/nature14177

Lucas, A., Verma, A., & Ritchie, M. D. (2022). hudson: A User-Friendly R Package to Extend Manhattan Plots (p. 2022.01.25.474274). bioRxiv. 10.1101/2022.01.25.474274

Magno, R., & Maia, A.-T. (2020). gwasrapidd: An R package to query, download and wrangle GWAS catalog data. Bioinformatics, 36(2), 649–650. 10.1093/bioinformatics/btz605

Mamontova, A., Séguret-Macé, S., Esposito, B., Chaniale, C., Bouly, M., Delhaye-Bouchaud, N., Luc, G., Staels, B., Duverger, N., Mariani, J., & Tedgui, A. (1998). Severe Atherosclerosis and Hypoalphalipoproteinemia in the Staggerer Mouse, a Mutant of the Nuclear Receptor RORα. Circulation, 98(24), 2738–2743. 10.1161/01.CIR.98.24.2738

Marchini, J. (2015). UK Biobank Phasing and Imputation Documentation Version 1.2. https://biobank.ctsu.ox.ac.uk/crystal/crystal/docs/impute_ukb_v1.pdf

Maxel, T., Smidt, K., Petersen, C. C., Honoré, B., Christensen, A. K., Jeppesen, P. B., Brock, B., Rungby, J., Palmfeldt, J., & Larsen, A. (2019). The zinc transporter Zip14 (SLC39a14) affects Beta-cell Function: Proteomics, Gene expression, and Insulin secretion studies in INS-1E cells. Scientific Reports, 9(1), 8589. 10.1038/s41598-019-44954-1

McCarthy, M. I., Abecasis, G. R., Cardon, L. R., Goldstein, D. B., Little, J., Ioannidis, J. P. A., & Hirschhorn, J. N. (2008). Genome-wide association studies for complex traits: Consensus, uncertainty and challenges. Nature Reviews Genetics, 9(5), 356–369. 10.1038/nrg2344

Mendez, K. M., Reinke, S. N., Kelly, R. S., Chen, Q., Su, M., McGeachie, M., Weiss, S., Broadhurst, D. I., & Lasky-Su, J. A. (2025). A roadmap to precision medicine through post-genomic electronic medical records. Nature Communications, 16(1), 1700. 10.1038/s41467-025-56442-4

Mitchell, R., Hemani, G., Dudding, T., Corbin, L., Harrison, S., & Paternoster, L. (2019). UK Biobank Genetic Data: MRC-IEU Quality Control, version 2. 10.5523/bris.1ovaau5sxunp2cv8rcy88688v

Morrison, A. C., Bis, J. C., Hwang, S.-J., Ehret, G. B., Lumley, T., Rice, K., Muzny, D., Gibbs, R. A., Boerwinkle, E., Psaty, B. M., Chakravarti, A., & Levy, D. (2014). Sequence Analysis of Six Blood Pressure Candidate Regions in 4,178 Individuals: The Cohorts for Heart and Aging Research in Genomic Epidemiology (CHARGE) Targeted Sequencing Study. PLoS ONE, 9(10), e109155. 10.1371/journal.pone.0109155

Munafò, M. R., Tilling, K., Taylor, A. E., Evans, D. M., & Davey Smith, G. (2018). Collider scope: When selection bias can substantially influence observed associations. International Journal of Epidemiology, 47(1), 226–235. 10.1093/ije/dyx206

Naito, T., Inoue, K., Sonehara, K., Baba, R., Kodama, T., Otagaki, Y., Okada, A., Itcho, K., Kobuke, K., Kishimoto, S., Yamamoto, K., BioBank Japan, Morisaki, T., Higashi, Y., Hinata, N., Arihiro, K., Hattori, N., Okada, Y., & Oki, K. (2023). Genetic Risk of Primary Aldosteronism and Its Contribution to Hypertension: A Cross-Ancestry Meta-Analysis of Genome-Wide Association Studies. Circulation, 147(14), 1097–1109. 10.1161/CIRCULATIONAHA.122.062349

National Institute for Health and Care Excellence. (2019, August 28). Hypertension in adults: Diagnosis and management. NICE. https://www.nice.org.uk/guidance/ng136

Nikpay, M., Goel, A., Won, H.-H., Hall, L. M., Willenborg, C., Kanoni, S., Saleheen, D., Kyriakou, T., Nelson, C. P., Hopewell, J. C., Webb, T. R., Zeng, L., Dehghan, A., Alver, M., Armasu, S. M., Auro, K., Bjonnes, A., Chasman, D. I., Chen, S., … the CARDIoGRAMplusC4D Consortium. (2015). A comprehensive 1000 Genomes–based genome-wide association meta-analysis of coronary artery disease. Nature Genetics, 47(10), 1121–1130. 10.1038/ng.3396

Pacanowski, M., Gong, Y., Cooper-DeHoff, R., Schork, N., Shriver, M., Langaee, T., Pepine, C., & Johnson, J. (2008). β-Adrenergic Receptor Gene Polymorphisms and β-Blocker Treatment Outcomes in Hypertension. Clinical Pharmacology & Therapeutics, 84(6), 715–721. 10.1038/clpt.2008.139

Palmer, C. J., Bruckner, R. J., Paulo, J. A., Kazak, L., Long, J. Z., Mina, A. I., Deng, Z., LeClair, K. B., Hall, J. A., Hong, S., Zushin, P.-J. H., Smith, K. L., Gygi, S. P., Hagen, S., Cohen, D. E., & Banks, A. S. (2017). Cdkal1, a type 2 diabetes susceptibility gene, regulates mitochondrial function in adipose tissue. Molecular Metabolism, 6(10), 1212–1225. 10.1016/j.molmet.2017.07.013

Parikh, N. I., Pencina, M. J., Wang, T. J., Benjamin, E. J., Lanier, K. J., Levy, D., D’Agostino, R. B., Kannel, W. B., & Vasan, R. S. (2008). A Risk Score for Predicting Near-Term Incidence of Hypertension: The Framingham Heart Study. Annals of Internal Medicine, 148(2), 102. 10.7326/0003-4819-148-2-200801150-00005

Park, S. Y., Lee, H.-J., Ji, S.-M., Kim, M. E., Jigden, B., Lim, J. E., & Oh, B. (2014). ANTXR2 is a potential causative gene in the genome-wide association study of the blood pressure locus 4q21. Hypertension Research, 37(9), 811–817. 10.1038/hr.2014.84

Pierce, B. L., & Burgess, S. (2013). Efficient Design for Mendelian Randomization Studies: Subsample and 2-Sample Instrumental Variable Estimators. American Journal of Epidemiology, 178(7), 1177–1184. 10.1093/aje/kwt084

Rantakari, P., Lagerbohm, H., Kaimainen, M., Suomela, J.-P., Strauss, L., Sainio, K., Pakarinen, P., & Poutanen, M. (2010). Hydroxysteroid (17β) Dehydrogenase 12 Is Essential for Mouse Organogenesis and Embryonic Survival. Endocrinology, 151(4), 1893–1901. 10.1210/en.2009-0929

Rison, S., Redfern, O., Dostal, I., Carvalho, C., Mathur, R., Raisi-Estabragh, Z., & Robson, J. (2023). Inequities in hypertension management: Observational cross-sectional study in North East London using electronic health records. British Journal of General Practice, 73(736), e798– e806. 10.3399/BJGP.2023.0077

Sabatti, C., Service, S. K., Hartikainen, A.-L., Pouta, A., Ripatti, S., Brodsky, J., Jones, C. G., Zaitlen, N. A., Varilo, T., Kaakinen, M., Sovio, U., Ruokonen, A., Laitinen, J., Jakkula, E., Coin, L., Hoggart, C., Collins, A., Turunen, H., Gabriel, S., … Peltonen, L. (2009). Genome-wide association analysis of metabolic traits in a birth cohort from a founder population. Nature Genetics, 41(1), 35–46. 10.1038/ng.271

Sanderson, E., Davey Smith, G., Windmeijer, F., & Bowden, J. (2019). An examination of multivariable Mendelian randomization in the single-sample and two-sample summary data settings. International Journal of Epidemiology, 48(3), 713–727. 10.1093/ije/dyy262

Sanderson, E., Glymour, M. M., Holmes, M. V., Kang, H., Morrison, J., Munafò, M. R., Palmer, T., Schooling, C. M., Wallace, C., Zhao, Q., & Davey Smith, G. (2022). Mendelian randomization. Nature Reviews Methods Primers, 2(1), 6. 10.1038/s43586-021-00092-5

Sanderson, E., Richardson, T. G., Hemani, G., & Davey Smith, G. (2021). The use of negative control outcomes in Mendelian randomization to detect potential population stratification. International Journal of Epidemiology, 50(4), 1350–1361. 10.1093/ije/dyaa288

Sanderson, E., Spiller, W., & Bowden, J. (2021). Testing and correcting for weak and pleiotropic instruments in two-sample multivariable Mendelian randomization. Statistics in Medicine, 40(25), 5434–5452. 10.1002/sim.9133

Schoeler, T., Speed, D., Porcu, E., Pirastu, N., Pingault, J.-B., & Kutalik, Z. (2023). Participation bias in the UK Biobank distorts genetic associations and downstream analyses. Nature Human Behaviour, 7(7), 1216–1227. 10.1038/s41562-023-01579-9

Smith, D. K., Lennon, R. P., & Carlsgaard, P. B. (2020). Managing Hypertension Using Combination Therapy. 101(6), 341–349.

Surendran, P., Feofanova, E. V., Lahrouchi, N., Ntalla, I., Karthikeyan, S., Cook, J., Chen, L., Mifsud, B., Yao, C., Kraja, A. T., Cartwright, J. H., Hellwege, J. N., Giri, A., Tragante, V., Thorleifsson, G., Liu, D. J., Prins, B. P., Stewart, I. D., Cabrera, C. P., … Howson, J. M. M. (2020). Discovery of rare variants associated with blood pressure regulation through meta-analysis of 1.3 million individuals. Nature Genetics, 52(12), 1314–1332. 10.1038/s41588-020-00713-x

Tobin, M. D., Sheehan, N. A., Scurrah, K. J., & Burton, P. R. (2005). Adjusting for treatment effects in studies of quantitative traits: Antihypertensive therapy and systolic blood pressure. Statistics in Medicine, 24(19), 2911–2935. 10.1002/sim.2165

Vishnolia, K. K., Hoene, C., Tarhbalouti, K., Revenstorff, J., Aherrahrou, Z., & Erdmann, J. (2020). Studies in Zebrafish Demonstrate That CNNM2 and NT5C2 Are Most Likely the Causal Genes at the Blood Pressure-Associated Locus on Human Chromosome 10q24.32. Frontiers in Cardiovascular Medicine, 7. 10.3389/fcvm.2020.00135

Visscher, P. M., Wray, N. R., Zhang, Q., Sklar, P., McCarthy, M. I., Brown, M. A., & Yang, J. (2017). 10 Years of GWAS Discovery: Biology, Function, and Translation. American Journal of Human Genetics, 101(1), 5–22. 10.1016/j.ajhg.2017.06.005

Wain, L. V., Vaez, A., Jansen, R., Joehanes, R., van der Most, P. J., Erzurumluoglu, A. M., O’Reilly, P. F., Cabrera, C. P., Warren, H. R., Rose, L. M., Verwoert, G. C., Hottenga, J.-J., Strawbridge, R. J., Esko, T., Arking, D. E., Hwang, S.-J., Guo, X., Kutalik, Z., Trompet, S., … Ehret, G. B. (2017). Novel Blood Pressure Locus and Gene Discovery Using Genome-Wide Association Study and Expression Data Sets From Blood and the Kidney. Hypertension, 70(3), e4–e19. 10.1161/HYPERTENSIONAHA.117.09438

Walker, V., Harrison, S., Carter, A., Gill, D., Tzoulaki, I., & Davies, N. (2021). The consequences of adjustment, correction and selection in genome-wide association studies used for two-sample Mendelian randomization. Wellcome Open Research, 6, 103. 10.12688/wellcomeopenres.16752.1

Ward, C. J., Hogan, M. C., Rossetti, S., Walker, D., Sneddon, T., Wang, X., Kubly, V., Cunningham, J. M., Bacallao, R., Ishibashi, M., Milliner, D. S., Torres, V. E., & Harris, P. C. (2002). The gene mutated in autosomal recessive polycystic kidney disease encodes a large, receptor-like protein. Nature Genetics, 30(3), 259–269. 10.1038/ng833

Watanabe, S., Wei, F.-Y., & Tomizawa, K. (2013). Functional characterization of Cdkal1, a risk factor of type 2 diabetes, and the translational opportunities. *Drug Discovery Today: Disease Models*, Diabetes, 10(2), e65–e69. 10.1016/j.ddmod.2012.12.001

Weale, M. E. (2010). Quality Control for Genome-Wide Association Studies. In M. R. Barnes & G. Breen (Eds), Genetic Variation: Methods and Protocols (pp. 341–372). Humana Press. 10.1007/978-1-60327-367-1_19

Wu, Y., Byrne, E. M., Zheng, Z., Kemper, K. E., Yengo, L., Mallett, A. J., Yang, J., Visscher, P. M., & Wray, N. R. (2019). Genome-wide association study of medication-use and associated disease in the UK Biobank. Nature Communications, 10(1), 1891. 10.1038/s41467-019-09572-5

Yamazaki, K., Terao, C., Takahashi, A., Kamatani, Y., Matsuda, K., Asai, S., & Takahashi, Y. (2023). Genome-wide Association Studies Categorized by Class of Antihypertensive Drugs Reveal Complex Pathogenesis of Hypertension with Drug Resistance. Clinical Pharmacology & Therapeutics, 114(2), 393–403. 10.1002/cpt.2934

Yang, J., Lee, S. H., Goddard, M. E., & Visscher, P. M. (2011). GCTA: A Tool for Genome-wide Complex Trait Analysis. American Journal of Human Genetics, 88(1), 76–82. 10.1016/j.ajhg.2010.11.011

