## Supplementary Material 1 for "Adjusting for medication use in GWAS and its impact on Mendelian randomization analyses: an example of systolic blood pressure in UK Biobank"

### **Supplementary Figure 1**. Histograms of observed and adjusted systolic blood pressure

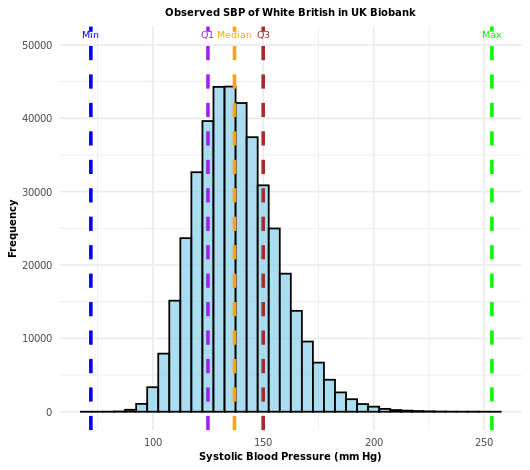

a) Unadjusted

b) Add constant +10mmHg

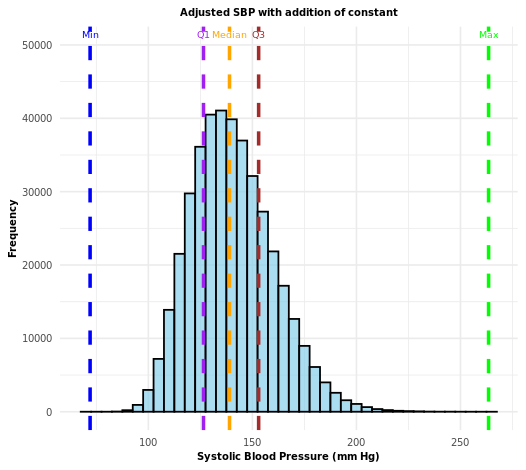

c) Add class-specific constant

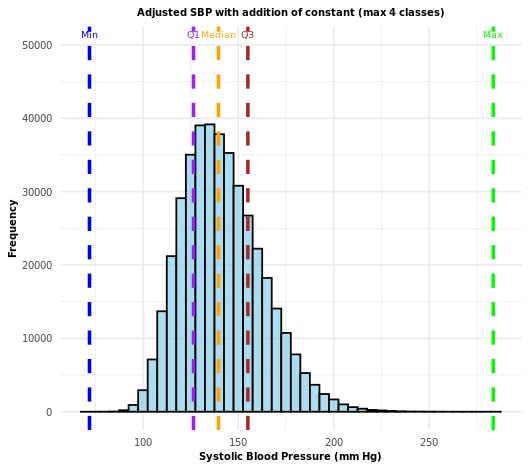

d) Censored normal regression

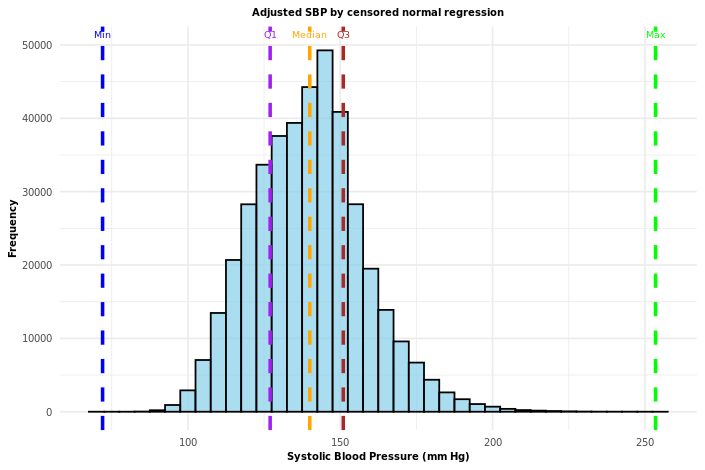

e) Restrict to untreated

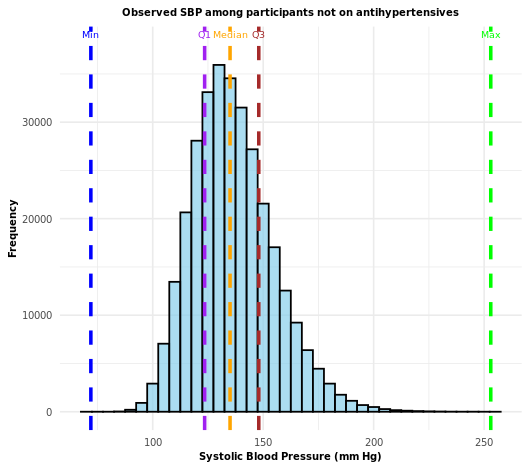

f) Restrict to treated

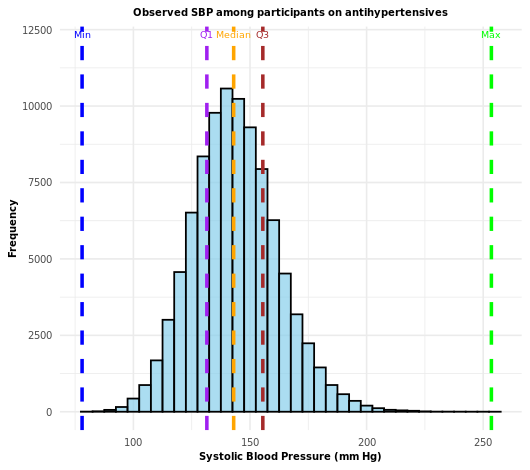

g) Restrict to 38-49 years

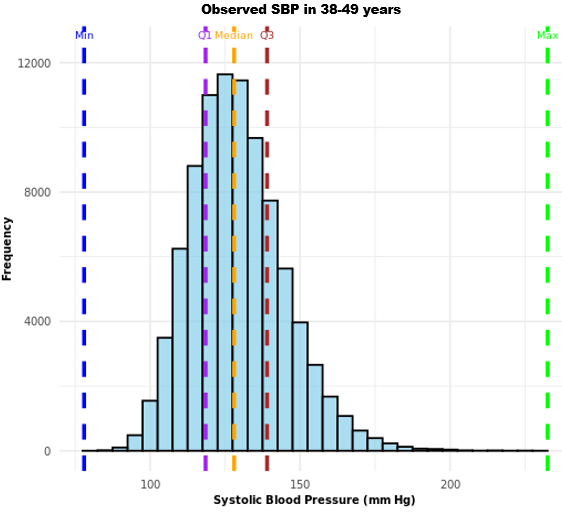

h) Restrict to 50-59 years

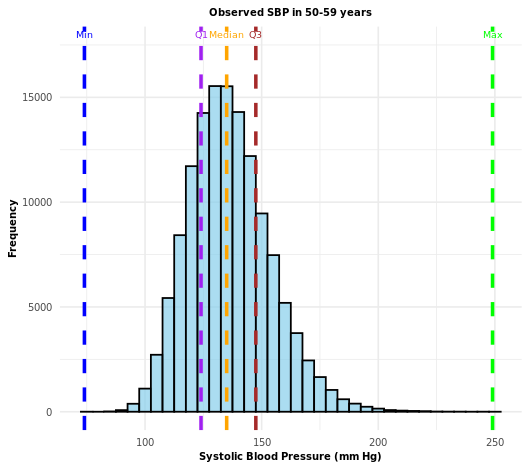

i) Restrict to 60-73 years

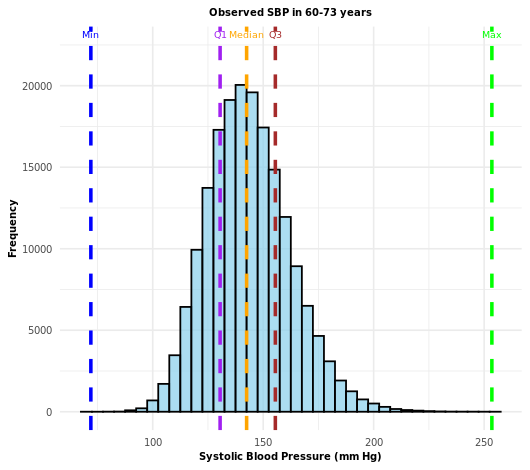

SBP: systolic blood pressure

SBP values of the cohort were plotted on histograms to check for the normality assumption of linearity. The y-axis shows the frequency counts (number of participants), whereas the x-axis shows the SBP values in millimetres of mercury. Histogram (a) shows the observed unadjusted SBP values, whereas Histograms (b) to (i) show the distribution of SBP values after various adjustments for medication use have been applied. The vertical dotted lines indicate the minimum, first quartile, median, third quartile and maximum values respectively.

### **Supplementary Figure 2**. Manhattan and QQ plots for GWAS of systolic blood pressure (unadjusted)

a) Manhattan Plot

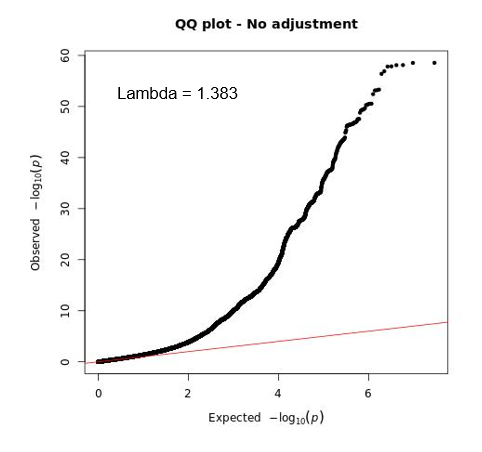

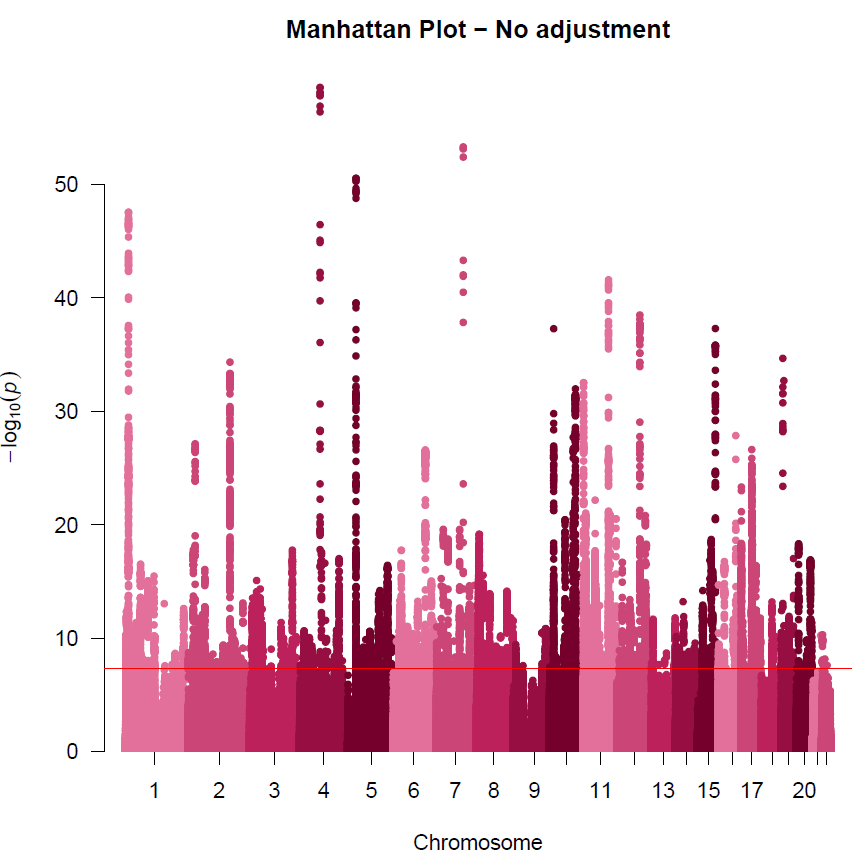

b) Quantile-Quantile Plot

Association testing (a) Manhattan plot and (b) QQ plot for systolic blood pressure (unadjusted) using fastGWA linear mixed models.

### **Supplementary Figure 3**. Manhattan and QQ plots for GWAS of systolic blood pressure (add constant +10 mmHg)

a) Manhattan Plot

b) Quantile-Quantile Plot

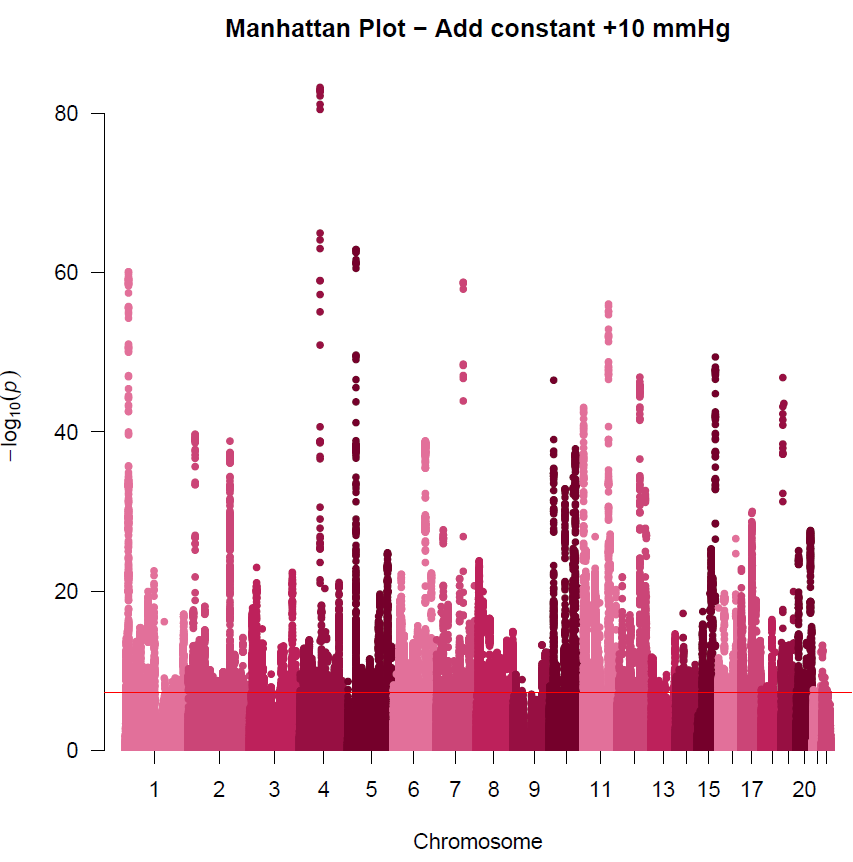

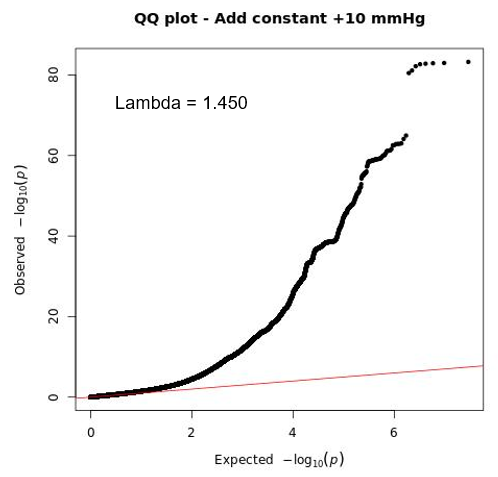

Association testing (a) Manhattan plot and (b) QQ plot for systolic blood pressure (add constant +10 mmHg) using fastGWA linear mixed models.

### **Supplementary Figure 4**. Manhattan and QQ plots for GWAS of systolic blood pressure (add class-specific constant)

a) Manhattan Plot

b) Quantile-Quantile Plot

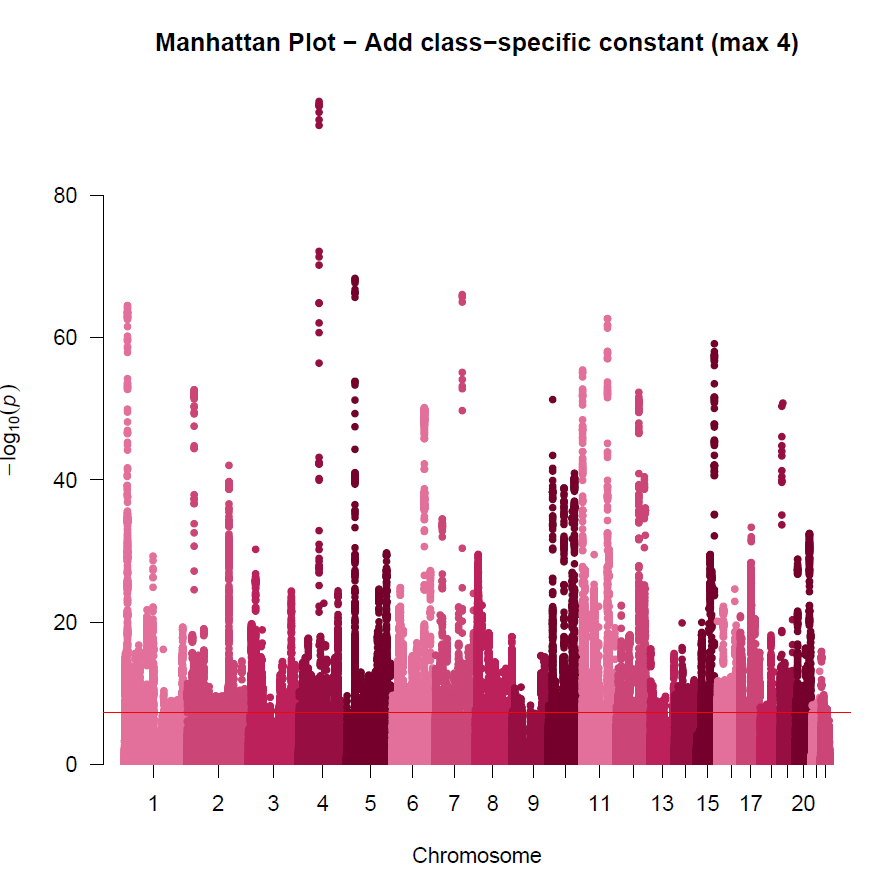

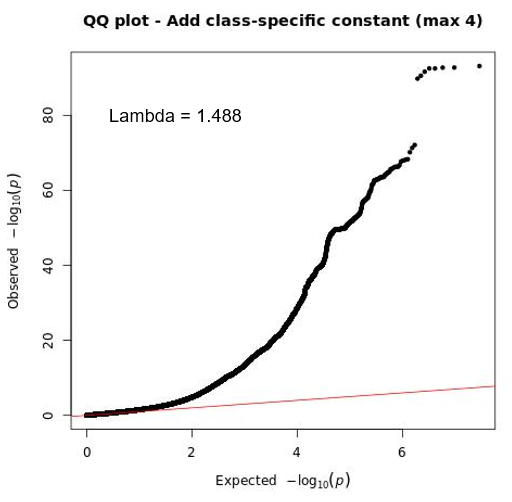

Association testing (a) Manhattan plot and (b) QQ plot for systolic blood pressure (add class-specific constant) using fastGWA linear mixed models.

### **Supplementary Figure 5**. Manhattan and QQ plots for GWAS of systolic blood pressure (censored normal regression)

a) Manhattan Plot

b) Quantile-Quantile Plot

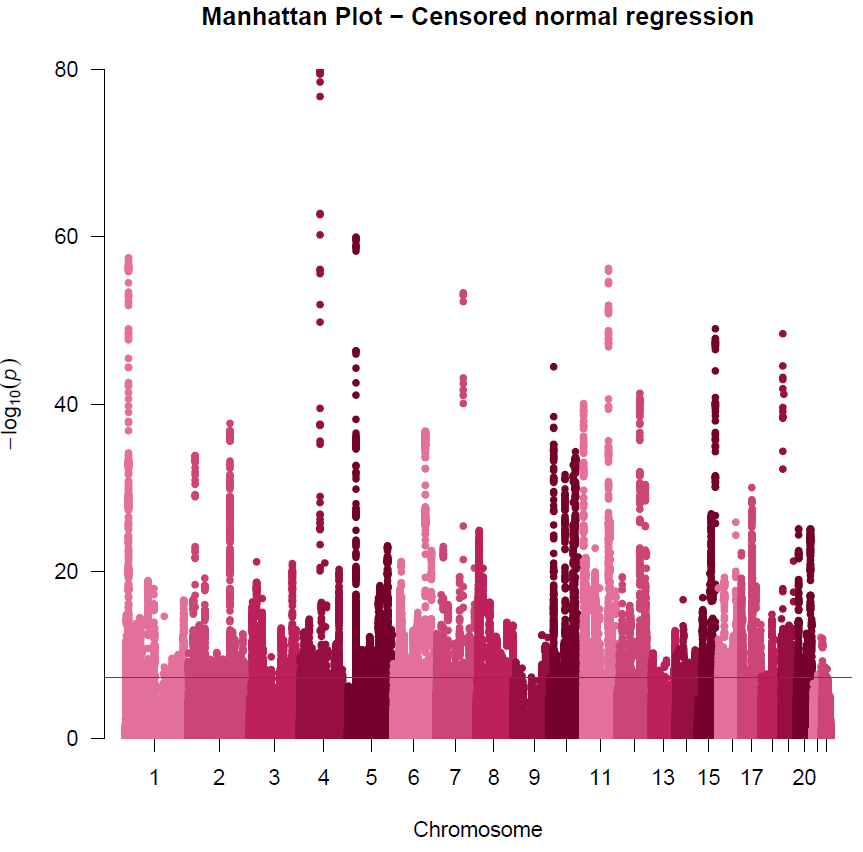

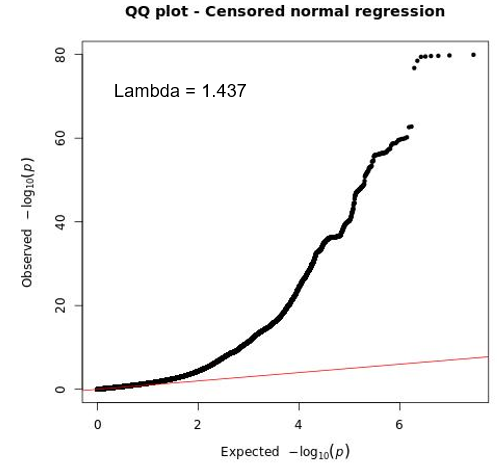

Association testing (a) Manhattan plot and (b) QQ plot for systolic blood pressure (censored normal regression) using fastGWA linear mixed models.

### **Supplementary Figure 6**. Manhattan and QQ plots for GWAS of systolic blood pressure (restrict to untreated)

a) Manhattan Plot

b) Quantile-Quantile Plot

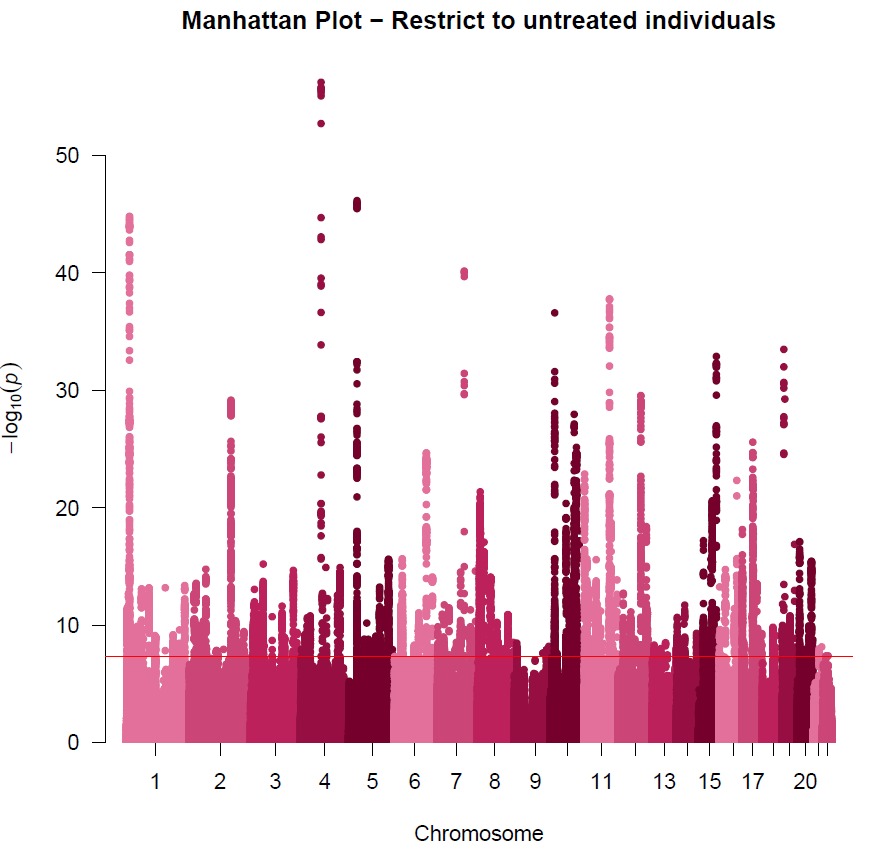

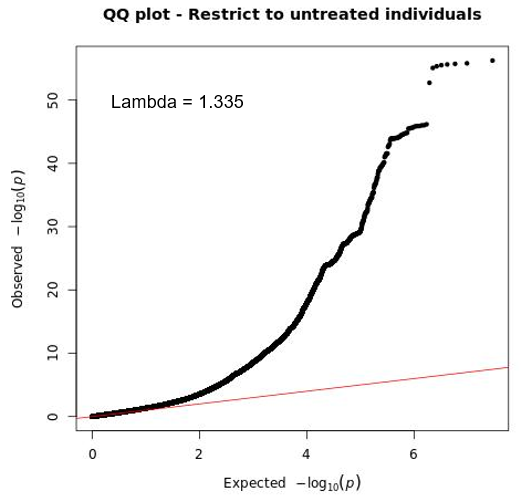

Association testing (a) Manhattan plot and (b) QQ plot for systolic blood pressure (restrict to untreated individuals) using fastGWA linear mixed models.

### **Supplementary Figure 7**. Manhattan and QQ plots for GWAS of systolic blood pressure (restrict to treated)

a) Manhattan Plot

b) Quantile-Quantile Plot

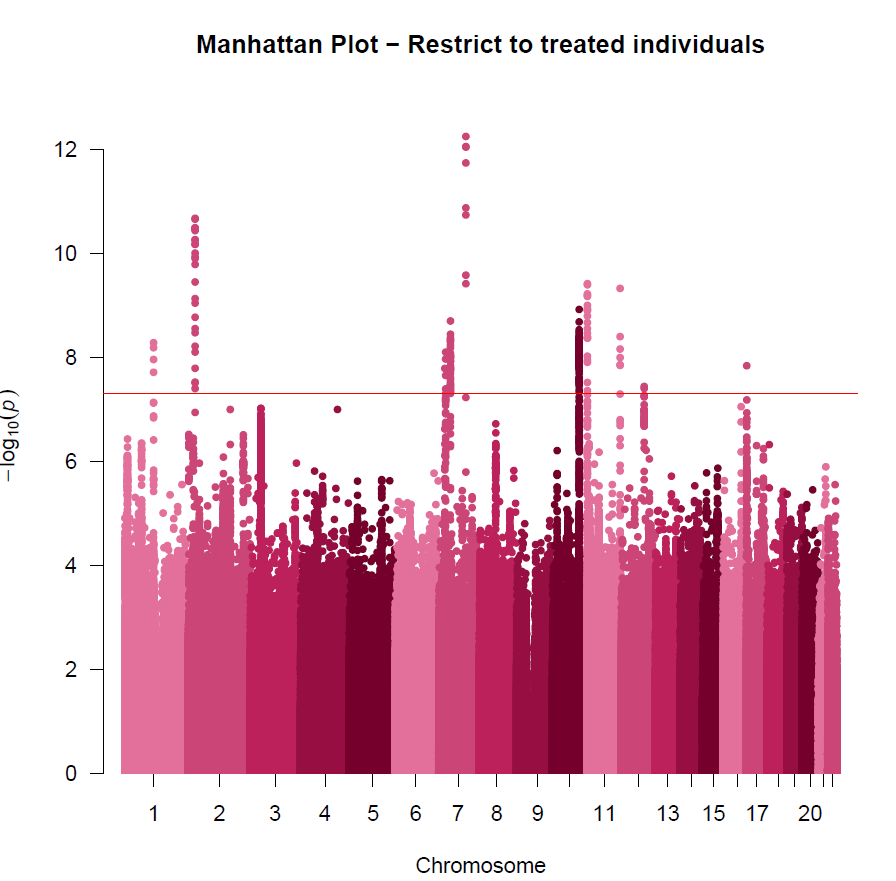

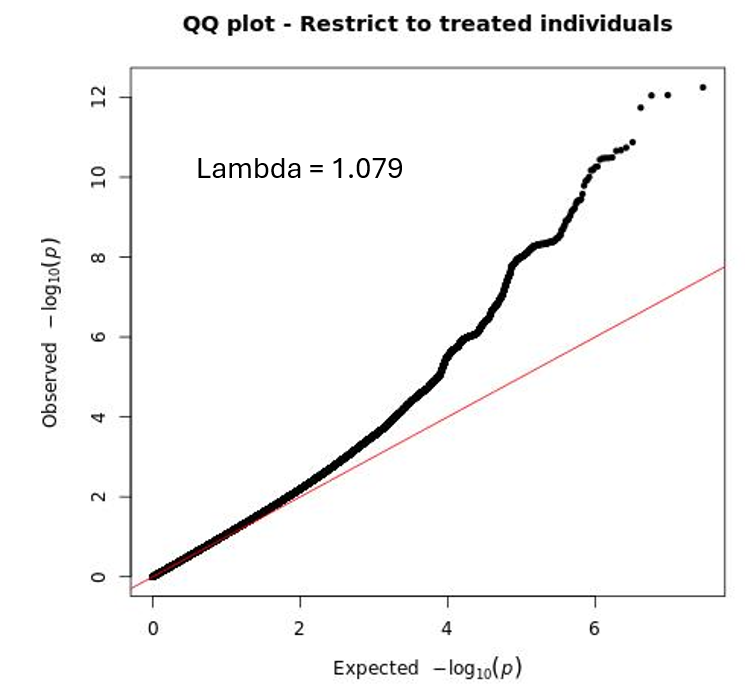

Association testing (a) Manhattan plot and (b) QQ plot for systolic blood pressure (restrict to treated individuals) using fastGWA linear mixed models.

### **Supplementary Figure 8**. Manhattan and QQ plots for GWAS of systolic blood pressure (restrict to 38-49 years)

a) Manhattan Plot

b) Quantile-Quantile Plot

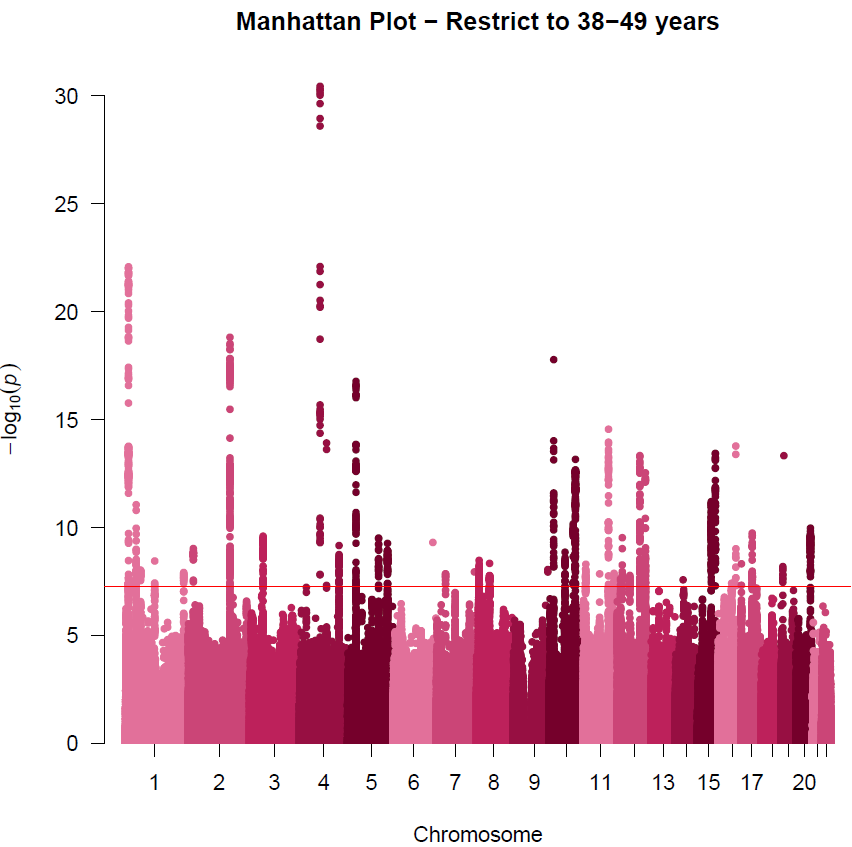

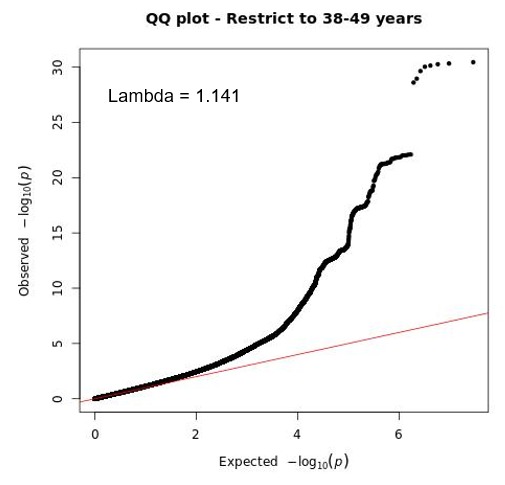

Association testing (a) Manhattan plot and (b) QQ plot for systolic blood pressure (restrict to 38-49 years) using fastGWA linear mixed models.

### **Supplementary Figure 9**. Manhattan and QQ plots for GWAS of systolic blood pressure (restrict to 50-59 years)

a) Manhattan Plot

b) Quantile-Quantile Plot

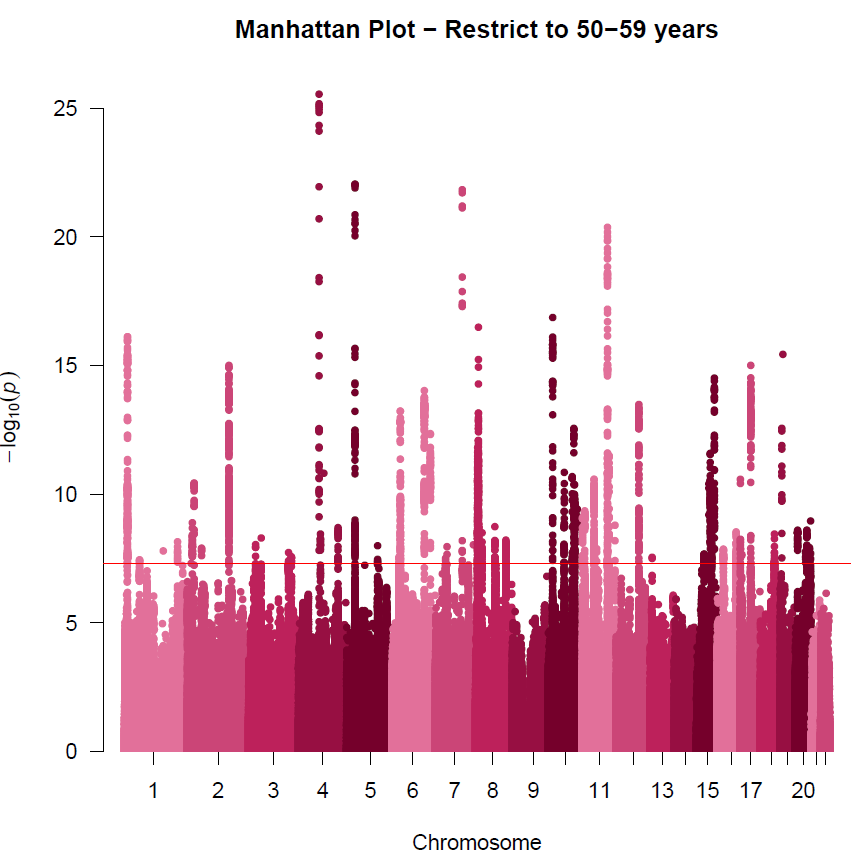

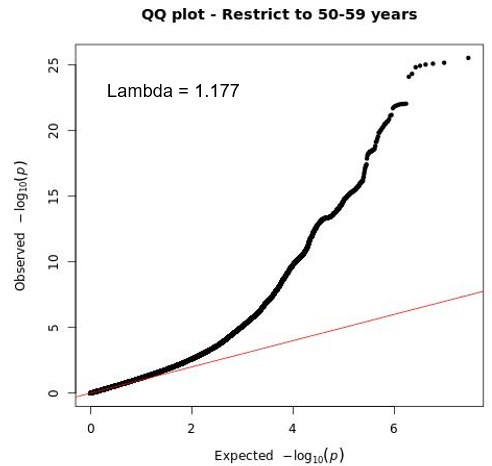

Association testing (a) Manhattan plot and (b) QQ plot for systolic blood pressure (restrict to 50-59 years) using fastGWA linear mixed models.

### **Supplementary Figure 10**. Manhattan and QQ plots for GWAS of systolic blood pressure (restrict to 60-73 years)

a) Manhattan Plot

b) Quantile-Quantile Plot

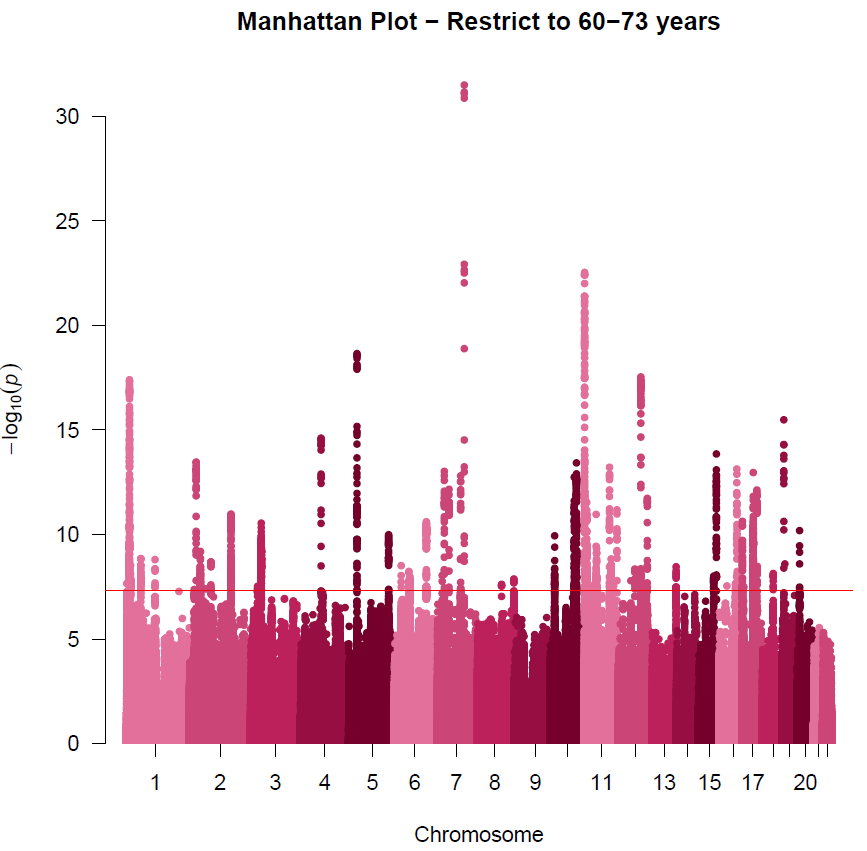

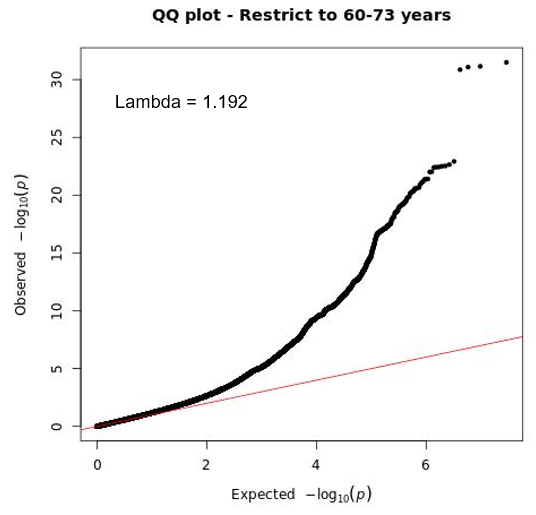

Association testing (a) Manhattan plot and (b) QQ plot for systolic blood pressure (restrict to 60-73 years) using fastGWA linear mixed models.

### **Supplementary Figure 11**. Manhattan and QQ plots for GWAS of systolic blood pressure with binary treatment status

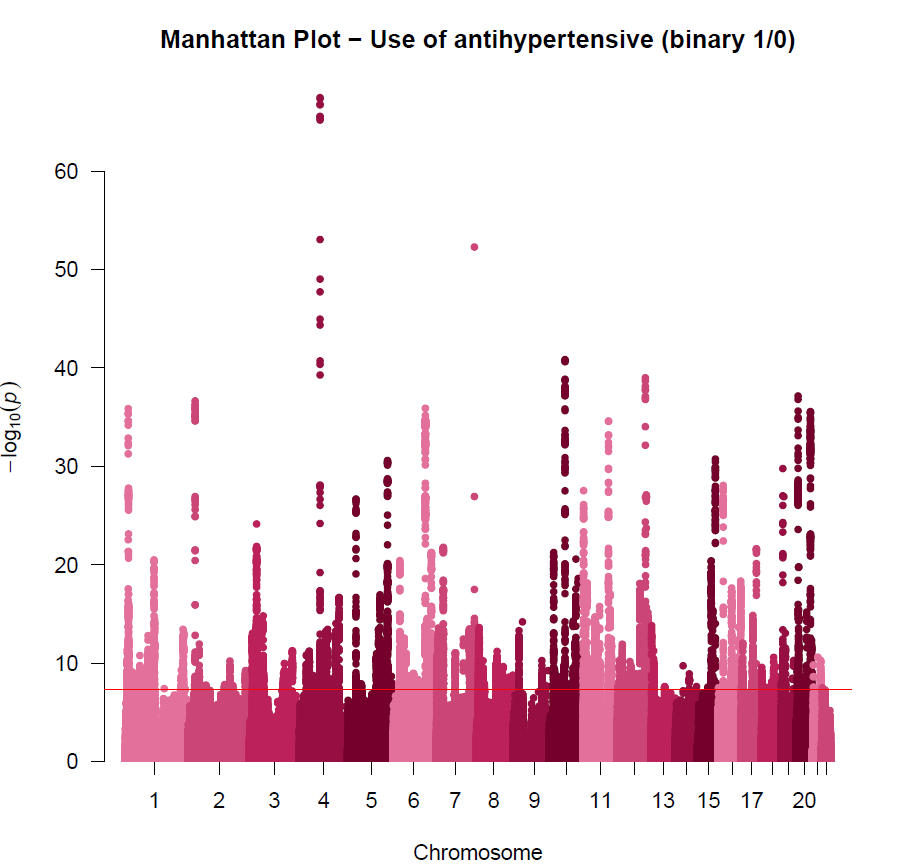

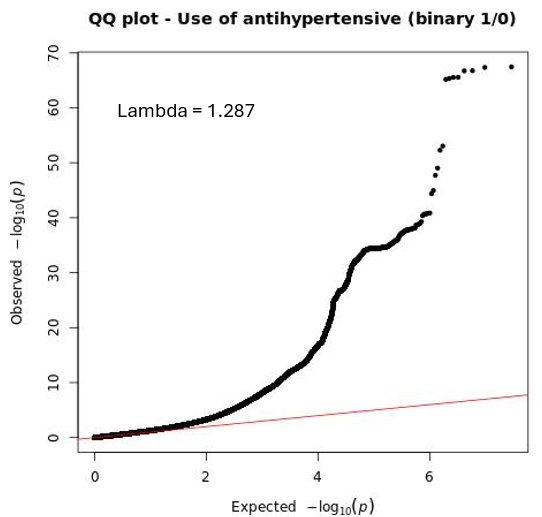

a) Manhattan Plot

b) Quantile-Quantile Plot

Association testing (a) Manhattan plot and (b) QQ plot for use of antihypertensive (binary trait) using fastGWA generalised linear mixed model.

### **Supplementary Figure 12**. Hudson plot of unadjusted versus add constant +10mmHg systolic blood pressure GWAS

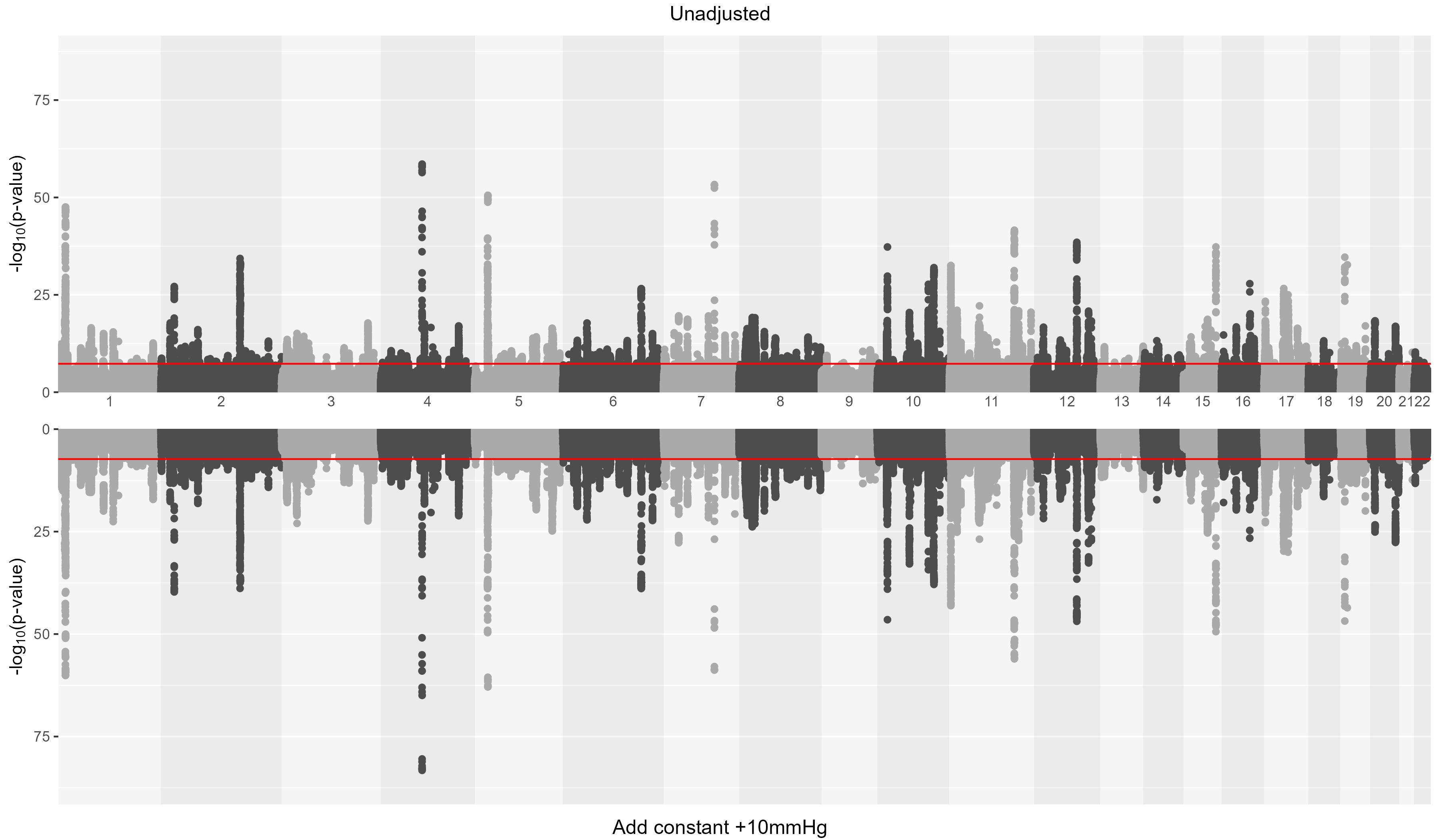

Each point represents a genetic variant, plotted by chromosomal position against −log₁₀(P). The top track shows the unadjusted analysis and the bottom track the analysis after adding +10 mmHg to the measured systolic blood pressure of individuals on antihypertensive medication. The horizontal red line marks the genome-wide significance threshold (P = 5×10⁻⁸). Analyses were conducted in fastGWA.

### **Supplementary Figure 13**. Hudson plot of unadjusted versus add class-specific constant systolic blood pressure GWAS

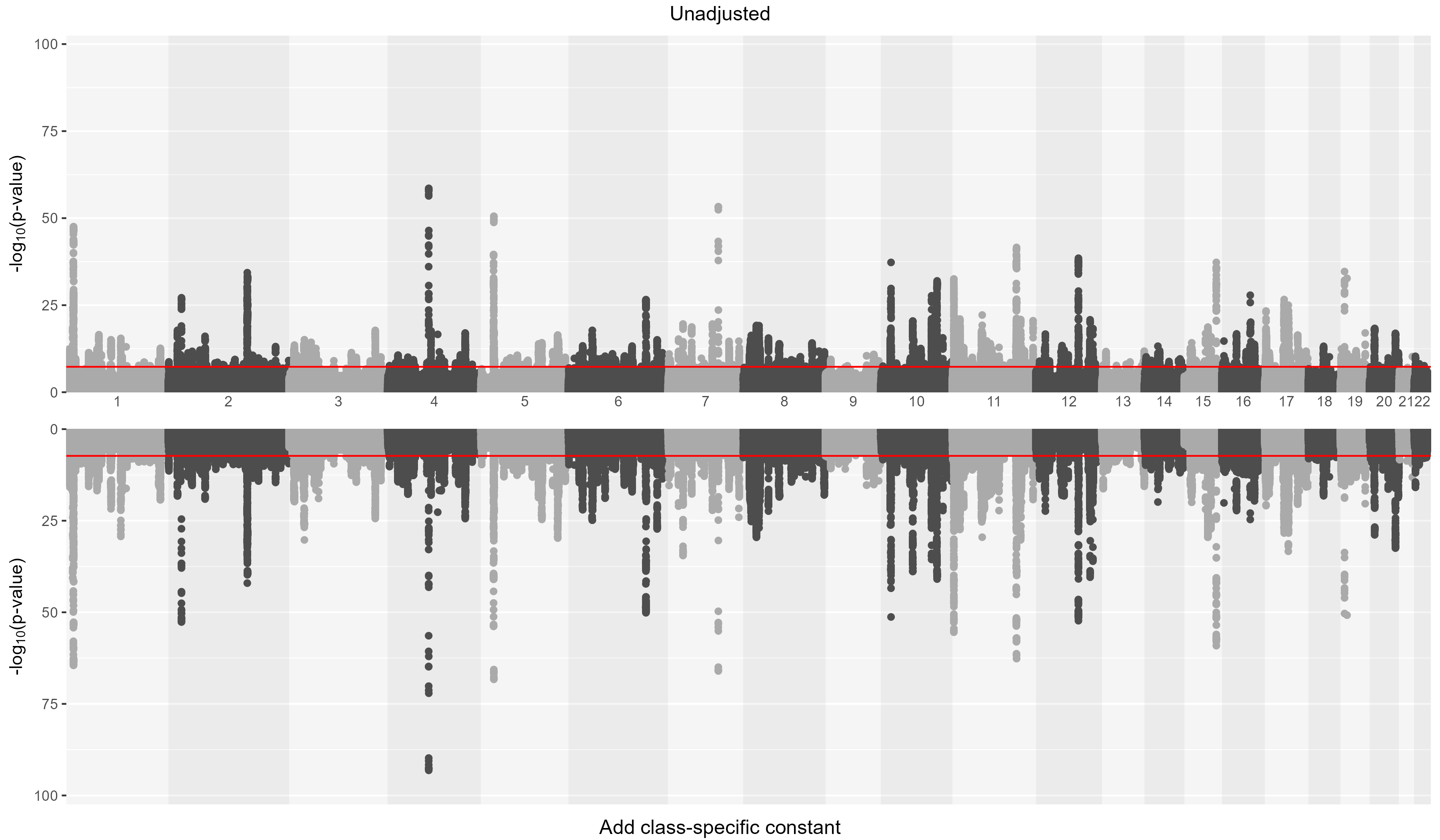

Each point represents a genetic variant, plotted by chromosomal position against −log₁₀(P). The top track shows the unadjusted analysis and the bottom track the analysis after adding class-specific constants to the measured systolic blood pressure of individuals on antihypertensive medication. The horizontal red line marks the genome-wide significance threshold (P = 5×10⁻⁸). Analyses were conducted in fastGWA.

### **Supplementary Figure 14**. Hudson plot of unadjusted versus censored normal regression systolic blood pressure GWAS

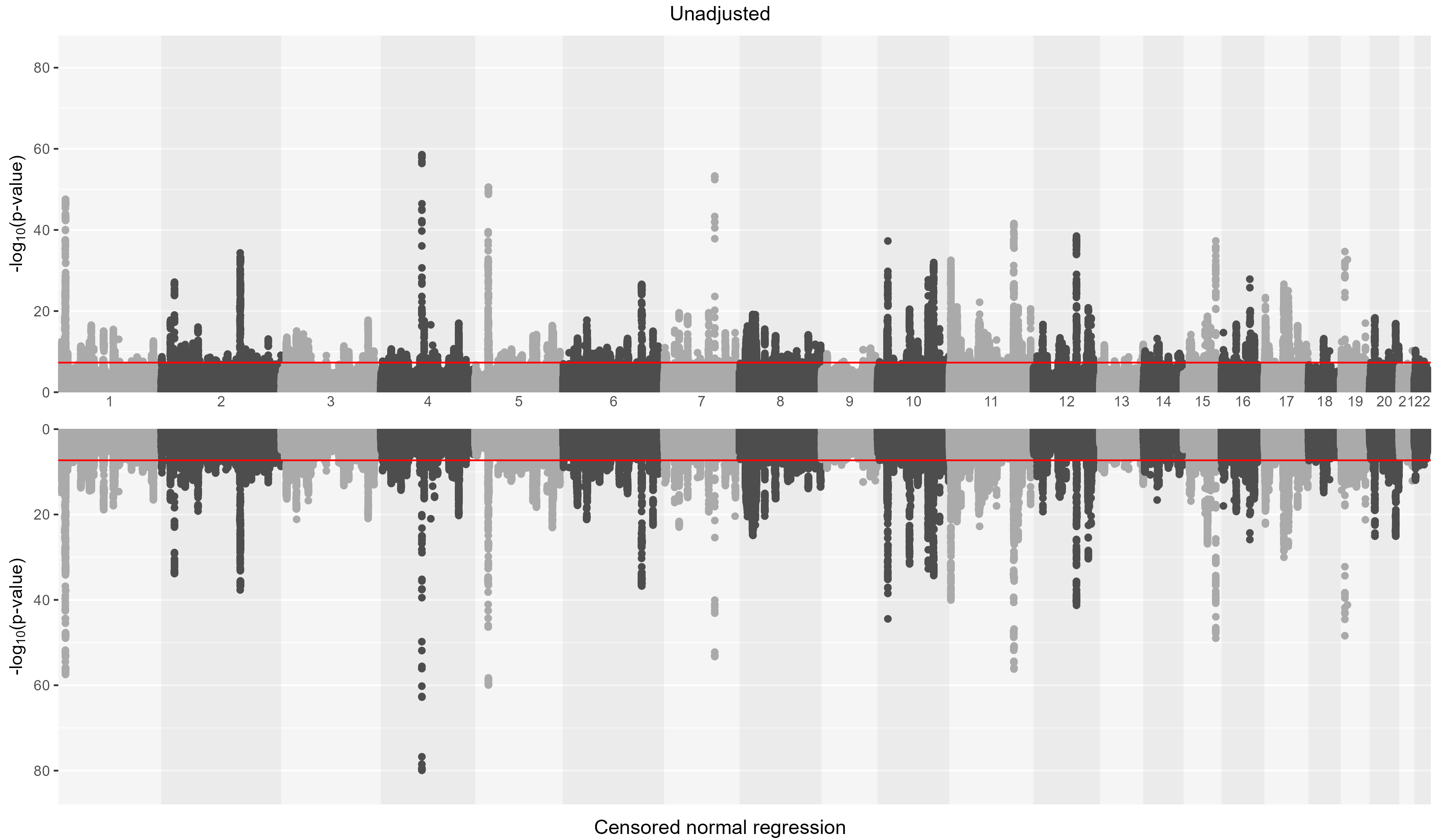

Each point represents a genetic variant, plotted by chromosomal position against −log₁₀(P). The top track shows the unadjusted analysis and the bottom track the analysis after medication adjustment by censored normal regression. The horizontal red line marks the genome-wide significance threshold (P = 5×10⁻⁸). Analyses were conducted in fastGWA.

### **Supplementary Figure 15**. Hudson plot of unadjusted versus restrict to untreated systolic blood pressure GWAS

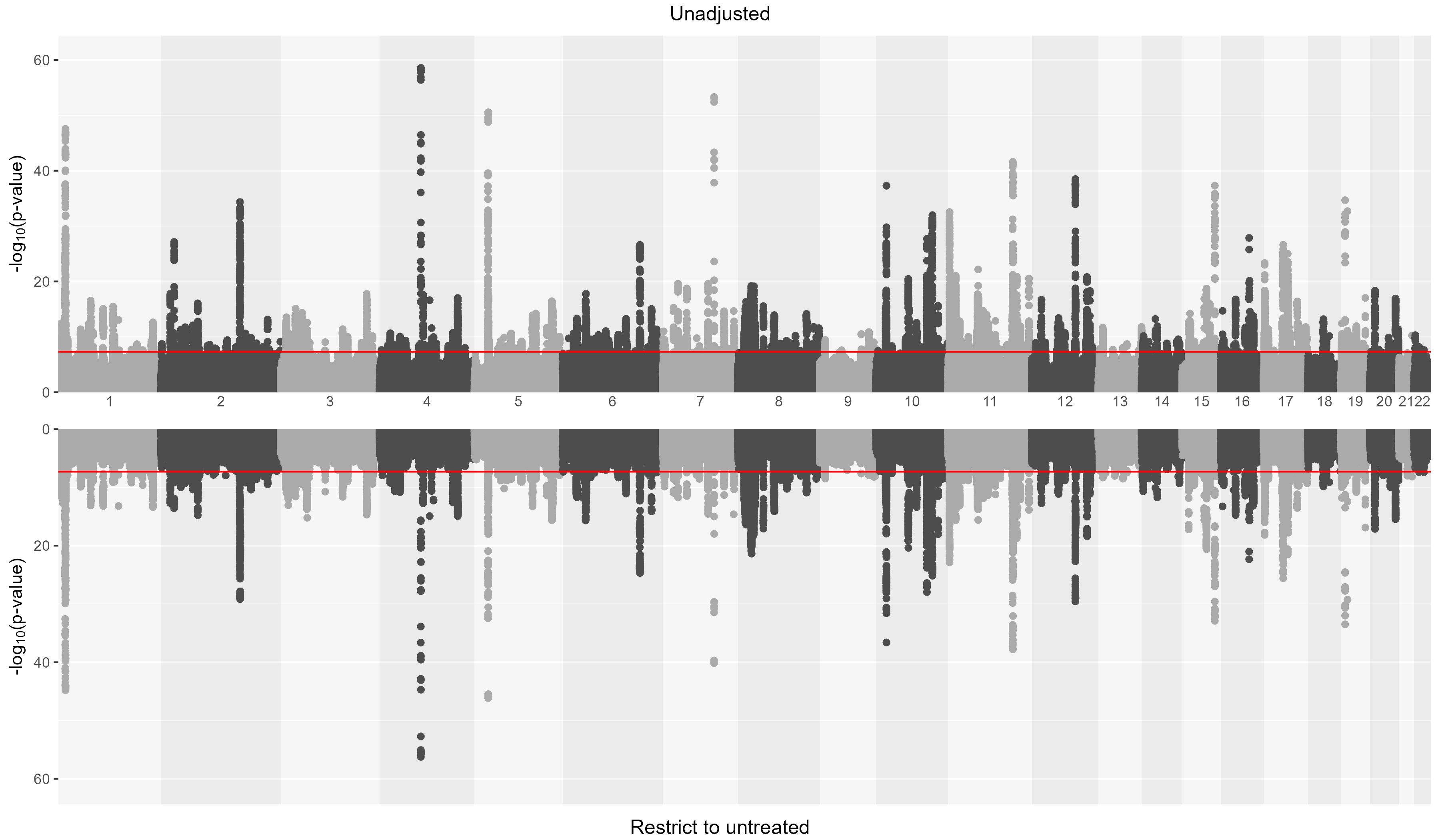

Each point represents a genetic variant, plotted by chromosomal position against −log₁₀(P). The top track shows the unadjusted analysis and the bottom track the analysis after medication adjustment by restriction to untreated individuals. The horizontal red line marks the genome-wide significance threshold (P = 5×10⁻⁸). Analyses were conducted in fastGWA.

### **Supplementary Figure 16**. Hudson plot of unadjusted versus restrict to 38-49 years systolic blood pressure GWAS

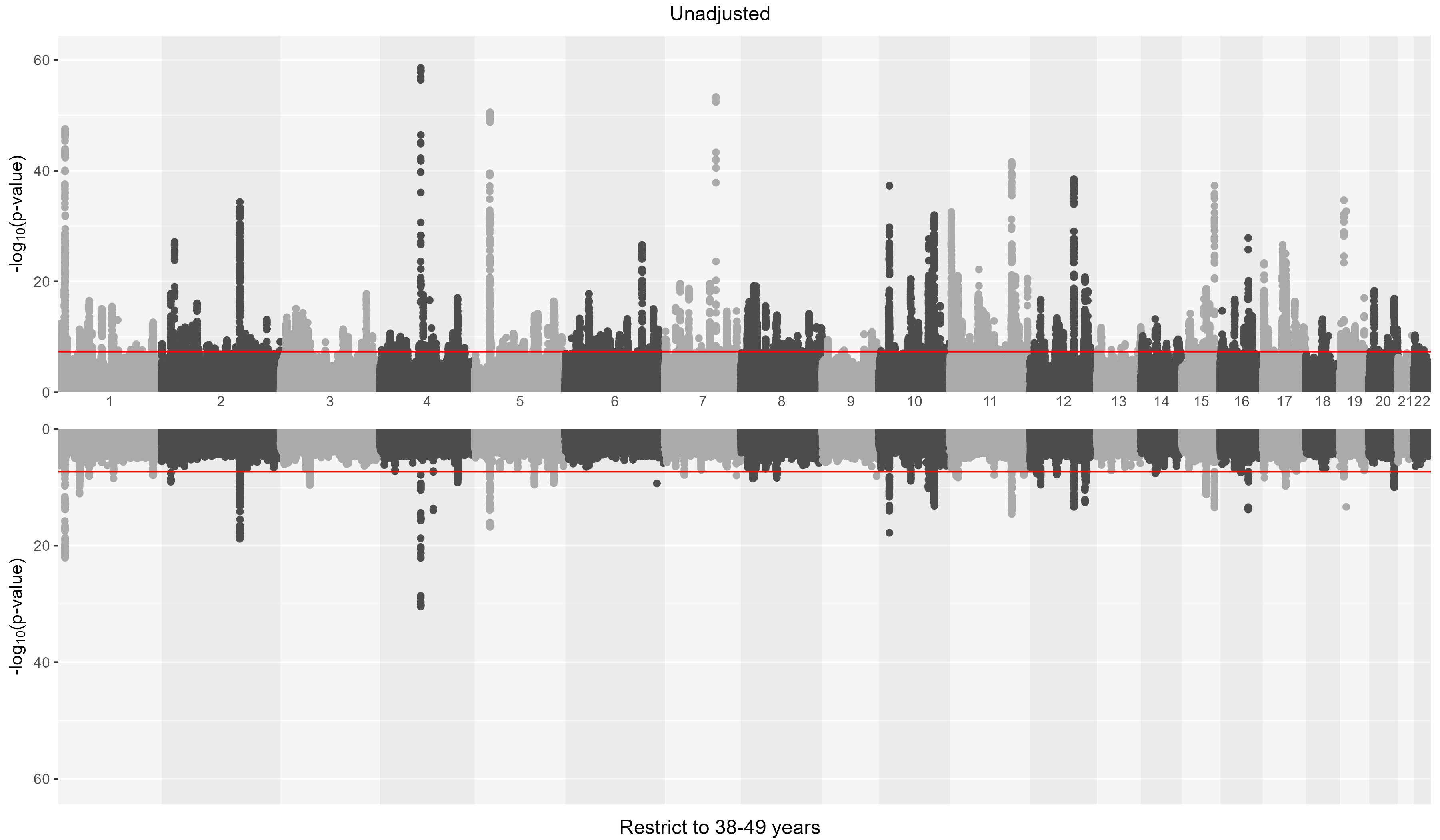

Each point represents a genetic variant, plotted by chromosomal position against −log₁₀(P). The top track shows the unadjusted analysis and the bottom track the analysis after medication adjustment by restriction to individuals aged 38-49 years. The horizontal red line marks the genome-wide significance threshold (P = 5×10⁻⁸). Analyses were conducted in fastGWA.

### **Supplementary Figure 17**. Hudson plot of unadjusted versus restrict to 50-59 years systolic blood pressure GWAS

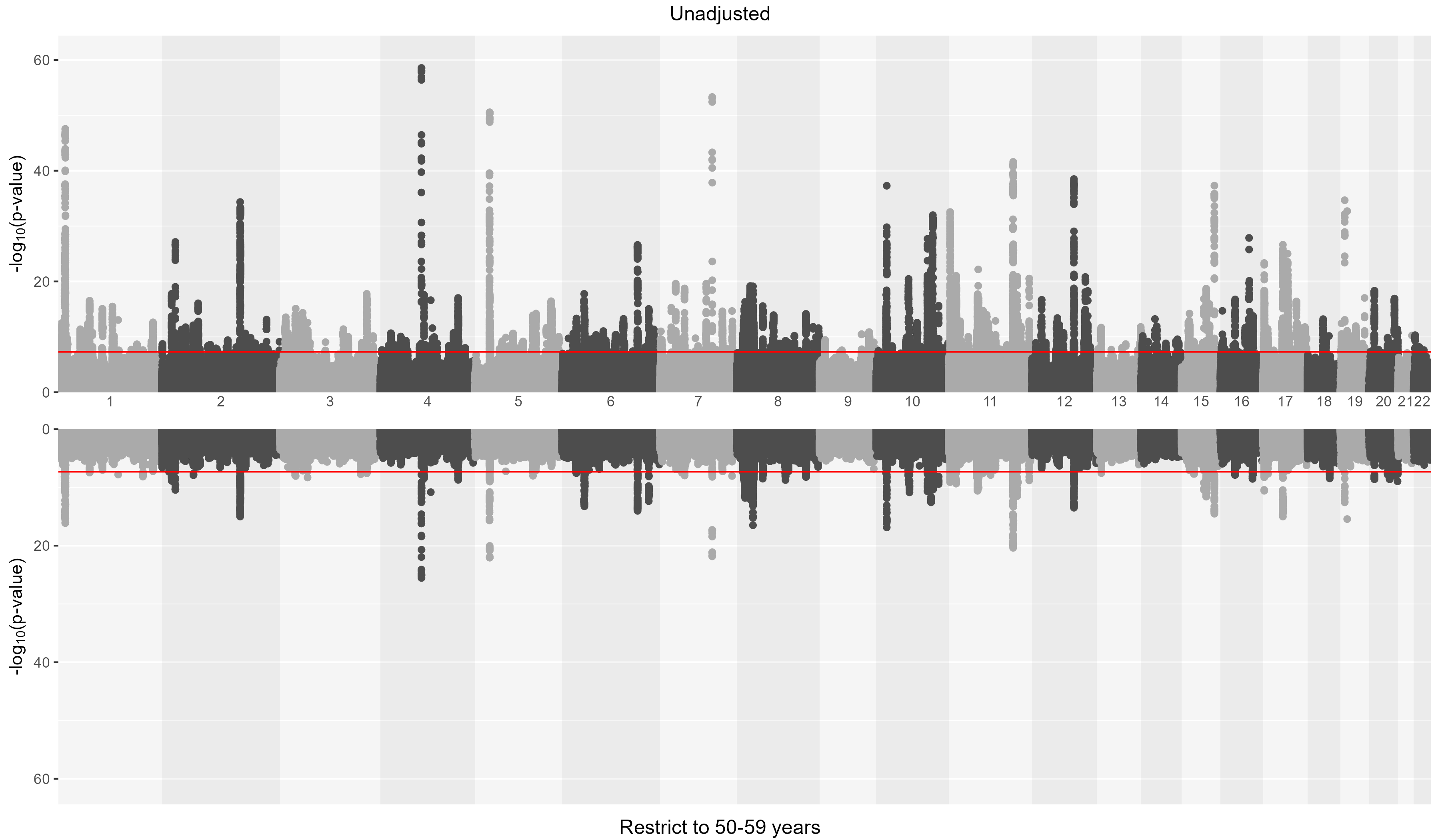

Each point represents a genetic variant, plotted by chromosomal position against −log₁₀(P). The top track shows the unadjusted analysis and the bottom track the analysis of restriction to individuals aged 50-59 years. The horizontal red line marks the genome-wide significance threshold (P = 5×10⁻⁸). Analyses were conducted in fastGWA.

### **Supplementary Figure 18**. Hudson plot of unadjusted versus restrict to 60-73 years systolic blood pressure GWAS

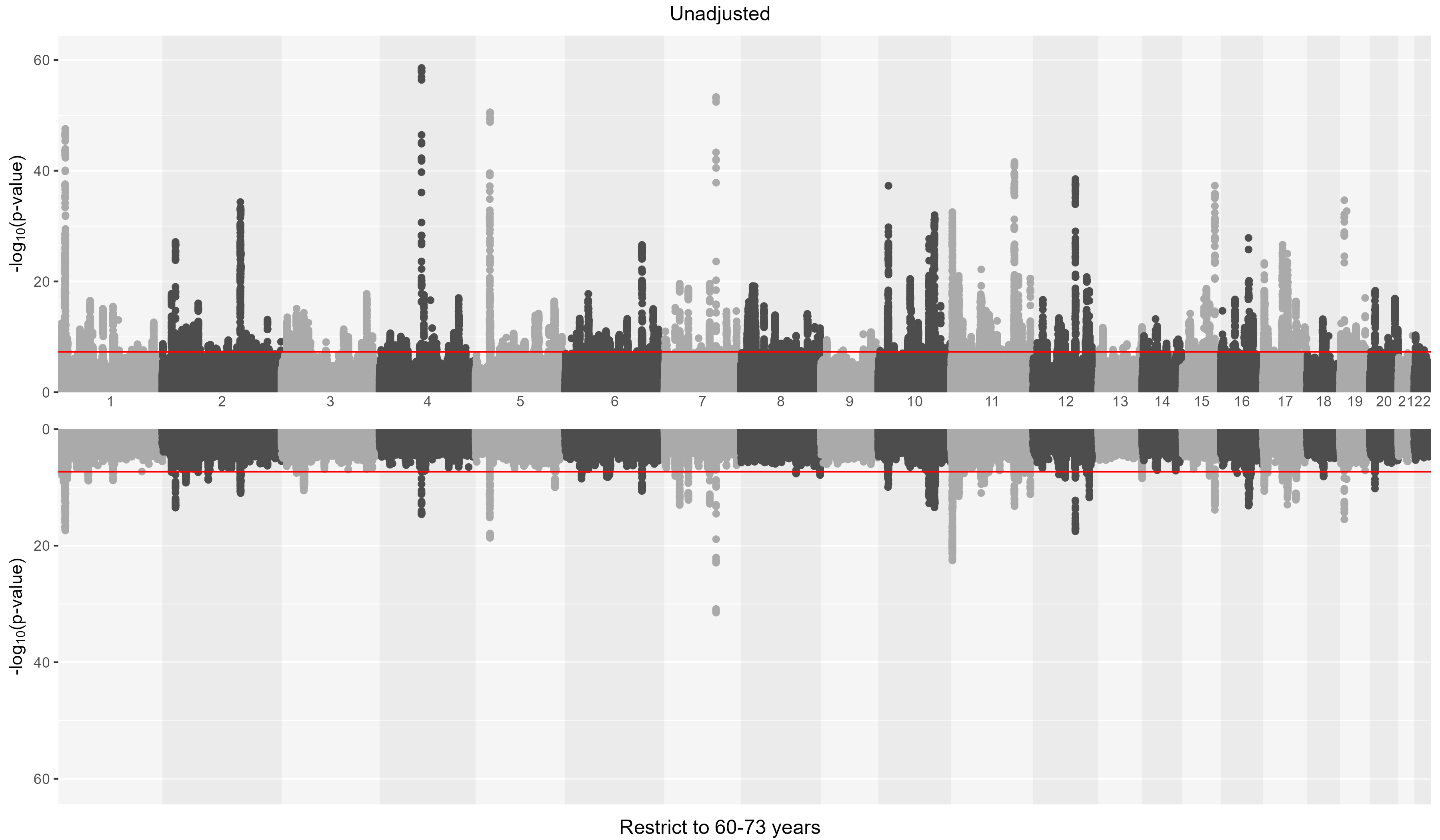

Each point represents a genetic variant, plotted by chromosomal position against −log₁₀(P). The top track shows the unadjusted analysis and the bottom track the analysis of restriction to individuals aged 60-73 years. The horizontal red line marks the genome-wide significance threshold (P = 5×10⁻⁸). Analyses were conducted in fastGWA.

### **Supplementary Figure 19**. Hudson plot of add constant +10mmHg versus add class-specific constant systolic blood pressure GWAS

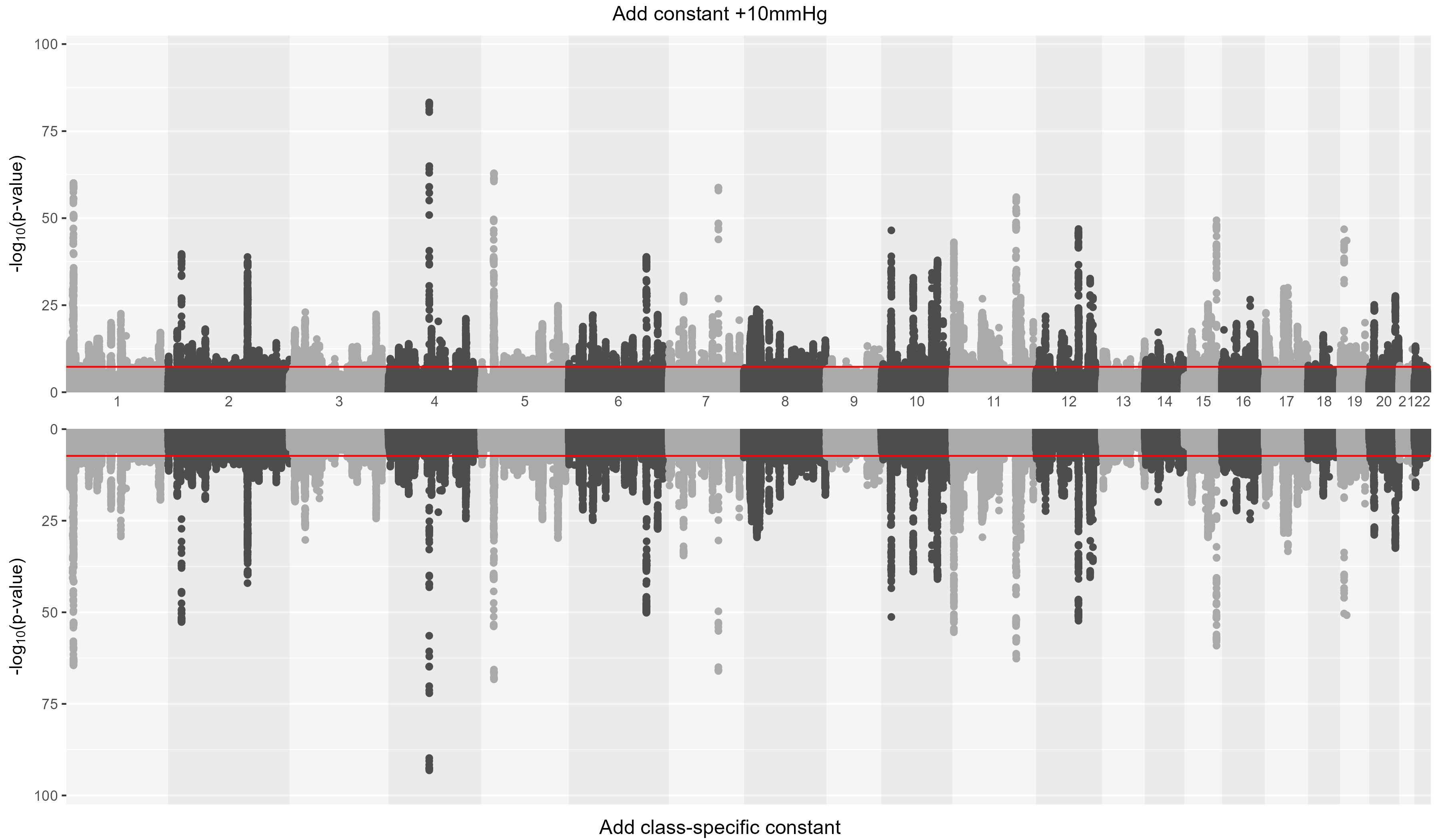

Each point represents a genetic variant, plotted by chromosomal position against −log₁₀(P). The top track shows the medication adjustment by adding constant of +10mmHg and the bottom track shows the analysis after medication adjustment by adding class-specific constants. The horizontal red line marks the genome-wide significance threshold (P = 5×10⁻⁸). Analyses were conducted in fastGWA.

### **Supplementary Figure 20**. Hudson plot of restrict to untreated versus restrict to treated systolic blood pressure GWAS

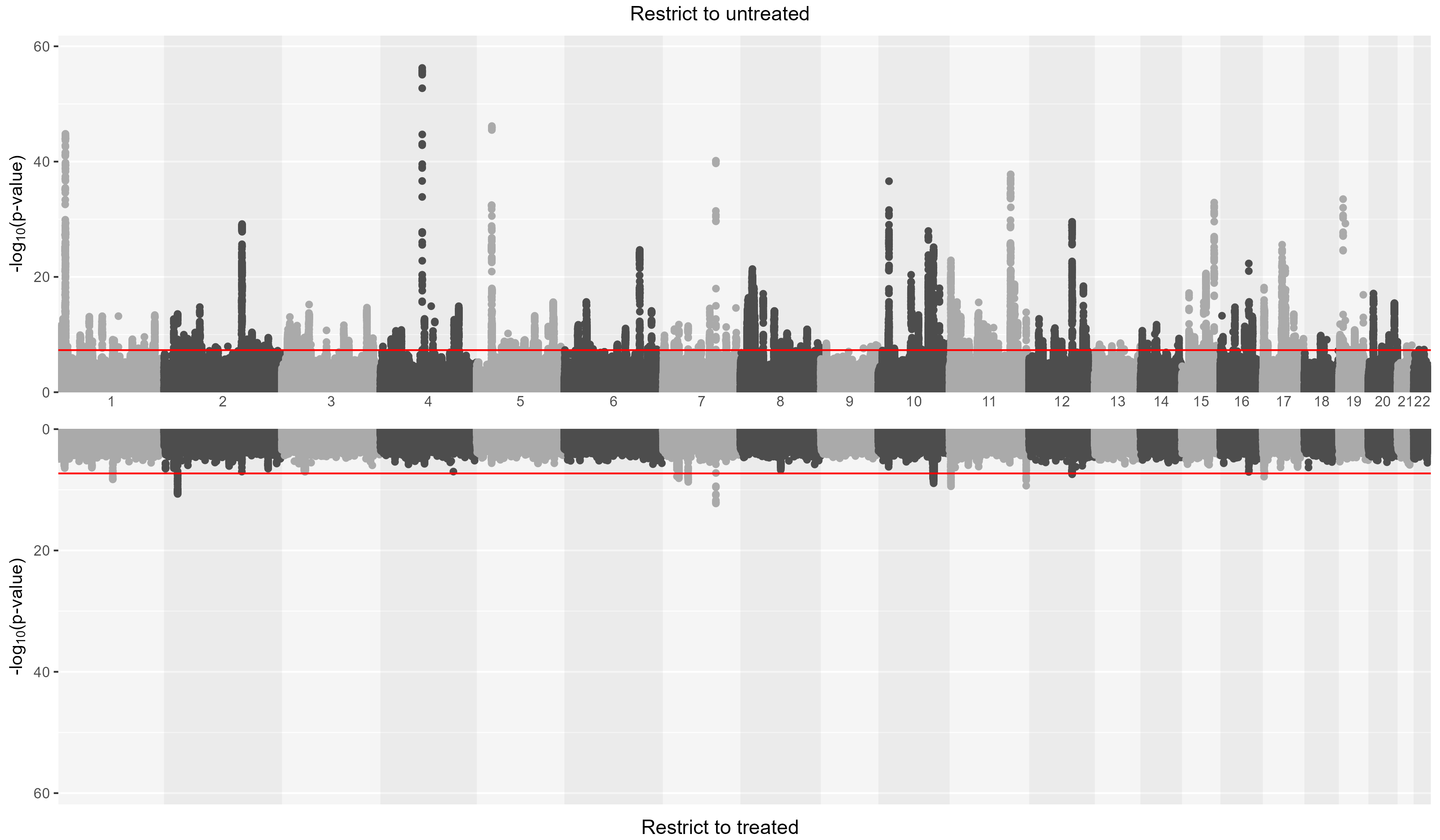

Each point represents a genetic variant, plotted by chromosomal position against −log₁₀(P). The top track shows the analysis restricted to untreated (with no direct adjustment to observed SBP values), and the bottom track shows the analysis restricted to treated. The horizontal red line marks the genome-wide significance threshold (P = 5×10⁻⁸). Analyses were conducted in fastGWA.

### **Supplementary Figure 21**. Venn diagrams of unclumped genetic variants from unadjusted (clumped reference) versus medication-adjusted GWAS

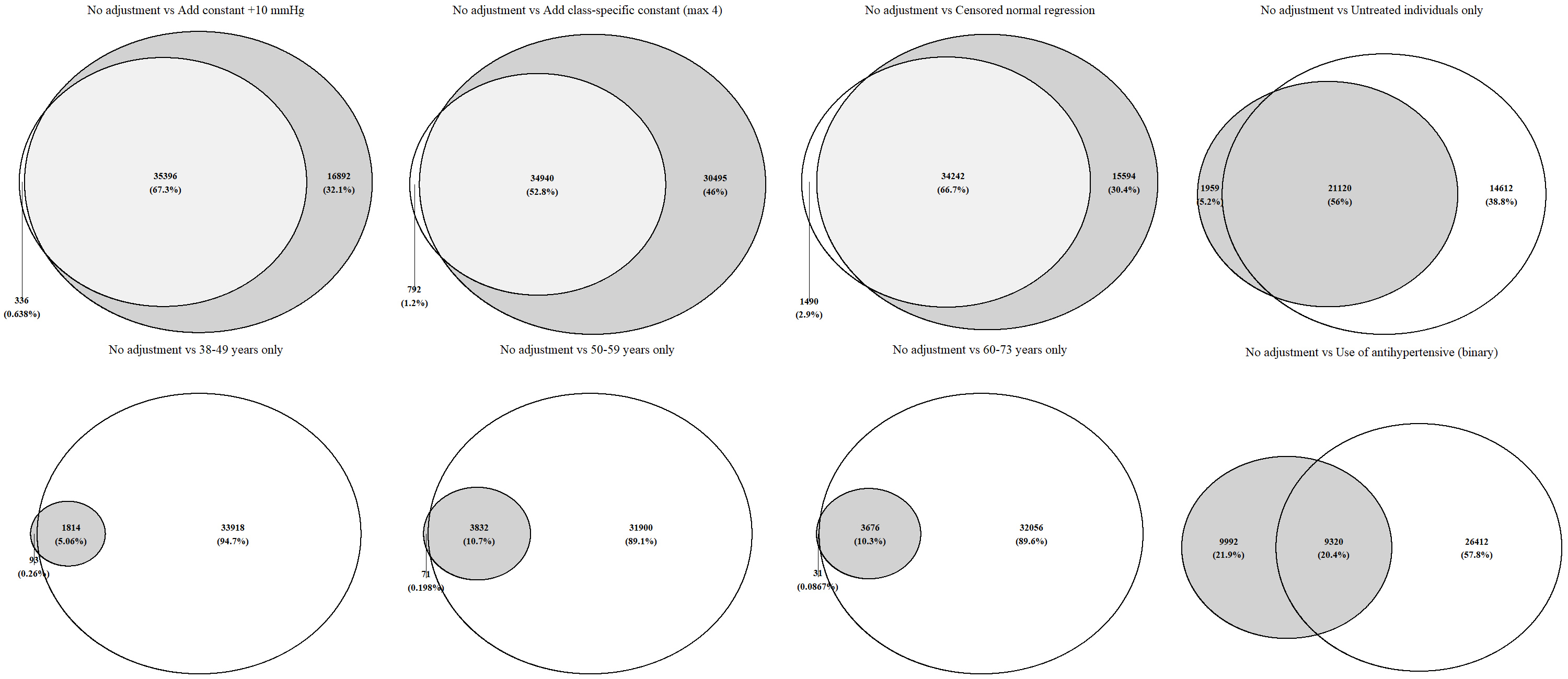

Unclumped top hits (p<5x10^-8^) from each SBP GWAS with medication adjustment (shaded) versus ‘no adjustment’ (unshaded) GWAS were compared. The number of unclumped genetic variants is shown in each venn diagram, with corresponding percentages in brackets. Various adjustment methods show a high degree of overlap compared to no adjustment, with the exception of the GWAS of ‘use of antihypertensives’ as a binary variable. Methods which applied direct adjustments to observed SBP values (add constant, add class-specific constant, censored normal regression) yielded at least 30% more unclumped top hits than no adjustment. In other methods which used restricted cohorts, much fewer unclumped top hits were identified, as a result of reduced power attributed to smaller sample sizes.

### **Supplementary Figure 22**. Venn diagrams of unclumped genetic variants of ‘Add constant +10mmHg’ (clumped reference) versus ‘Add class-specific constant’ and ‘Censored normal regression’ medication-adjusted GWAS

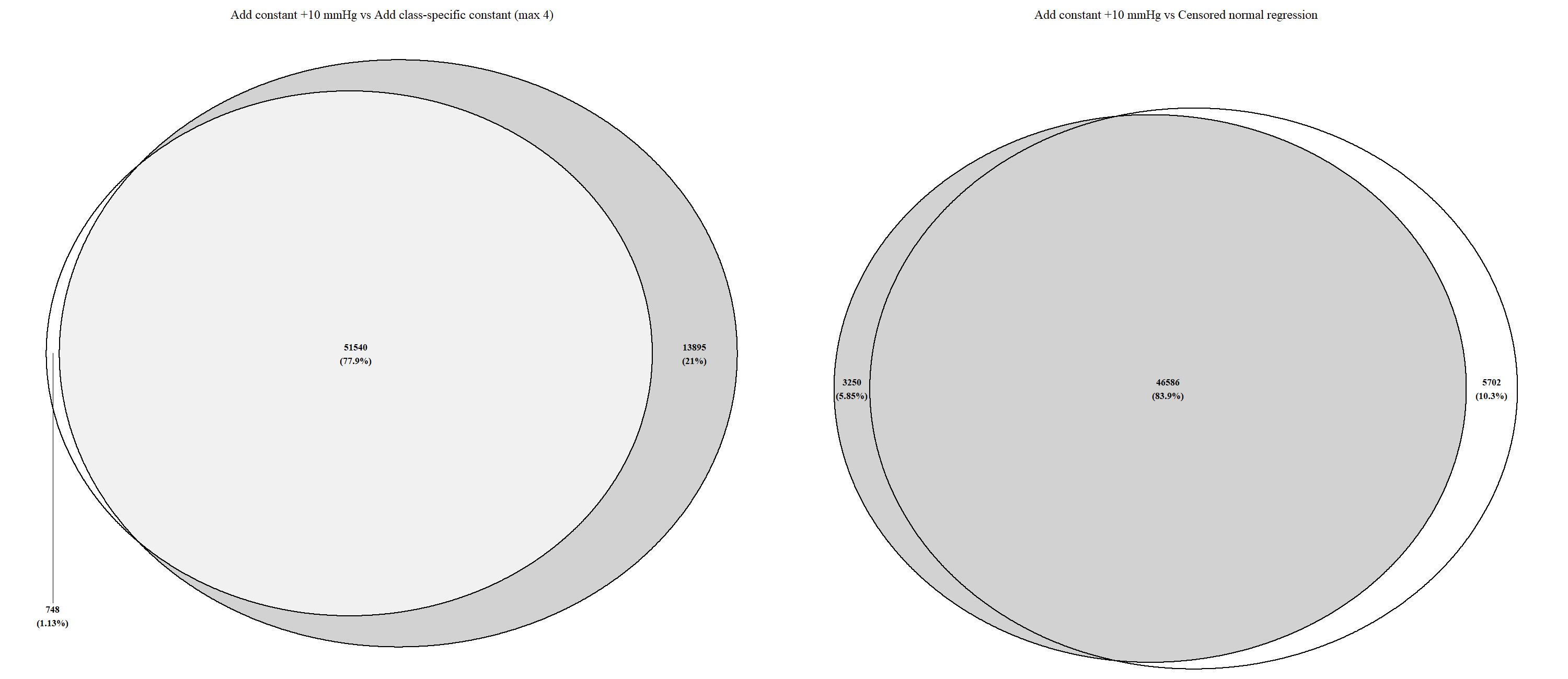

Unclumped top hits (p<5x10^-8^) from add constant +10mmHg adjusted GWAS (unshaded) versus add class-specific constant (shaded) and censored normal regression adjusted GWAS (shaded) were compared. The number of unclumped genetic variants is shown in each venn diagram, with corresponding percentages in brackets. There is a high degree of overlap among various adjustment methods, although add class-specific constant yielded more unclumped top hits as compared to add constant +10mmHg.

### **Supplementary Figure 23**. Scatter plot of independent genetic variants p-values of ‘Add constant +10mmHg’ (clumped reference) versus ‘Add class-specific constant’ medication-adjusted GWAS

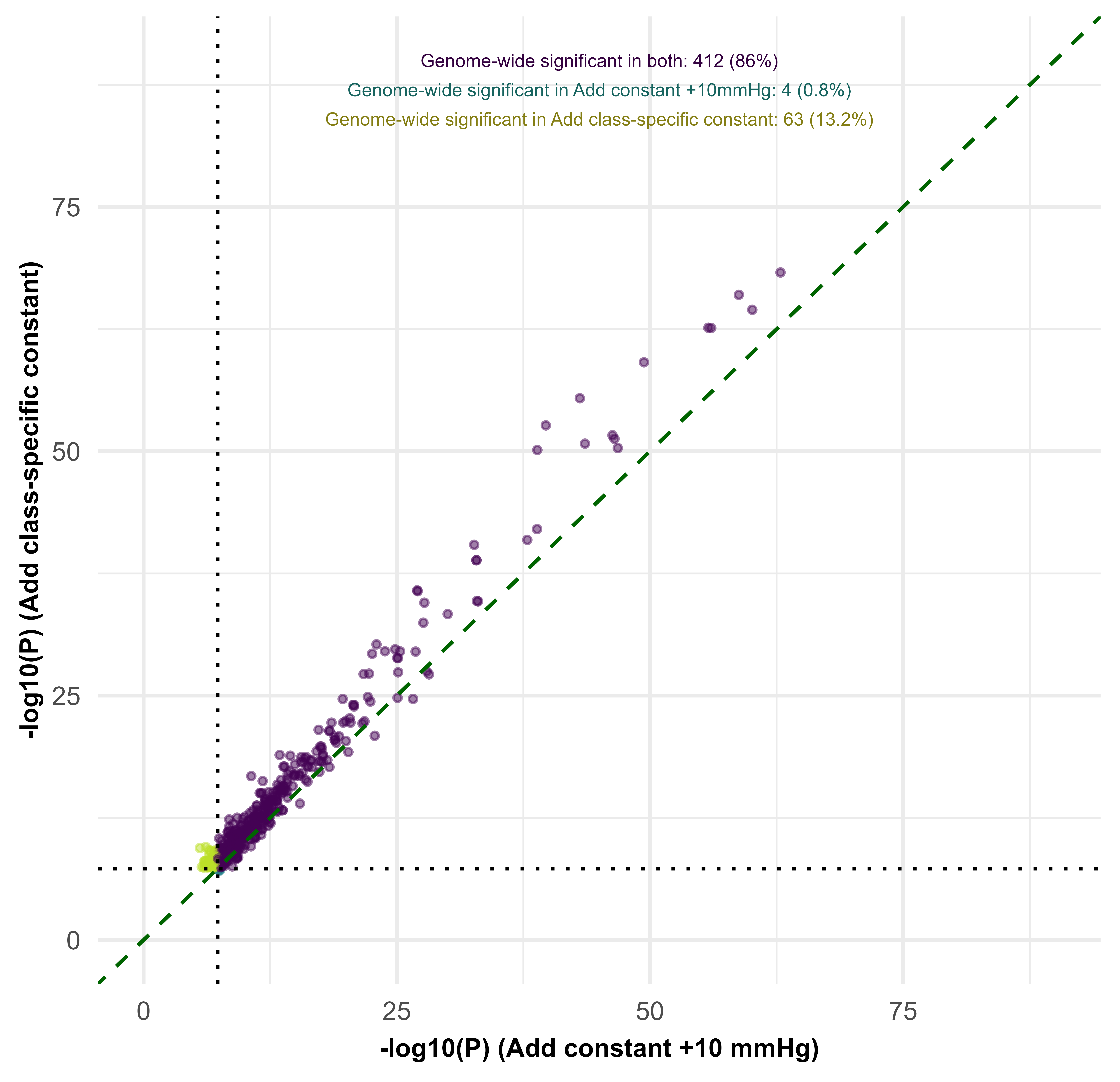

GWAS: genome-wide association study; P: p-value
The x-axis and y-axis shows the p-values in the ‘add constant +10mmHg’ medication-adjusted GWAS and ‘add class-specific constants’ medication-adjusted GWAS respectively. Each dot represents a clumped variant. Purple dots indicate variants which are genome-wide significant; Dark green dots show variants which are genome-wide significant in the ‘add constant +10mmHg’ GWAS but not the ‘add class-specific constant’ GWAS; Light green dots show variants which are genome-wide significant in the ‘add class-specific constant’ GWAS but not the ‘add constant +10mmHg’ GWAS. The dotted diagonal green line has a gradient of 1; if p-values from both GWAS are similar, we would expect to observe dots along the green line. A steeper slope greater than 1 indicates that the ‘add class-specific constant’ adjusted GWAS has a lower p-value than the ‘add constant +10mmHg’ adjusted GWAS, and vice-versa.

### **Supplementary Figure 24**. Scatter plot of independent genetic variants p-values of untreated versus treated GWAS

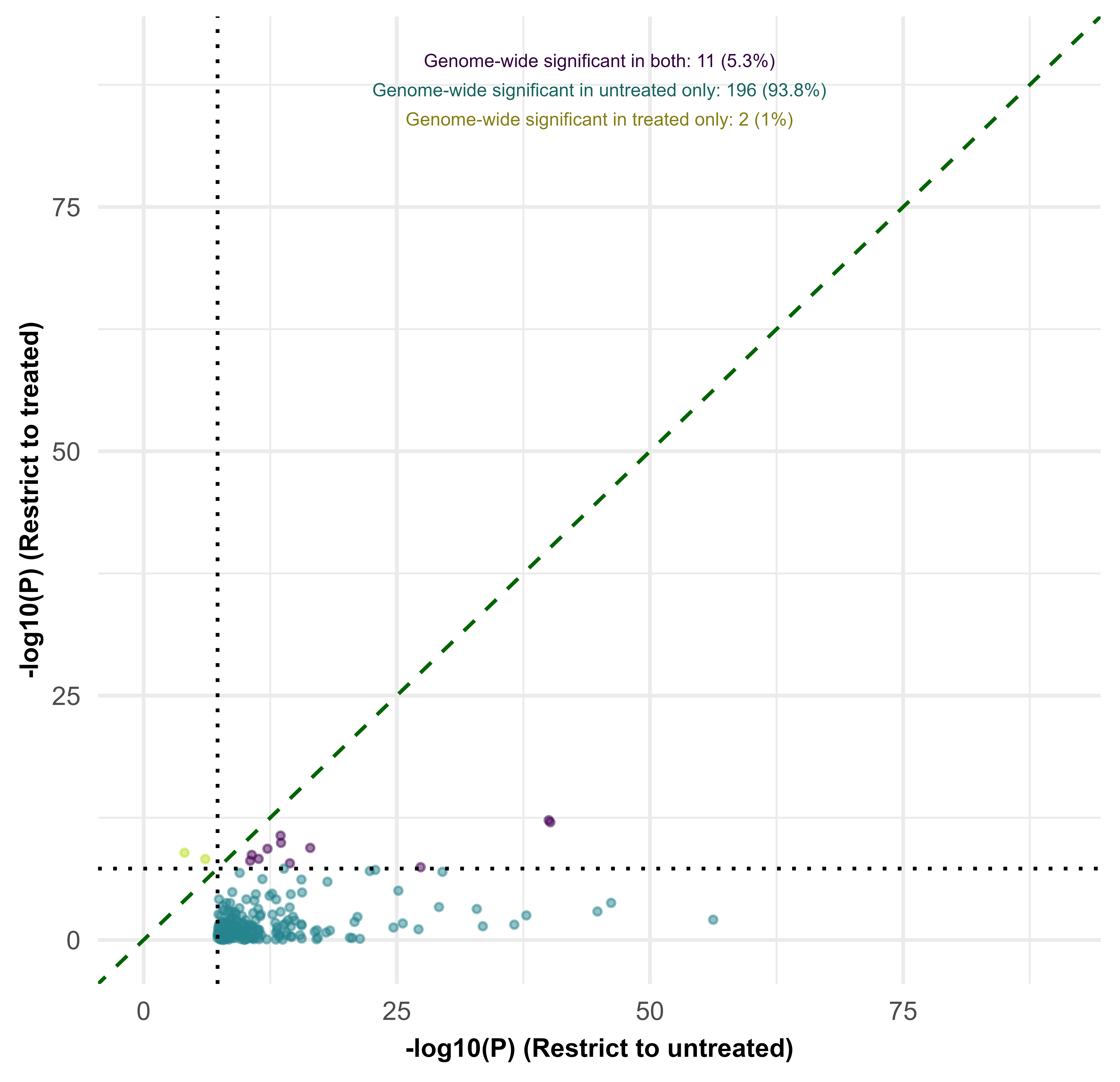

GWAS: genome-wide association study; P: p-value
The x-axis and y-axis shows the p-values in the untreated GWAS and treated GWAS respectively. Each dot represents a clumped variant. Purple dots indicate variants which are genome-wide significant; Dark green dots show variants which are genome-wide significant in the untreated GWAS but not the treated GWAS; Light green dots show variants which are genome-wide significant in the treated GWAS but not the untreated GWAS. The dotted diagonal green line has a gradient of 1; if p-values from both GWAS are similar, we would expect to observe dots along the green line. A steeper slope greater than 1 indicates that the treated GWAS has a more significant p-value than the untreated GWAS, and vice-versa.

### **Supplementary Figure 25**. Scatter plots of effect estimates of independent genetic variants from medication-adjusted (clumped reference) versus unadjusted GWAS

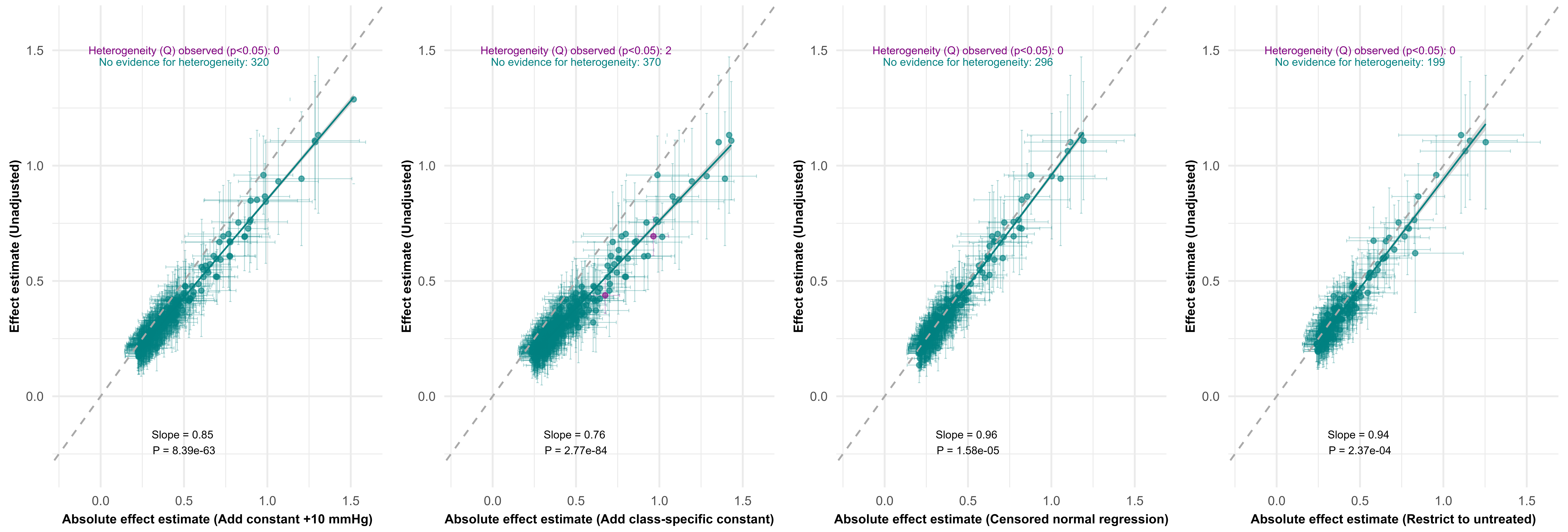

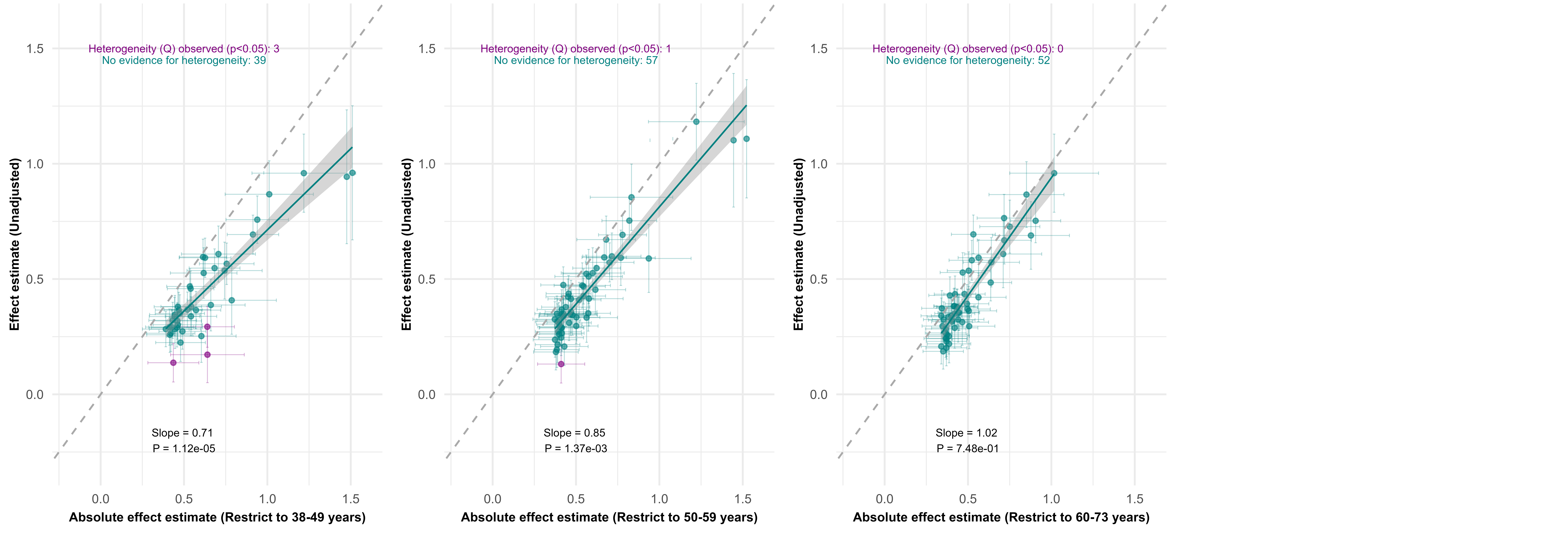

GWAS: genome-wide association study; P: p-value
The x-axis shows the absolute effect estimates of independent genetic variants from medication-adjusted GWAS (clumping was conducted on the medication-adjusted GWAS). The y-axis shows the corresponding effect estimates of these variants in the unadjusted GWAS. Each dot represents a single genetic variant with its corresponding standard error intervals. Green dots indicate that there is no evidence for heterogeneity of effect estimates for genetic variants. Purple dots indicate evidence (p<0.05) of heterogeneity between effect estimates for the same genetic variant in both GWAS after correction for multiple testing. The grey shaded region shows the 95% confidence intervals around the fitted regression line. The dotted green line has a gradient of 1; if effect estimates from both GWAS are similar, we will observe dots close to the green line. A positive slope >1 indicates stronger absolute effect estimates in the unadjusted GWAS, a positive slope <1 indicates stronger absolute effect estimated in the adjusted GWAS . The reported p-values are from a two-tailed t-test evaluating if the slope term differs from 1.

### **Supplementary Figure 26**. Scatter plots of independent genetic variants effect estimates of ‘Add constant +10mmHg’ (clumped reference) versus ‘Add class-specific constant’ medication-adjusted GWAS

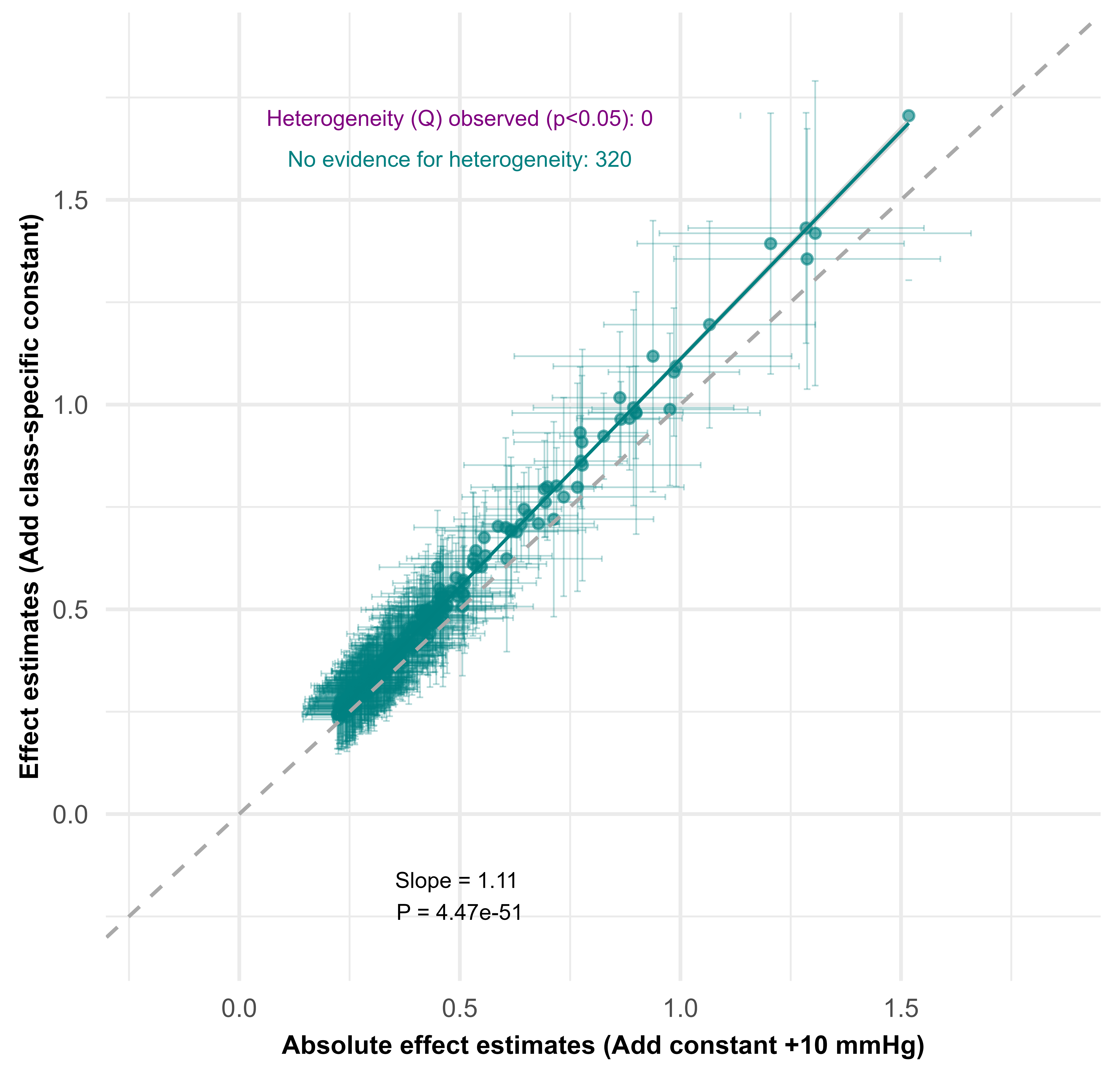

GWAS: genome-wide association study; P: p-value
The x-axis shows the absolute effect estimates of independent genetic variants from ‘add constant +10mmHg’ medication-adjusted GWAS (clumping was conducted on the ‘add constant +10mmHg’ GWAS). The y-axis shows the corresponding effect estimates of these variants in the ‘add class-specific constant’ medication-adjusted GWAS. Each dot represents a single genetic variant with its corresponding standard error intervals. Green dots indicate that there is no evidence for heterogeneity of effect estimates for genetic variants. The grey shaded region shows the 95% confidence intervals around the fitted regression line. The dotted green line has a gradient of 1; if effect estimates from both GWAS are similar, we will observe dots close to the green line. A positive slope >1 indicates stronger absolute effect estimates in the ‘add class-specific constant’ adjusted GWAS. The reported p-values are from a two-tailed t-test evaluating if the slope term differs from 1.

### **Supplementary Figure 27.** Scatter plots of independent genetic variants effect estimates of ‘Add class-specific constant’ (clumped reference) versus ‘Add constant +10mmHg’ medication-adjusted GWAS

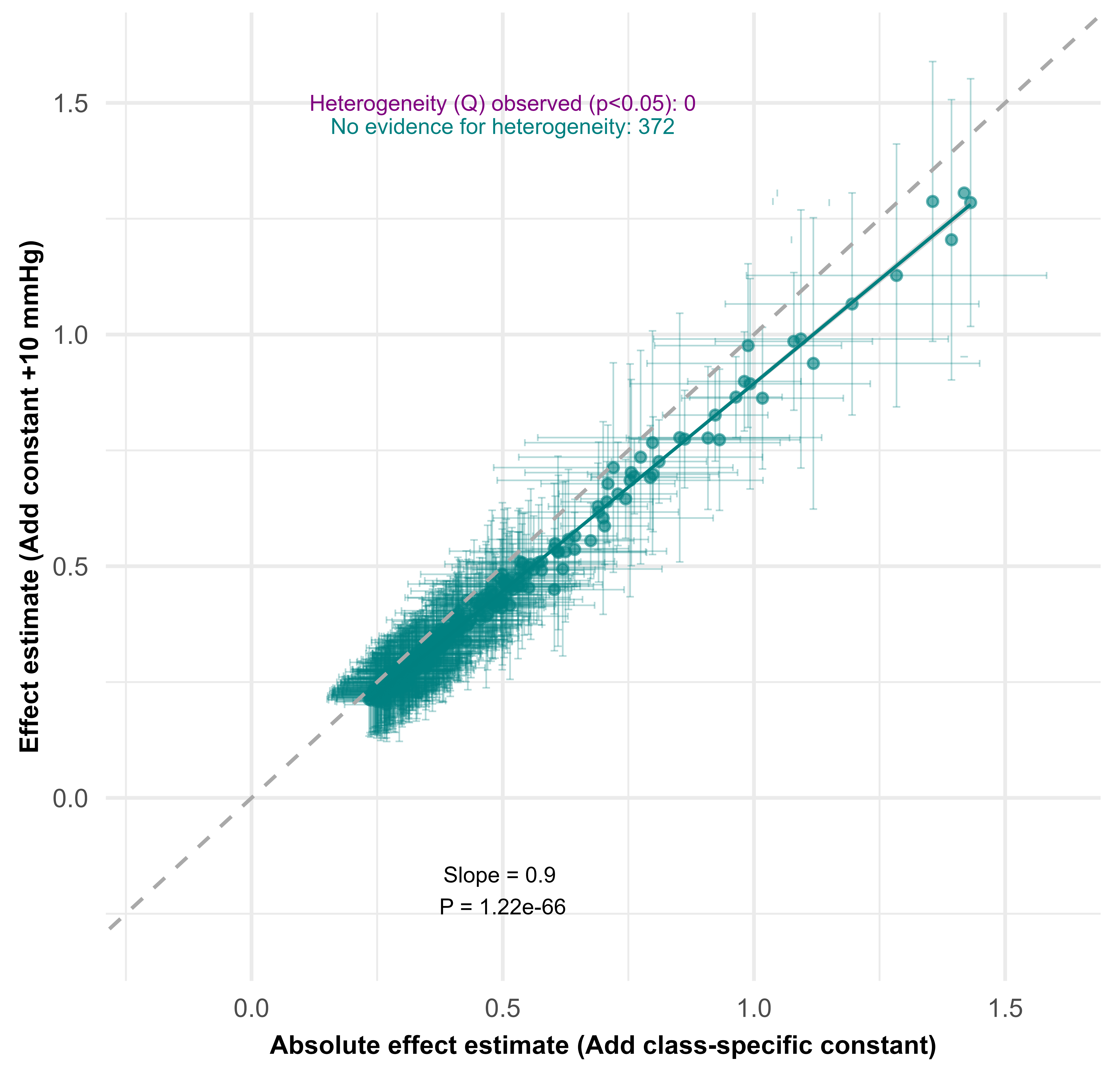

GWAS: genome-wide association study; P: p-value
The x-axis shows the absolute effect estimates of independent genetic variants from ‘add class-specific constant’ adjusted GWAS (clumping was conducted on the ‘add class-specific constant’ GWAS). The y-axis shows the corresponding effect estimates of these variants in the ‘add constant +10 mmHg’ adjusted GWAS. Each dot represents a single genetic variant with its corresponding standard error intervals. Green dots indicate that there is no evidence for heterogeneity of effect estimates for genetic variants. The grey shaded region shows the 95% confidence intervals around the fitted regression line. The dotted green line has a gradient of 1; if effect estimates from both GWAS are similar, we will observe dots close to the green line. A positive slope <1 indicates stronger absolute effect estimates in the ‘add class-specific constant’ adjusted GWAS. The reported p-values are from a two-tailed t-test evaluating if the slope term differs from 1.

### **Supplementary Figure 28**. Scatter plot of independent genetic variants effect estimates of treated (clumped reference) versus untreated GWAS

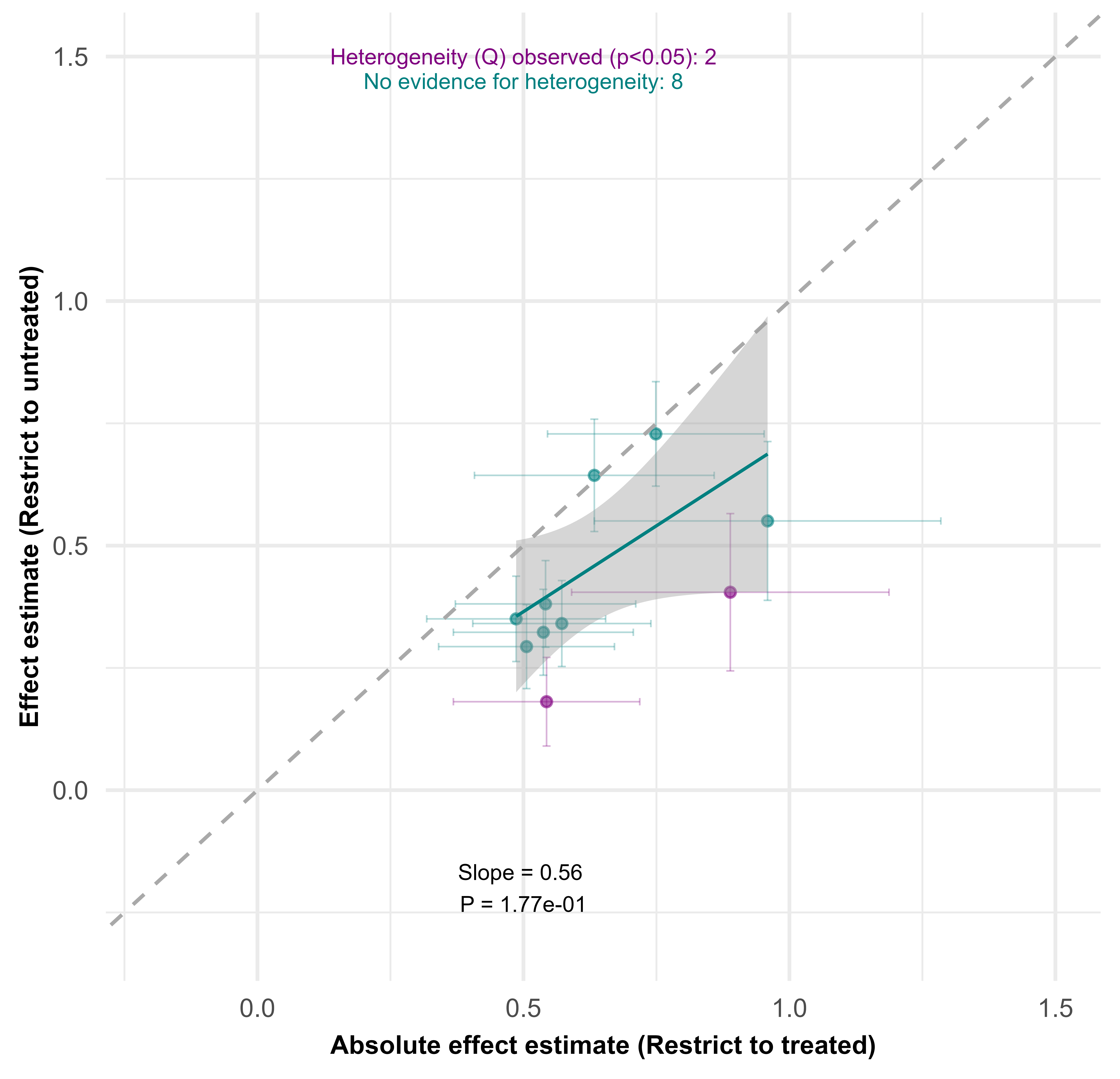

GWAS: genome-wide association study; P: p-value
The x-axis shows the absolute effect estimates of clumped top hits from GWAS restricted to treated individuals. The y-axis shows the corresponding effect estimates of these variants in the GWAS restricted to untreated individuals. Each dot represents a single genetic variant with its corresponding standard error intervals. Green dots indicate that there is no evidence for heterogeneity of effect estimates for genetic variants. Purple dots indicate evidence (p<0.05) of heterogeneity between effect estimates for the same genetic variant in both GWAS after correction for multiple testing. The grey shaded region shows the 95% confidence intervals around the fitted regression line. The dotted green line has a gradient of 1; if effect estimates from both GWAS are similar, we will observe dots close to the green line. A positive slope <1 indicates stronger absolute effect estimates in the GWAS restricted to treated individuals. The reported p-values are from a two-tailed t-test evaluating if the slope term differs from 1.

### **Supplementary Table 1**. Definition of antihypertensive use by BNF categories

| **Code** | **Description** |
| --- | --- |
| **Inclusion (Subparagraph)** | |
| 0205040 | Alpha-adrenoceptor blockers |
| 0205051 | Angiotensin-converting enzyme inhibitors |
| 0205052 | Angiotensin II receptor blockers |
| 0204000 | Beta-adrenoceptor blockers |
| 0206020 | Calcium channel blockers |
| 0205020 | Centrally acting antihypertensive drugs |
| 0202020 | Loop diuretics |
| 0202030 | Potassium-sparing diuretics and aldosterone antagonists |
| 0206010 | Nitrates |
| 0205053 | Renin inhibitors |
| 0202010 | Thiazides and related diuretics |
| 0205010 | Vasodilator antihypertensives |
| **Exclusion (Chemical substance)** | |
| 0205020g0 | Guanfacine hydrochloride |
| 0206010a0 | Amyl nitrite |
| 0206010f0 | Glyceryl trinitrate |
| 0205010aa | Macitentan |
| 0205010ab | Riociguat |
| 0205010ac | Vericiguat |
| 0205010u0 | Bosentan |
| 0205010v0 | Iloprost |
| 0205010w0 | Sitaxentan sodium |
| 0205010x0 | Ambrisentan |
| 0205010y0 | Sildenafil (Vasodilator Antihypertensive) |
| 0205010z0 | Tadalafil (Vasodilator Antihypertensive) |

### **Supplementary Table 2**. LD score regression results.

| **Medication-adjustment** | **N** | **SNPs**‡ | **SNP h^2^ (SE)** | **λ_GC** | **Mean χ²** | **Intercept (SE)** | **Ratio (SE)** |
| --- | --- | --- | --- | --- | --- | --- | --- |
| Unadjusted | 407960 | 1160310 | 0.1315 (0.0048) | 1.7688 | 2.2080 | 1.1173 (0.0144) | 0.0971 (0.0119) |
| Add constant +10 mmHg | 407960 | 1160310 | 0.1617 (0.0056) | 1.9136 | 2.5015 | 1.1538 (0.0158) | 0.1025 (0.0105) |
| Add class-specific constant† | 407960 | 1160310 | 0.1822 (0.0061) | 1.9923 | 2.6969 | 1.1759 (0.0163) | 0.1037 (0.0096) |
| Censored normal regression | 407960 | 1160310 | 0.1558 (0.0055) | 1.8891 | 2.4437 | 1.1453 (0.0154) | 0.1006 (0.0107) |
| Restrict to untreated | 314510 | 1160319 | 0.1453 (0.0056) | 1.6715 | 2.0182 | 1.0896 (0.0134) | 0.0880 (0.0131) |
| Restrict to treated | 93450 | 1160343 | 0.0885 (0.0074) | 1.1459 | 1.1742 | 1.0110 (0.0076) | 0.0631 (0.0435) |
| Restrict to 38-49 years | 88772 | 1160256 | 0.1880 (0.0109) | 1.2731 | 1.3550 | 1.0241 (0.0086) | 0.0680 (0.0241) |
| Restrict to 50-59 years | 134297 | 1160307 | 0.1533 (0.0086) | 1.3374 | 1.4553 | 1.0388 (0.0099) | 0.0852 (0.0218) |
| Restrict to 60-73 years | 184891 | 1160350 | 0.1235 (0.0058) | 1.3650 | 1.4925 | 1.0384 (0.0104) | 0.0780 (0.0211) |

†Up to a maximum of 4 medication classes
‡SNPs retained after restricting to HapMap3 SNPs and pre-computed European Scores from 1000 Genomes

### **Supplementary Table 3.** Sensitivity analyses for effect of systolic blood pressure on coronary artery disease

| **Adjustments to use of antihypertensives** | **N SNPs** | **IVW**  **OR (95% CI)** | **MR Egger OR (95% CI)** | **Weighted median**  **OR (95% CI)** | **Weighted**  **mode**  **OR (95% CI)** |
| --- | --- | --- | --- | --- | --- |
| Exposure: SBP, Outcome: Coronary artery disease | | | | | |
| (1) Unadjusted | 227 | 1.92  (1.65-2.24) | 1.94  (1.23-3.05) | 1.81  (1.58-2.08) | 1.74  (1.25-2.42) |
| (2) Add constant +10 mmHg | 302 | 1.92  (1.71-2.16) | 1.81  (1.28-2.54) | 1.81  (1.62-2.03) | 1.68  (1.22-2.32) |
| (3) Add class-specific constant | 348 | 1.91  (1.73-2.11) | 1.60  (1.21-2.12) | 1.83  (1.65-2.03) | 1.78  (1.32-2.38) |
| (4) Censored normal regression | 284 | 1.92  (1.69-2.18) | 1.80  (1.24-2.63) | 1.84  (1.64-2.08) | 1.68  (1.22-2.32) |
| (5) Restrict to untreated | 189 | 1.93  (1.69-2.19) | 1.78  (1.18-2.69) | 1.71  (1.49-1.95) | 1.61  (1.21-2.13) |
| (6) Restrict to 38-49 years | 40 | 1.82  (1.50-2.19) | 1.67  (0.82-3.38) | 1.82  (1.54-2.15) | 2.15  (1.54-3.00) |
| (7) Restrict to 50-59 years | 52 | 1.67  (1.37-2.04) | 1.68  (0.85-3.31) | 1.50  (1.27-1.78) | 1.50  (1.11-2.03) |
| (8) Restrict to 60-73 years | 48 | 1.43  (1.11-1.85) | 1.46  (0.55-3.89) | 1.38  (1.14-1.68) | 1.36  (1.02-1.80) |
| (9) MVMR with binary treatment status as a second exposure | 262 | 1.07  (0.82-1.40) | - | - | - |

IVW: inverse-variance weighted; MVMR: multivariable mendelian randomization; SBP: systolic blood pressure

### **Supplementary Table 4**. Sensitivity analyses for effect of body mass index on systolic blood pressure

| **Adjustments to use of antihypertensives** | **N SNPs** | **IVW**  **Beta**  **(95% CI)** | **MR Egger**  **Beta**  **(95% CI)** | **Weighted median**  **Beta**  **(95% CI)** | **Weighted**  **mode**  **Beta**  **(95% CI)** |
| --- | --- | --- | --- | --- | --- |
| Exposure: BMI, Outcome: SBP | | | | | |
| (1) Unadjusted | 78 | 1.29 (0.39-2.19) | -0.01  (-2.19-2.16) | 1.81  (1.13-2.49) | 1.92  (0.98-2.85) |
| (2) Add constant +10 mmHg | 78 | 1.98  (0.96-3.01) | 0.50  (-1.99-2.98) | 2.80  (2.06-3.54) | 2.82  (1.81-3.83) |
| (3) Add class-specific constant | 78 | 2.63  (1.49-3.76) | 1.06  (-1.70-3.81) | 3.16  (2.29-4.02) | 3.61  (2.46-4.76) |
| (4) Censored normal regression | 78 | 2.21  (1.31-3.10) | 0.89  (-1.29-3.06) | 2.82  (2.17-3.47) | 3.00  (2.05-3.95) |
| (5) Restrict to untreated | 78 | 1.45  (0.53-2.37) | 0.31  (-1.92-2.55) | 2.27  (1.48-3.06) | 2.36  (1.43-3.29) |
| (6) Restrict to 38-49 years | 78 | 2.76  (1.71-3.82) | 2.51  (-0.08-5.11) | 3.51  (2.36-4.67) | 4.39  (2.58-6.20) |
| (7) Restrict to 50-59 years | 78 | 1.75  (0.60-2.90) | 0.48  (-2.31-3.27) | 1.18  (0.12-2.25) | 1.08  (-0.44-2.61) |
| (8) Restrict to 60-73 years | 78 | 0.21  (-0.75-1.17) | -1.66  (-3.97-0.64) | 0.16  (-1.04-1.36) | 0.29  (-1.15-1.73) |

BMI: body mass index; IVW: inverse-variance weighted; SBP: systolic blood pressure

### **Supplementary Table 5**. Sensitivity analyses for effect of systolic blood pressure on natural hair colour

| **Adjustments to use of antihypertensives** | **N SNPs** | **IVW**  **Beta**  **(95% CI)** | **MR Egger**  **Beta**  **(95% CI)** | **Weighted median**  **Beta**  **(95% CI)** | **Weighted**  **mode**  **Beta**  **(95% CI)** |
| --- | --- | --- | --- | --- | --- |
| Exposure: SBP, Outcome: Natural hair colour | | | | | |
| (1) Unadjusted | 233 | 0.002  (0.000-0.003) | -0.002  (-0.008-0.003) | 0.001  (0.000-0.003) | 0.001  (-0.003-0.005) |
| (2) Add constant +10 mmHg | 304 | 0.001  (-0.001-0.006) | -0.001  (-0.005-0.003) | 0.001  (0.000-0.002) | 0.001  (-0.002-0.005) |
| (3) Add class-specific constant | 354 | 0.001  (-0.001-0.002) | 0.000  (-0.004-0.003) | 0.001  (0.000-0.002) | 0.002  (-0.001-0.005) |
| (4) Censored normal regression | 288 | 0.002  (0.000-0.003) | -0.003  (-0.008-0.001) | 0.001  (0.000-0.002) | 0.001  (-0.003-0.006) |
| (5) Restrict to untreated | 192 | 0.002  (0.000-0.003) | -0.002  (-0.008-0.003) | 0.002  (0.000-0.003) | 0.003  (-0.001-0.007) |
| (6) Restrict to 38-49 years | 41 | 0.000  (-0.004-0.004) | -0.015  (-0.028--0.002) | 0.001  (-0.001-0.003) | 0.002  (-0.001-0.005) |
| (7) Restrict to 50-59 years | 54 | 0.001  (-0.002-0.004) | 0.000  (-0.010-0.011) | 0.002  (0.000-0.004) | 0.003  (-0.001-0.007) |
| (8) Restrict to 60-73 years | 48 | 0.001  (-0.002-0.004) | -0.007  (-0.020-0.006) | 0.001  (-0.001-0.003) | 0.001  (-0.003-0.005) |
| (9) MVMR with binary treatment status as a second exposure | 269 | 0.006  (0.001-0.012) | - | - | - |

IVW: inverse-variance weighted; MVMR: multivariable mendelian randomization; SBP: systolic blood pressure

### **Supplementary Table 6**. F-statistics for univariable MR IVW analyses

| **Adjustments to use of antihypertensives** | **N SNPs** | **Mean F-statistic** | **Min F-statistic** | **Max F-statistic** |
| --- | --- | --- | --- | --- |
| Exposure: SBP, Outcome: Coronary artery disease | | | | |
| (1) Unadjusted | 227 | 55.5 | 29.8 | 263.6 |
| (2) Add constant +10 mmHg | 302 | 60.0 | 29.8 | 377.0 |
| (3) Add class-specific constant | 348 | 62.5 | 29.9 | 422.5 |
| (4) Censored normal regression | 284 | 59.4 | 29.9 | 361.8 |
| (5) Restrict to untreated | 189 | 53.2 | 29.8 | 253.0 |
| (6) Restrict to 38-49 years | 40 | 46.9 | 29.9 | 134.9 |
| (7) Restrict to 50-59 years | 52 | 45.9 | 30.4 | 112.4 |
| (8) Restrict to 60-73 years | 48 | 48.7 | 30.0 | 139.6 |
| Exposure: BMI, Outcome: SBP | | | | |
| BMI | 78 | 66.0 | 29.9 | 716.5 |
| Exposure: SBP, Outcome: Natural hair colour | | | | |
| (1) Unadjusted | 233 | 55.2 | 29.8 | 263.6 |
| (2) Add constant +10 mmHg | 304 | 60.1 | 29.8 | 377.0 |
| (3) Add class-specific constant | 354 | 62.9 | 29.9 | 422.5 |
| (4) Censored normal regression | 288 | 59.4 | 29.9 | 361.8 |
| (5) Restrict to untreated | 192 | 53.2 | 29.8 | 253.0 |
| (6) Restrict to 38-49 years | 41 | 47.2 | 29.9 | 134.9 |
| (7) Restrict to 50-59 years | 54 | 46.0 | 30.4 | 112.4 |
| (8) Restrict to 60-73 years | 48 | 48.7 | 30.0 | 139.6 |

BMI: body mass index, IVW: inverse-variance weighted; MR: mendelian randomization; SBP: systolic blood pressure

### **Supplementary Text 1**. LD score regression

We performed LD score regression to all the unadjusted and medication-adjusted GWAS, to check for genomic inflation and calculate SNP-based heritability. This was done by using Bulik-Sullivan et al. LDSC version 1.01. We restricted the analysis to HapMap3 SNPs and genetic variants with MAF>0.01. All other default settings were applied, such as removing duplicated or non-matching rsids and strand-ambiguous SNPs. LD scores were computed from European 1000 Genomes Phase 3 reference panel, and heritability was estimated on the observed scale. For each GWAS, we reported the SNP h^2^ estimate with its standard error, Lambda GC, Mean Chi-square, LDSC intercept and ratio. The LDSC intercept and ratio was used to assess for genomic inflation, and the proportion which can be ascribed to causes other than polygenic heritability.

### **Supplementary Text 2**. Characterisation of additional independent genetic variants

For direct medication-adjustment methods (add constant +10 mmHg, add class-specific constants, censored normal regression), we identified additional independent genetic variants detected using medication adjustment methods but not in unadjusted analyses. For each additional independent genetic variant, we computed the physical distance and the LD r^2^ to the nearest unadjusted index variant. For purposes of classifying whether the additional variant was independent of the unadjusted analysis, we applied the following threshold: more than 1,000 kb from every unadjusted index variant, or within 1,000 kb but with r² < 0.1. We annotated nearest genes and functional consequences using Ensembl Variant Effect Predictor REST API (GRCh37 assembly).

To understand whether each additional independent genetic variant had been previously published, we used two resources: the NHGRI-EBI GWAS Catalog and OpenGWAS. The NHGRI-EBI GWAS Catalog was accessed through the ‘gwasrapidd’ R package. We searched for ‘EFO_0006335’ which is indicative of systolic blood pressure studies, but added exclusions where the trait was not solely systolic blood pressure (e.g., interaction studies, childhood, response, preeclampsia etc.). For OpenGWAS, we searched for ‘systolic blood pressure’ trait, via the ‘ieugwasr’ package. In both resources, we filtered for genetic variants which were genome-wide significant. We recorded that the genetic variant was previously published if it met the genome-wide significant threshold, and had a PubMedID record in the databases.

We also compared if these variants had reported heterogeneous effect estimates between unadjusted and medication-adjusted analyses, as well as between treated and untreated analyses.
